# The relationship between genotype- and phenotype-based estimates of genetic liability to psychiatric disorders, in practice and in theory

**DOI:** 10.1101/2023.06.19.23291606

**Authors:** Morten Dybdahl Krebs, Vivek Appadurai, Kajsa-Lotta Georgii Hellberg, Henrik Ohlsson, Jette Steinbach, Emil Pedersen, iPSYCH Study Consortium, Thomas Werge, Jan Sundquist, Kristina Sundquist, Na Cai, Noah Zaitlen, Andy Dahl, Bjarni Vilhjalmsson, Jonathan Flint, Silviu-Alin Bacanu, Andrew J. Schork, Kenneth S. Kendler

**Affiliations:** Institute of Biological Psychiatry, Mental Health Center - Sct Hans, Copenhagen University Hospital – Mental Health Services CPH, Copenhagen, Denmark; Center for Primary Health Care Research, Lund University, Malmö, Sweden; NCRR - National Centre for Register-Based Research, Business and Social Sciences, Aarhus University, Aarhus V, Denmark; Section for Geogenetics, GLOBE Institute, Faculty of Health and Medical Science, Copenhagen University; Helmholtz Pioneer Campus, Helmholtz Zentrum München, Neuherberg, Germany; Department of Neurology, University of California, Los Angeles, California 90024, USA; Department of Computational Medicine, University of California, Los Angeles, California, USA; Section of Genetic Medicine, Department of Medicine, University of Chicago, Chicago, Illinois, USA; Department of Biomedicine, Aarhus University, Aarhus, Denmark; Center for Neurobehavioral Genetics, Semel Institute for Neuroscience and Human Behavior, University of California, Los Angeles, CA, USA; Virginia Institute for Psychiatric and Behavioral Genetics, Virginia Commonwealth University, Richmond, VA, USA; Department of Psychiatry, Virginia Commonwealth University, Richmond, VA, USA

**Author notes:** Please address correspondence to: Morten Krebs, Andrew J. Schork, Andrew, Kenneth S. Kendler. iPSYCH Study Consortium authors can be found in the Supplementary Table S0. These authors jointly supervised this work (Andrew J. Schork, Kenneth S. Kendler).

## Abstract

Genetics as a science has roots in studying phenotypes of relatives, but molecular approaches facilitate direct measurements of genomic variation within individuals. Agricultural and human biomedical research are both emphasizing genotype-based instruments, like polygenic scores, as the future of breeding programs or precision medicine and genetic epidemiology. However, unlike in agriculture, there is an emerging consensus that family variables act nearly independent of genotypes in models of human disease. To advance our understanding of this phenomenon, we use 2,066,057 family records of 99,645 genotyped probands from the iPSYCH2015 case-cohort study to show that state-of-the-field genotype- and phenotype-based genetic instruments explain largely independent components of liability to psychiatric disorders. We support these empirical results with novel theoretical analysis and simulations to describe, in a human biomedical context, parameters affecting current and future performance of the two approaches, their expected interrelationships, and consistency of observed results with expectations under simple additive, polygenic liability models of disease. We conclude, at least for psychiatric disorders, that phenotype- and genotype-based genetic instruments are likely noisy measures of the same underlying additive genetic liability, should be seen for the near future as complementary, and integrated to a greater extent.

## Introduction

In the first decades of genetics as a science, the only approach for estimating genetic contributions to traits was by assessing phenotypes of relatives^1,2^, but now molecular approaches provide direct measures of individual genomes cheaply, efficiently, and at scale. In agricultural applications, e.g., estimating breeding values of dairy cattle^3,4^, “genotype-based genetic instruments” derived from molecular data are proving so effective that “phenotype-based genetic instruments” obtained from pedigree records are becoming redundant^3,5–7^. At the same time, human biomedical research is emphasizing genotype-based instruments, polygenic scores (PGS)^8^, in precision medicine^9–11^ and genetic epidemiology^12^. However, it is not known how and why phenotype-based and genotype-based genetic instruments complement each other in human biomedical research. We examine these questions by combining theoretical analysis, simulations, and applications of state-of-the-field genotype- and phenotype-based genetic instruments in a unique psychiatric genetics data resource.

Somewhat counterintuitively, PGS and indicators of positive family history (FH) have low correlations and contribute nearly independently to classification and prediction. PGS and parental history for depression^13^, autism^14^, or schizophrenia^15^ are near-independent in epidemiological models, with similar results in others diseases^16,17^. Studies^18–20^ using family genetic risk scores (FGRS), shared environment adjusted, kinship weighted sums of diagnoses in proband genealogies, have reported a number of associations that are mirrored by studies using PGS (e.g.,^21–24^). Model-based estimates of liability from family records, liability threshold on family history (LT-FH)^16^ and Pearson-Aitken Family Genetic Risk Scores (PA-FGRS)^21^, have low correlations with PGS and contribute nearly independently to classification. Many intuit this as evidence that family variables reflect effects of shared environment but it is not clear that these reports are inconsistent with large additive genetic components in complex trait etiology^25,26^.

In this work we describe the relationships among phenotype-based genetic instruments (FH and PA-FGRS), genotype-based genetic instruments (PGS), and liability for five major psychiatric disorders. We use data from two independent cohorts of the iPSYCH2015 case-cohort study that include diagnostic information for 99,645 genotyped probands (62,357 psychiatric cases, 37,288 controls) and 2,066,657 relatives. We propose that the lack of redundancy between PA-FGRS (or FH) and PGS in human biomedical research is consistent with the hypothesis that both are simply very noisy measures of a highly similar underlying genetic liability. Different from trends in agriculture applications, we suggest that such instruments should be emphasized as complementary, at least over the near-term, and should be better-integrated going forward.

## Results

### Genotype- and phenotype-based genetic instruments explain nearly independent components of liability to psychiatric disorders

Genotype-based and phenotype-based genetic instruments are both known to contribute to models of human disease. Here, we estimate the variance in liability to five psychiatric disorders explained by combinations of polygenic scores (PGS), indicators of positive family-history (FH), and Pearson-Aitken Family Genetic Risk Scores (PA-FGRS) using a two-stage sampling weighted probit regression (Online Methods; Figure 1; Supplementary Table S1, S2). We introduce the two-stage sampling weighted probit as a preferred estimating approach because common estimators^27^ produced qualitatively different results when applied to our real data (Supplementary Figure S1) and subsequent simulation experiments suggested our new approach to be the least biased (Supplementary Information S1; Supplementary Figure S2; Supplementary Table S3). Nagelkirke’s pseudo-R^2^ from logistic models are shown in Supplementary Figure S3. In these models, PGS, FH, and PA-FGRS each explain significant variance in liability to each disorder, a result that replicates across two independent samples (Figure 1A,B, orange, yellow, green bars). For all disorders, PA-FGRS explain more liability than FH (mean increase 30%, range 11% to 46%; Supplementary Table S2). The variance explained by PGS and PA-FGRS (or FH) when fit together is nearly the sum of variance explained by PGS and PA-FGRS (or FH) when fit alone (Figure 1A,B, blue and purple; Supplementary Table S2). Including FH did not increase the variance explained of models including PA-FGRS and PGS (4^th^ vs. 6^th^ bars in each set, Figure 1A,B, Supplementary Table S2; exception: MDD in iPSYCH 2012). However, including PA-FGRS in models with PGS and FH did generally result in small but significant increases in variance explained (5^th^ vs. 6^th^ bars in each set, Figure 1A,B, Supplementary Table S2; exceptions: BPD, SCZ). Estimated odds ratios (OR) for PA-FGRS (or PGS) were only modestly reduced (mean reduction: 5%) when adjusting for PGS (or PA-FGRS, mean reduction: 8%; Figure 1C,D, Supplementary Table S3). Estimated ORs of FH adjusting for PA-FGRS were reduced to a larger extent (mean: 72%) than ORs of PA-FGRS when adjusting for FH (mean: 23%; Supplementary Figure S4-5, Supplementary Table S4). Taken together, this suggests PA-FGRS is a better instrument for capturing familial genetics than FH. Finally, we observe quite modest correlations between PGS and PA-FGRS in random population samples (Pearson correlation coefficient (r), 0.03 to 0.09; Figure 1E,F) that were consistently, but modestly, larger than between PGS and FH (r, 0.02 to 0.07), and much lower than between PA-FGRS and FH (r, 0.72 to 0.82, Supplementary Figures S6-7).

**Figure 1.**
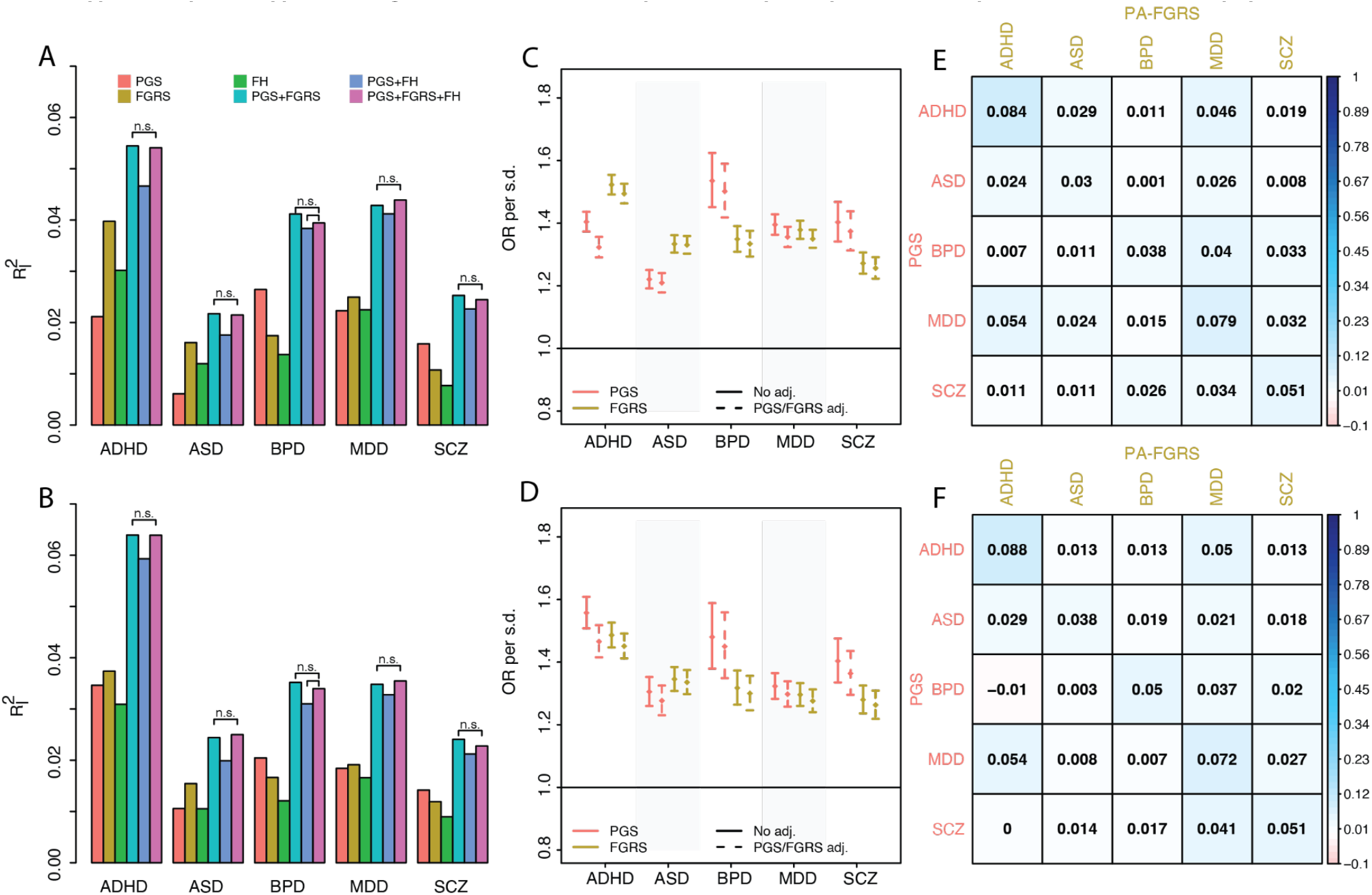
Genotype- and phenotype-based genetic instruments explain nearly independent components of liability to psychiatric disorders. Multiple genetic instruments, alone and in combination, explain variance on the liability scale for five major psychiatric disorders after accounting for covariates in two independent cohorts: (A) the iPSYCH2012 cohort and (B) the iPSYCH2015i cohort. Increases in variance explained associated with adding each variable to each model was tested via permutations and all were significant after Bonferroni correction (p < 0.05/110 = 4.55 × 10^−4^) unless annotated with n.s.. For (C) iPSYCH2012 and (D) iPSYCH2015i cohorts, the unadjusted (solid line) and adjusted (dashed line) OR for PGS and PA-FGRS in logistic models are similar for all disorders. In the (E) iPSYCH2012 and (F) iPSYCH2015i random population sub-cohorts, the pearson correlations among PGS and PA-FGRS instruments are modest, in general, but slightly larger within disorder. ADHD, Attention deficit hyperactivity disorder; ASD, autism spectrum disorder; BPD, bipolar disorder; MDD, major depressive disorder; SCZ, schizophrenia; PGS, polygenic scores; FH, family history indicators; PA-FGRS, Pearson-Aitken Family Genetic Risk Scores; n.s., no significant difference; OR, odds ratio; adj., adjustment; s.d., standard deviation.

### The current, future, and asymptotic performance of PA-FGRS depends on pedigree structure and trait architecture

In order to understand how and why these instruments appear independent, we need to understand how PA-FGRS is expected to behave in data and under generative models relevant to human biomedical research. Assuming the simple polygenic liability threshold model, we derived (Supplementary Information S2-6) expectations for the *accuracy* of PA-FGRS (*r*_*g,ĝPA-FGRS′*_, the correlation between our instrument, ĝ_*PA-FGRS′*_ and the true latent genetic value, *g*; Online Methods, Eqs. 9-10) as well as for the *performance* of PA-FGRS (*R*^*2*^_l,_ĝ_*PA-FGRS′*_ the variance in total liability, *L*, explained by the instrument; Online Methods). We use these equations to describe the expected behavior of PA-FGRS in predicting four hypothetical traits from human-relevant pedigree data (Figure 2A-F; Supplementary Figures S8-S16). First, PA-FGRS will never capture all additive genetic effects in human pedigrees for two well-described^6,28^ reasons: 1) unless a proband has *half-sibling offspring* (i.e., offspring from different mates such that they are half-siblings to each other) PA-FGRS becomes an estimate of between family genetic variance (i.e., mid-parental genotype, g_MPV_) and the accuracy and performance of bounded 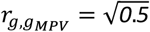 and 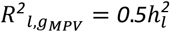 (Figures 2A,B, dashed lines), and 2) it would require unrealistically many half-sibling offspring to achieve full accuracy 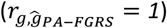 and performance 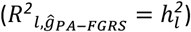 (Figures 2C,D, dashed lines). As such, we expect that incorporating additional phenotypes of full siblings (Figure 2A,B) to be less useful than such half-sibling offspring (Figure 2C,D). For plausible human pedigrees (N^rel^ < 10), the accuracy and performance of PA-FGRS are well below theoretical maxima (Figure 2A-D, Supplementary Figures S8-S13). Second, PA-FGRS that use complex, extended pedigrees do not gain information beyond four generations of records (i.e., beyond great-grandparents and their descendants; Figure 2E-H). Finally as an intuitive measure of information in a pedigree, we use the expected accuracy of PA-FGRS to define a *number of full sibling equivalents* (N_sibe_; Online Methods, Eq. 12) from any arbitrary set of relatives (Figure 2I,J). For a trait with heritability of 0.7 and prevalence of 0.01, N_sibe_ of 5 is achieved by ∼1 monozygotic twin, ∼4 half-sibling offspring, or approximately one entire great-grandparent founded pedigree with two children per generation (for other simulated trait architectures see, Supplementary Figures S17). Family diagnoses are always more informative for common traits (e.g., Figure 2A-H, purple vs. orange lines), but more distant relatives (e.g., half-siblings or cousins, green or blue lines, respectively, Figure 2I,J) are *relatively* more predictive for rarer traits (i.e., correspond to higher N_sibe_), while closer relatives (i.e., half-sibling offspring, yellow solid lines, Figure 2I,J) are relatively more informative for common traits. Simulations including shared (or “familial”) environmental contributions from siblings and parents show that PA-FGRS will incorporate confounded genetic and familial variance into predictions and this results in overperformance (i.e., excess variance explained) relative to the level expected from genetics alone (Supplementary Figures S18-S21). PA-FGRS will perform best for highly heritable, common traits and with many close relatives, but even in optimistic human pedigree data (N_sibe_=5) it will explain a modest proportion of heritability.

**Figure 2.**
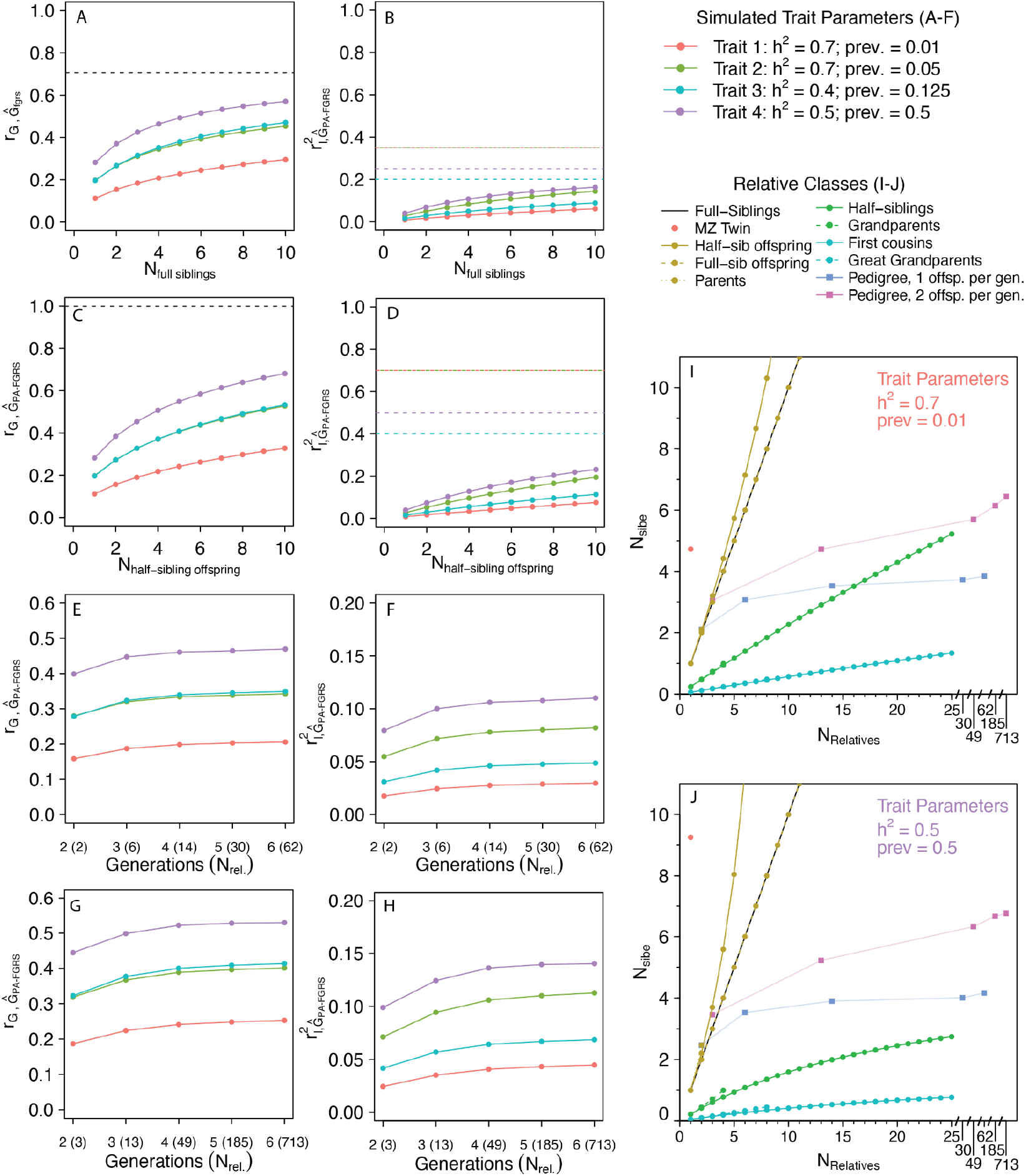
The expected and asymptotic accuracy and performance of PA-FGRS depend on pedigree structure, trait architecture, and realistic bounds on numbers of observed relatives. The (A) expected accuracy of PA-FGRS 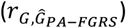 from records of increasing numbers of full siblings varies as a function of trait h^2^ and prev. with a theoretical asymptote at 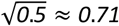 (dashed line) and a practical asymptote (N_Full sibliings_ < 10) well below this value. This is mirrored by trends in (B) expected performance of full sibling PA-FGRS 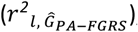, with theoretical asymptote of 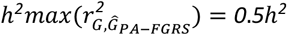, and a practical asymptote (N_Full sibliings_ < 10) well below. The (C) expected accuracy and (D) performance of PA-FGRS incorporating records of half-sibling offspring vary by h^2^ and prev., have, uniquely, theoretical asymptotes at 1 and *h*^*2*^, respectively, but practical asymptotes (N_half-sibling offspring_ < 10) well below these values. Multi-generation pedigrees that assume one (E,F) or two (G,H) children per mate pair are bounded in accuracy (E,G) and performance (F,H) by the same principles, with practical asymptotes occurring at approximately four generations of pedigree depth and well below theoretical maxima (depicted in A,B). The number of full sibling equivalents (N_sibe_) is a measure of information contained in an arbitrary set of relatives that can be used to compare and benchmark estimates of accuracy from familial configurations (I,J). Relative to full siblings, distant relatives are more important for rare traits (I) and close relatives are more important for common traits (J). PA-FGRS, Pearson-Aitken Family Genetic Risk Scores; h^2^, narrow-sense heritability; 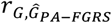, PA-FGRS accuracy; 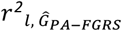, PA-FGRS performance; prev, lifetime prevalence; l, Liability; N_sibe_, Number of full sibling equivalents; Rel., Relatives; Sib., Siblings; Equiv., Equivalents; MZ, Monozygotic; Offsp., Offspring; gen., generation

### PA-FGRS is expected to outperform PGS when GWAS samples are small

Understanding expectations for the realized performance of PGS is important for predicting if and when phenotype-based instruments may become redundant. This is because PGS can, in theory, achieve full accuracy (i.e., *r*_g,PGS_ =1) and performance (i.e., 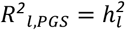) if incorporating all causative variants and errorless effect sizes, but, in practice, limited or biased training GWAS introduce noise. We adapt published frameworks^29,30^ (Online Methods) to describe the expected performance of PGS trained on samples taken from the same population (Figure 3). We emphasize a set of plausible parameters, PGS tagging the human common variant genome (i.e., capturing m=60,000 independent genetic factors^6,31^) and accounting for half of the narrow sense heritability (i.e., 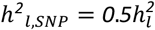), and describe alternate parameters as supplementary analysis (Supplementary Figures S22-24). We then select the sample size needed for PGS to match the performance of PA-FGRS with N_sibe_=1, 3, or 5 (Online Methods; Supplementary Information S7-8; Figure 3). The performance of PA-FGRS with N_sibe_=1 should equal a PGS with effects taken from GWAS with N=65,000 samples from the same population (range of sample size estimates across simulated traits: 61,546 to 71,356; Figure 3A-D, Supplementary Table S5). The performance of PA-FGRS with N_sibe_=3 and N_sibe_=5 hould equal a PGS trained using GWAS with N=190,000 (range: 176,047 to 214,070) and N=300,000 (range: 280,368 to 356,784), respectively (Figure 3A-D, Supplementary Table S5). Parameters defining trait architecture (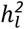 and prevalence) have smaller impacts on these relationships than parameters defining the architecture of the PGS instrument (e.g., m, the number of independent genetic factors covered, h^2^_l,SNP_/h^2^, the proportion of heritability captured by the markers, and p, an effective polygenicity parameter; Supplementary Figures S22-S24; Supplementary Table S5). For rarer traits, maximally powered case-control sampling (c.c.; 50% cases, 50% controls) can increase performance of PGS substantially relative to PA-FGRS (e.g., Figure 3A), but total population size remains a limiting factor. For more common traits, the difference between the PGS and PA-FGRS is less, even under case-control sampling (e.g., Figure 3D). Minimal (N_sibe_=1), realistic (N_sibe_=3), and optimistic (N_sibe_=5) PA-FGRS can outperform PGS when training GWAS have modest sample sizes or the architecture of the PGS is unfavorable, with the relative performance of PA-FGRS being larger for more common traits.

**Figure 3.**
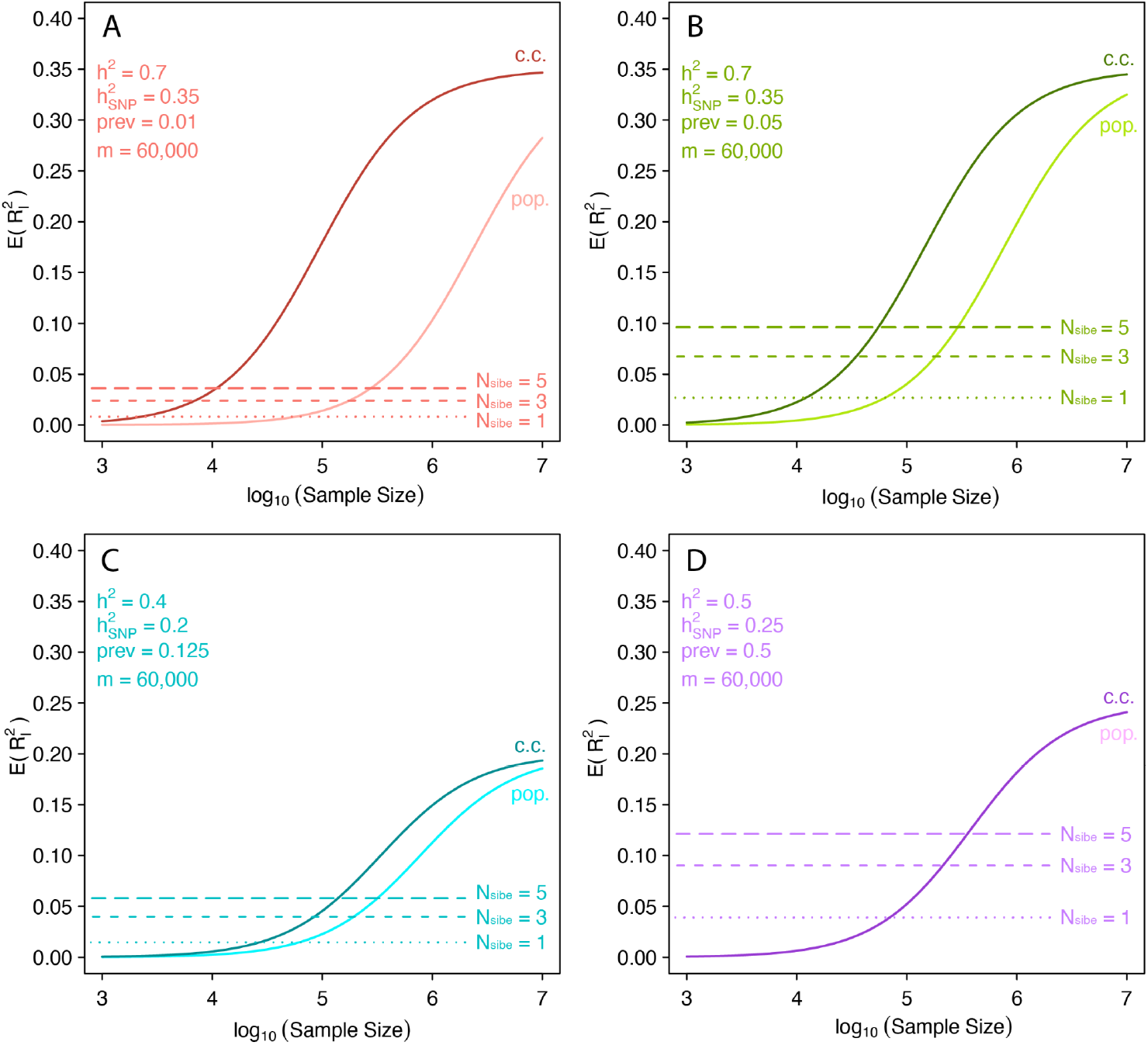
PA-FGRS are expected to outperform PGS unless training GWAS are large. The expected performance of PGS varies as a function of 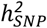, prevalence, and training GWAS sample size (A,B,C,D) in scenarios that assume 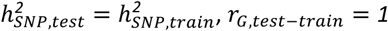, the number of markers in the PGS is m=60,000, and the effective proportion of causal markers is p=1. The expected performance of PA-FGRS with N_sibe_=1,3, or 5 (dashed lines) is not dependent on a training sample and can outperform PGS when training samples are small or of poor quality. The training GWAS sample sizes needed to match the performance of PA-FGRS with N_sibe_=1,3, or 5 are qualitatively similar across traits (x-axis value where solid and dashed lines intersect), but PA-FGRS do relatively better for common traits (dashed lines vs. 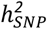 in A,B vs C,D). PA-FGRS, Pearson-Aitken Family Genetic Risk Scores; PGS, polygenic score; h^2^, narrow-sense heritability; *h*^*2*^, SNP heritability of disorder in test/train sample; *r*_*G,test* −*train*_, genetic correlation between disorder in training and test sample; prev, lifetime prevalence; N_sibe_, number of full sibling equivalents; c.c., case-control sampling; pop., population sampling.

### PA-FGRS are expected to complement PGS for at least the near future

The expected correlation and joint performance of the two instruments can be modeled from the previous expectations. Under a simple polygenic liability model (Online Methods), PGS and PA-FGRS can be viewed as estimators of the same latent additive genetic value, *G*, and from this, the expected correlation, as well as joint and conditional performances, can be written as simple functions of their accuracies (Online Methods, Eqs 15-17; Figure 4, Supplementary Figure S24, Supplementary Information S9). When the accuracy of PGS and PA-FGRS are modest, the correlation between the instruments is expected to be modest, even in generative models with no contribution from shared environment (Figure 4A-D). The consistently low accuracy of PA-FGRS across considered scenarios implies consistency modest correlations with PGS (Supplementary Figures S26-S28). Modest correlations between PGS and PA-FGRS imply that, even as PGS training GWAS sample sizes grow, PA-FGRS should remain complementary to PGS (Figures 4E-H). This trend is stronger when trait prevalence is higher (e.g., Figure 4E vs Figure 4F) and when the PGS is limited due to, e.g., small training GWAS or large contributions from non-tagged variants (i.e., when h^2^_SNP_/h^2^ is low or m is high; Figure 4, Supplementary Figures S29-31). Simulating shared environmental contributions from siblings and parents demonstrates that confounded familial and genetic variance in PA-FGRS reduces its empirical correlation with PGS relative to the expectations we derive (Supplementary Figures S18-S21). Taken together, low accuracies of PGS and PA-FGRS imply low to modest correlations between genotype- and phenotype-based genetic instruments are expected, even under a simple polygenic liability threshold model. This trend should persist even as GWAS sample sizes grow.

**Figure 4.**
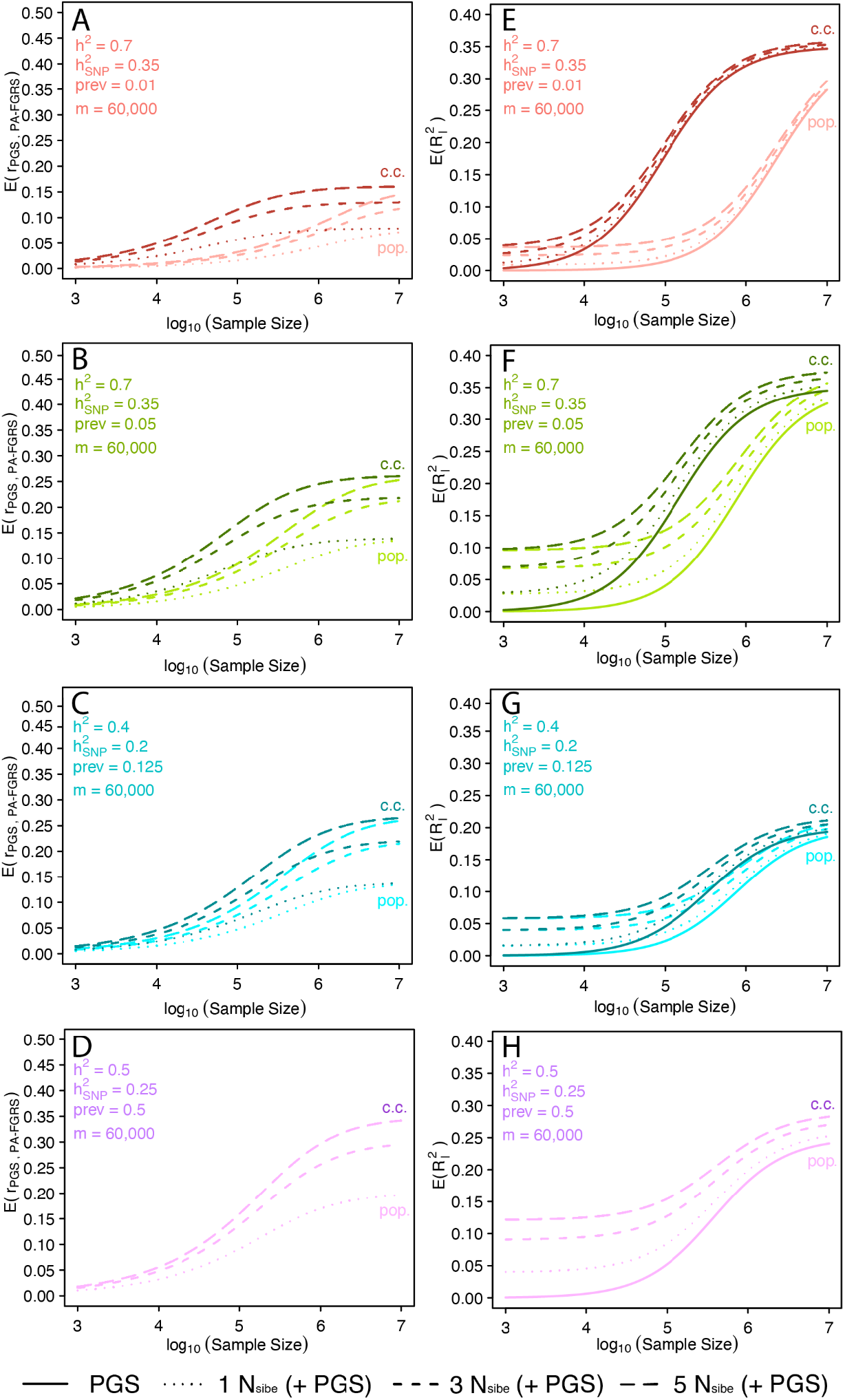
PA-FGRS are expected to complement PGS at current sample sizes and in the near future. The expected correlation between PGS and PA-FGRS (A-D) and the expected joint performance of PGS and PA-FGRS (E-H) vary as a function of 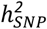, prevalence, and training GWAS sample size in scenarios that assume 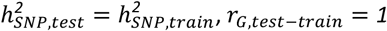, the number of markers in the PGS is m=60,000, and the effective proportion of causal markers is p=1.. The expected correlation between the two instruments (A-D) is expected to remain modest, even as PGS approach maximum performance, especially for rarer traits (A,B). Such modest correlations imply PA-FGRS are expected to remain complementary to PGS in joint models (E-H, dashed lines) explaining independent variance in liability over and above PGS alone (E-H, solid lines). This trend is larger when PGS are trained using smaller GWAS and when traits are more common where higher expected correlations are offset by the higher expected performance of PA-FGRS (EF vs. GH). PA-FGRS, Pearson-Aitken Family Genetic Risk Scores; PGS, polygenic score; 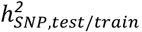, SNP heritability of disorder in test/train sample; *r*_*G,test* −*train*_, genetic correlation between disorder in training and test sample; prev, lifetime prevalence; c.c., case-control sampling; pop., population sampling.

### Expected and observed performance of genetic instruments are in alignment

Finally, we compared the empirical performance of our instruments (Figure 1) to our expectations under a simple polygenic liability model and given our particular input data (Online Methods; Figure 5; Supplementary Tables S5,S6). The iPSYCH proband pedigrees contain on average ∼22 relatives, but many are distant relatives and age-censored such that they correspond to average N_sibe_ of 1.43, 1.16, 2.11, 1.28, 1.70, when predicting ADHD, ASD, BPD, MDD, and SCZ, respectively (Online Methods; Supplementary Table S7). The modest empirical performance of PA-FGRS in our data (Figure 1AB) is broadly consistent with expectations for our theory, with the exception of SCZ which underperforms (Figure 5A). Similarly, the empirical performances of PGS in our data (Figure 1A,B) are broadly consistent with expectations, with the exception of ADHD which overperforms (Figure 5B, Online Methods, Supplementary Table S6). The modest empirical correlations between PGS and FGRS (Figure 1E,F) are, in fact, larger than expected from individual performances (Figure 5C). Our PA-FGRS (Observed N_sibe_, 1.14 to 2.14, Supplementary Table S1) perform empirically as well as we expect PGS trained on population samples of 190,000 to 1,140,000 individuals from the same target population to perform (Figure 5D, Supplementary Table S7). Our PGS (Observed N_Pop. GWAS_, 100,000 to 2,100,000, Supplementary Table S1) perform empirically as well as we expect PA-FGRS constructed from pedigrees with N_sibe_ from 0.3 to 4.7 (Figure 5E, Supplementary Table S7). In contrast with the expectations above, we observe PGS to require many more than 65,000 training samples to achieve the performance of PA-FGRS with N_sibe_=1. We attribute this to differences in simulated versus empirical parameters, namely, in practice, h^2^_SNP_/h^2^ trend much less than 0.5 and genetic correlations between training and target populations trend less than 1 (Supplementary Table S6). These results do not suggest large confounding by shared environment, which would predict higher than expected performance of PA-FGRS and lower than expected correlation between PA-FGRS and PGS (Supplementary Figures S18-21). As such, low empirical correlations can be consistent with simple polygenic liability models of inheritance and PA-FGS with only a few relatives can be as useful for predicting disease, in practice, as PGS constructed using large GWAS.

**Figure 5.**
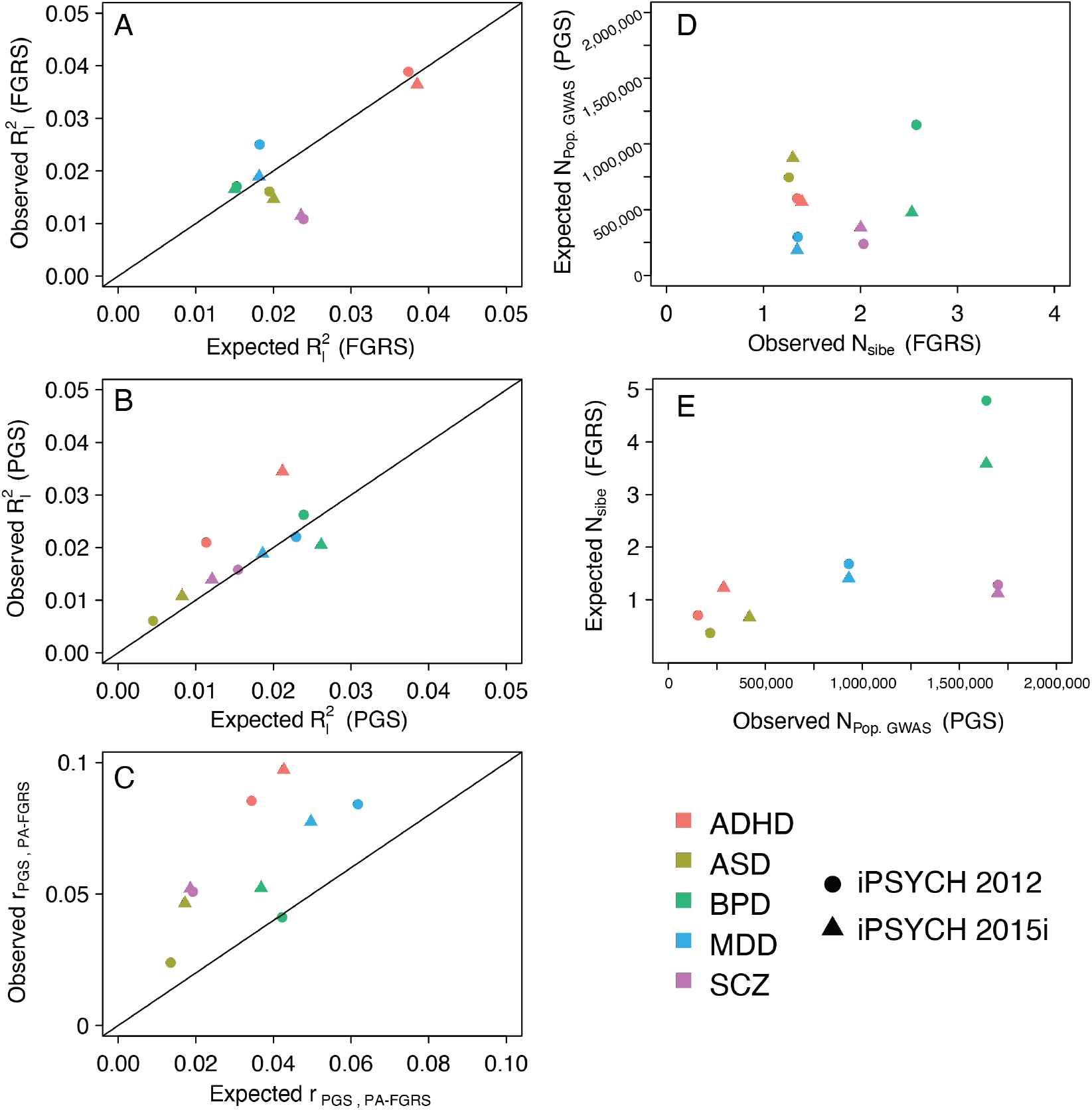
Observed performances of genetic instruments are generally in alignment with expectations assuming simple polygenic liability models. (A) The observed performance of PA-FGRS is broadly consistent with expectations of a simple additive polygenic model and parameters describing our real data (*N*_*sibe*_, 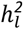, prev., etc.). (B) The observed performance of PGS is similarly broadly consistent with expectations given estimated 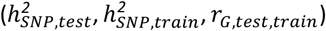 and assumed (prev., m=60,000, p=1) parameters. (C) The observed correlations between PGS and PA-FGRS are modest and larger than predicted by marginal performance of the instruments. (D) The empirical performance of the 10 PA-FGRS (x-axis, with *N*_*sibe*_ ≈ 1.14 to 2.14) is equivalent to that expected from PGS trained in very large samples (y-axis, N ≈ 190,000 to 1,140,000; assuming 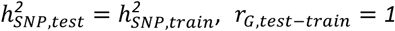). (E) The empirical performance of the 10 PGS (x-axis, with *N*_*Pop*.*GW AS Equiv*_ ≈ 150,000 to 2,100,000) is equivalent to that expected from PA-FGRS with *N*_*sibe*_ ≈ 0.3 to 4.7. PA-FGRS, Pearson-Aitken family genetic risk scores; PGS, polygenic score; Liab., liability; Sib., sibling; Equiv., Equivalence; ADHD, Attention deficit hyperactivity disorder; ASD, autism spectrum disorder; BPD, bipolar disorder; MDD, major depressive disorder; SCZ, schizophrenia; 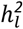, narrow-sense heritability; 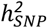, SNP heritability; prev., lifetime prevalence; *r*_*G*_, genetic correlation; *N*_*sibe*_, Number of siblings equivalent to pedigree; *N*_*Pop*.*GW AS*_, equivalent N assuming a population sampling GWAS.

## Discussion

We demonstrate and replicate that phenotype-based genetic instruments (PA-FGRS, FH) and genotype-based genetic instruments (PGS) explain comparable, nearly independent, proportions of disease liability to five psychiatric disorders. This trend - that family phenotypes and genotypes are quite modestly correlated in models of human disease - is reflected in a number of contemporary studies^13–17,21^, but the reason for this behavior has been unknown. We explored this phenomenon, concluding that PA-FGRS, FH, and PGS behave consistent with noisy estimators of a highly similar underlying genetic liability. We believe that even as GWAS grow to sample sizes in the millions, phenotype-based genetic instruments will remain complementary. The current emphasis on molecular approaches in human genetics should not discard the continued potential for familial phenotypic records in genetic medicine and genetic epidemiology.

Phenotype and genotype-based instruments have been compared, contrasted, and combined much more extensively in agricultural sciences^3,5–7^. In such applications, genomic predictions are now the preferred method for assessing breeding potential, attributable to the genetic architectures of most agricultural species, which are very different from those underlying trait variation in human populations. Decades of controlled breeding have greatly reduced genetic diversity and simplified the genetic architecture of studied traits^6^. Pedigrees also have different densities of different relatives, all of whom may have available molecular genotypes^6^. A key parameter in determining the performance of a PGS is the number of independent genetic elements segregating among genomes of the target population^29,30^, which is a function of genetic diversity. For dairy cattle this number may be ∼5,000, but for humans it may range from ∼60,000 (the genome captured by SNP-arrays) to ∼500,000 (the genome captured by whole genome sequencing)^6^. This, along with a focus on binary disease outcomes, limits the pace at which PGS could make phenotypic records redundant in human biomedical applications.

Our proposal, that genomic prediction is a hard problem in human biomedicine and genetic instruments are simply very noisy, is in accordance with the preponderance of evidence from quantitative genetics^25,26^. Twin and family studies repeatedly purport a large additive genetic contribution to complex disorder etiology, with sparser, less consistent, and smaller contributions from shared (i.e., familial) environment. Among the five psychiatric disorders we studied, the contributions from shared environment may be largest for major depressive disorder. However, for MDD, the most direct and powerful tests for familial environment, comparing half-siblings reared together and apart^32^ or children and adoptive/step-parents^33^, and found significant, but very modest effects of shared environment. For some specific traits or disorders, e.g., substance use^34–36^ or educational attainment^37^, substantial shared environmental effects have been reported, but these appear to be exceptions rather than a rule^26^. Still, we can not interpret our results as suggesting effects of shared environment are always zero. It is, however, more likely, parsimonious, and consistent with a broader body of work to ascribe currently reported modest correlations between genotype- and phenotype-based instruments to error in measuring highly similar additive genetic liability.

At the moment, PGS outperform PA-FGRS for SCZ and BPD, but not for ADHD and ASD. For SCZ and BPD, this can be attributed to two things: the large training GWAS for BPD^38,39^ and SCZ^38,39^ with population sample sizes >1.5 million and the rarity and later onset of SCZ and BPD limiting PA-FGRS. In comparison, for ADHD and ASD, PGS are trained in much smaller GWAS (population sample sizes: 150,000 to 400,000) and PA-FGRS use more complete records as onset is younger and, at least for ADHD, prevalence is higher. However, we observe that our ADHD PGS outperforms BPD and SCZ PGS, despite the latter having much larger training samples. Here, being trained on a highly comparable sample (train-test sample genetic correlations: ∼1 for ADHD vs. ∼0.45 for SCZ and ∼0.78 for BPD) overcomes differences in training GWAS of >1,000,000 population samples. This illustrates that the best single genetic predictor will depend on disease parameters (h^2^ and prevalence), the usefulness of familial records, and, critically for PGS^40^, the genetic similarity between the training and target samples. The various instruments, then, have unique sources of error, further supporting integration of multiple predictors and expectations of modest overlap. Differences in, at least, genetic ancestry^41^, ascertainment schema^42^, and phenotype quality^43^ are known to reduce genetic correlation among test-training pairs, and will continue to affect PGS performance if not given attention. The within family nature of PA-FGRS mitigates some of this loss of efficiency. In scenarios where financial resources, trait (genetic) architecture, or obtainable/comparable population sizes are limiting, opting to collect detailed familial records may remain a more efficient approach for building best-in-practice genetic instruments.

Our work informs future research using genetic instruments. First, we found that widely implemented approaches for estimating variance explained on the liability scale^27^ can be biased for PA-FGRS or FH. We propose directly estimating liability scale variance using appropriately weighted probit models in order to protect against reporting strongly biased estimates. Second, the concept of heritability has a central place in generative and etiological models, but these models are abstract and describe asymptotic (i.e., infinite sample size) performance of potentially practical predictive instruments. As we have shown, for human biomedical research, genetic predictions based on phenotypes of relatives will *never* account for the full heritability, in practice - it would require *at least* hundreds to thousands of half-sibling offspring records per proband. Whole genome sequence based PGS, which may need to capture ∼500,000 independent genomic factors^6^, could require training samples equal to, or beyond, the current global population to match performance to heritability for some diseases. Such extreme sample sizes are, in part, a function of lost power when converting a continuous liability score to a binary diagnosis^44^ and may not be required for quantitative traits, such as human height^45^. There are and will continue to be methodological advances that improve efficiency of PGS beyond our simple modeling framework, but it is worth discussing if heritability is the right goal post for *practical* aims of human biomedicine, such as prediction or classification. Third, if our interpretation of PGS, PA-FGRS, and FH as highly noisy estimates of the same genetic liability is correct, covarying or adjusting, e.g., FH by PGS, will not provide adequate control for confounding of the underlying genetic liability construct. Models that consider or make adjustments for the expected amount of noise in an instrument^46^, a quantity we show can be estimated with plausible accuracy, could be preferred.

Our study should be viewed in light of some limitations. First, our familial records are incomplete but still may overestimate effect sizes of predictions in clinical settings. Our modeling leverages all familial information available at the end of follow-up, which is valid for etiological models, but reflects information that may not be available at the time of a clinical decision (e.g., birth or first hospital contact, etc.). Imperfect patient recall or age-censoring in relatives regarding diagnoses made subsequent to the time of evaluation of the proband would reduce the information available. Our results may not generalize to predictive studies which should carefully account for such “conditioning on the future” to better mimic real clinical assessments or risk-screens. Second, we found that the modest correlation observed between PA-FGRS and PGS was actually *higher* than our simple framework predicts, indicating that our model does not reflect the generative model underlying our real data. This could be due to a number of features we do not model, such as non-random mating, indirect genetic effects, frequency dependent genetic architectures, or other phenomena that we have not considered, but the trend is nevertheless, inconsistent with confounding by shared environment. Third, and related, PA-FGRS is derived under a simple, additive liability model - the predominant model for dichotomous traits in genetics for nearly a century - but this currently limits our ability to consider these more nuanced contributions.

Genetics as a field is rooted in resemblance among relatives and even as molecular techniques provide increasingly deep and specific access to variability across the genomes of millions of individuals, incorporating these foundational sources of data is expected to play a continued, critical role in describing genetic liability for complex traits and disorders.

## Online Methods

### iPSYCH2015 Case-Cohort Study

The Lundbeck Foundation initiative for Integrative Psychiatric Research (iPSYCH)^47,48^ samples singleton births between 1981 and 2008, alive on their first birthday, with known mothers residing in Denmark (N=1,657,449). The iPSYCH 2012 case-cohort includes 86,189 individuals (30,000 random population controls; 57,377 psychiatric cases)^47^, while the iPSYCH 2015i case-cohort includes an independent 56,233 individuals (19,982 random population controls; 36,741 psychiatric cases)^47,48^. Taken together, these two ascertainments comprise the iPSYCH 2015 case-cohort study. DNA was taken from dried blood spots housed at the Danish Neonatal Screening Biobank^49^. The Infinium PsychChip v1.0 array (2012) or the Global Screening Array v2 (2015i) were used for genotyping. Register based diagnoses are made by licensed practitioners during in- or out-patient specialty care (diagnoses or treatments assigned in primary care are not included) and are obtained from the Danish Psychiatric Central Research Register (PCRR)^50^ and the Danish National Patient Register (DNPR)^51^. Unique identifiers of the Danish Civil Registration System^52^ enables linkage across population registers, to parental identifiers, and to the neonatal blood spots. The use of this data follows standards of the Danish Scientific Ethics Committee, the Danish Health Data Authority, the Danish Data Protection Agency, and the Danish Neonatal Screening Biobank Steering Committee. Data access was via secure portals in accordance with Danish data protection guidelines set by the Danish Data Protection Agency, the Danish Health Data Authority, and Statistics Denmark.

Imputation and quality control of genotypes followed custom, mirrored protocols^53^

BEAGLEv5.1^54,55^ was used to phase and impute the data in conjunction with reference haplotypes from the Haplotype Reference Consortium v1.1 (HRC)^56^. SNPs were checked for missing data across SNPs and individuals, deviations from Hardy-Weinberg equilibrium in controls, abnormal heterozygosity, minor allele frequency (MAF), batch artifacts, and imputation quality. 7,649,999 imputed allele dosages were retained for analysis. Samples were checked for genotype-phenotype sex discordance and abnormal heterozygosity. KING^57^ was used to estimate kinship and ensure no second degree or higher relatives remained within either cohort or across cohorts. The smartpca module of EIGENSOFT^58^ was used to estimate PCs and a classifier was used to remove individuals distant from those with parents and four grandparents born in Denmark. For this study, in particular, we defined the following disorders according to ICD-10 codes associated with at least one registered treatment: major depressive disorder (MDD; F32-3), bipolar disorder (BPD; F31), schizophrenia (SCZ; F20), autism spectrum disorders (ASD; F84, F84.1, F84.5, or F84.9), and attention deficit hyperactivity disorder (ADHD; F90.0). In total, we retain, in iPSYCH 2012 24,266 random probands, 14,970 ADHD probands, 12,101 ASD probands, 1581 BPD probands, 18238 MDD probands, 2624 SCZ probands, and in iPSYCH 2015i 15,381 random probands, 7499 ADHD probands, 5600 ASD probands, 1141 BPD probands, 8668 MDD probands, 2721 SCZ probands.

### iPSYCH 2015 case-cohort genealogies

Details of the iPSYCH 2015 case-cohort genealogy are described previously^21^ following previous approaches^59^. Briefly, 2,066,657 unique relatives were obtained using mother-father-offspring linkages for the 141,265^60^ (before QC) genotyped probands. A population graph was constructed using *kinship2*^*61*^ and *FamAgg*^*60*^ packages for R with edges defined by recorded trios. Relatedness coefficients were estimated by summing unique paths between two individuals, weighted by (0.5)^(number of edges in the path)^62^. Same-sex twins were given kinship of 0.375 and maternal siblings with missing paternal records were given kinship of 0.25 according to prior comparisons with SNP-data^21^.

Diagnoses made between 1968 and 1994 are limited to in-patient contacts and ICD-8 codes and were assigned to disorder labels to match ICD-10 groups as follows: major depressive disorder (MDD; 296.09, 296.29, 298.09, or 300.49, but not 296.19, 296.39, or 298.19), bipolar disorder (BPD; 296.19, 296.39, or 298.19), schizophrenia (SCZ; 295.x9, excluding 295.79), autism spectrum disorders (ASD; 299.01-299.04), and attention deficit hyperactivity disorder (ADHD; 308.01)^47^. FH variables were defined as having at least one affected parent, half sibling, or full sibling.

Narrow sense heritability (h^2^) for each psychiatric disorder was estimated using relatives in the genealogies of the random population sample. To account for cohort effects, we selected 250,057 sibling pairs in these genealogies such that both siblings were born in the same decade (either, 1960-1970, 1970-1980, 1980-1990, 1990-2000, 2000-2010). We then estimated the sib-pair tetrachoric correlation for each disorder within each decade, combining the estimates in an inverse variance weighted meta-analysis. The meta-analyzed tetrachoric correlation and its standard deviation were divided by the sibling kinship (0.5) to give an estimate of h^2^ and compute 95% confidence intervals.

### A simple, additive polygenic liability threshold model

Our approach is based on a simple additive polygenic liability threshold model, which is tractable, useful, and commonly employed. Briefly, We assume that for a fully observed individual *i* their binary disease status, *D*_*i*_, is the result of a latent, normally distributed liability, *L*_*i*_. *D*_*i*_ is expressed when *L*_*i*_ is above a critical threshold, *t*, that is the upper tail standard normal quantile corresponding to the lifetime prevalence of the disease, *K*_pop_.

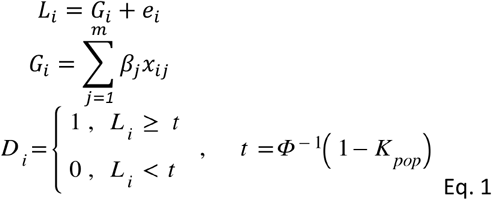

The liability *L*_*i*_ is the sum of a genetic value, *G*_*i*_, and an environmental deviation, *e*_*i*_, and we assume no G-E interactions and no contributions from shared environment. *G* is an additive function of genetic factors, *x*, with true allelic average effects, *β*, and explains h^2^ proportion of population variance in *L*. The genetic value may itself have two components, a GWAS related SNP component, *G*_*i,SNP*_, explaining h^2^_SNP_ proportion of variance in liability, and genetic residual, explaining h^2^ - h^2^_SNP_ that may represent incomplete linkage or other genetic factors.

### Computing Pearson-Aitken Family Genetic Risk Scores (PA-FGRS)

PA-FGRS^21^ estimates the genetic component of latent disease liability under assumptions of a modified polygenic liability threshold model that relaxes the assumption above that individuals are fully observed by defining

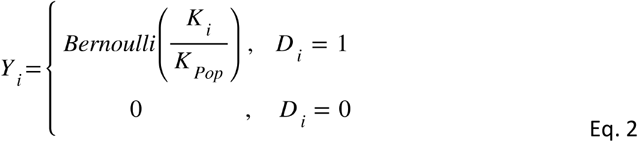

where *K*_*i*_ is the cumulative incidence of the disease in the age window during which individual *i* was observed. The key difference from above is that here, the probability of a true disease status *D*_*i*_ being observed (i.e., that *Y*_*i*_ = *1* | *D*_*i*_ = 1) is related to the proportion of the lifetime incidence for which an individual was observed. As a result, we model the expected liabilities of cases and controls as *mixtures* of truncated normals such that observed controls are treated as a probabilistic mixture of a true case and a true control.

PA-FGRS estimates the genetic component of latent disease liability under this model for a proband, *p*, from the observed disease statues of their *n*_*rel*_ relatives, 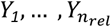 a modified relatedness matrix, *Σ*, describing the relationship between trait liability in relatives and genetic liability of the proband, and known parameters, *θ*, such as the age of each relative at the end of follow up, and the age-specific incidence of the trait.

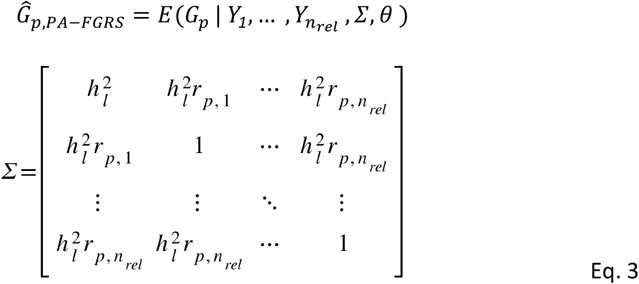

where *r*_*p,i*_ is the relatedness coefficient between the proband and their i-th relative. Briefly, we estimate *Ĝ*_*p,PA-FGRS*_ according to the Pearson-Aitken (PA) selection formula to provide a direct, efficient, and highly accurate solution for an estimator that otherwise requires expensive resampling or numerical integration techniques ^21^.

The PA updating proceeds as follows. First, the vector of mean expected liabilities for all relatives, *μ*, and the expected covariance among liabilities of relatives, *Ω*, are initialized to expectations in the population assuming no observations have been made: *μ*_*0*_ = *0* and *Ω*_*0*_ = *Σ*. Then, *μ* and *Ω* are updated iteratively, conditioning on each relative *x* = *1*,… *n*_*rel*_ in sequence. At each step, the current expected liability of the selected relative, *μ*_*x*_, is updated to a conditional expectation, 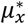, based on the current estimate, the observed disease status of the relative, and other known parameters.

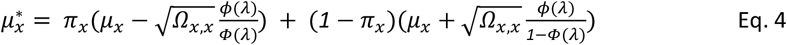

with *ϕ*(⋅), the PDF of the standard normal distribution, Ф(⋅), the CDF of the standard normal distribution, 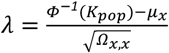, and 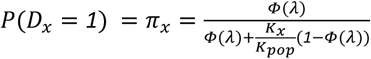 if *Y*_*x*_ = *0* or *π*_*x*_ = *1* if *Y*_*x*_ = 1. All remaining (i.e., unselected) liabilities are then updated conditional on the updated liability of the selected relative.

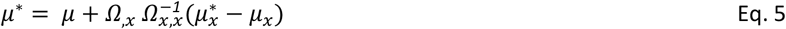

This can be thought of as propagating a non-linear (i.e., liability threshold model appropriate) weighting of the observed disease status of the selected relative into the expected liabilities of their genetic relatives, including the proband.

Similarly, the liability covariance, *Ω*, is updated at each step by first updating the variance of the mean expected liability for the selected relative, and then the rest of the covariance conditional on this updated variance for the selected relative.

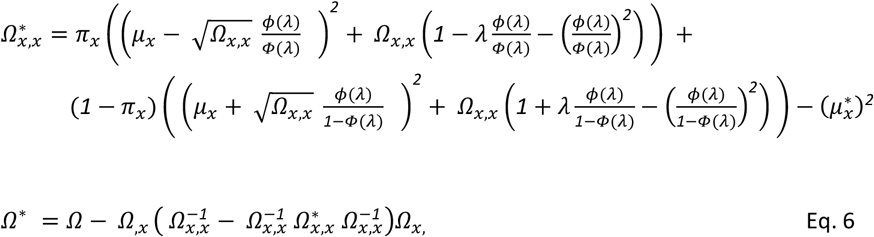

The procedure is repeated until only the proband remains unselected (i.e., proband liability has been conditioned on all relatives). For further details see Krebs et al ^21^ and Supplementary Information. We computed these PA-FGRS for individuals from the iPSYCH 2012 and iPSYCH 2015i cohorts separately (Supplementary Table S1) using trait parameters described in Supplementary Table S6.

### Computing polygenic scores (PGS)

PGS approximate the genetic value under the polygenic liability threshold model by substituting a set of observed SNP marker genotypes 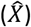 for an individual, i, for the unknown generative genetic factors (X), and using transformations of per allele effect estimates from a training GWAS 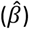 as a surrogate for generative allelic effects (*β*).

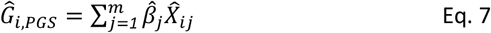

We computed PGS combining imputed dosages with 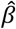 estimated using SBayesR^63^ to update results provided by large GWAS. For SBayesR we followed the developer recommendations using provided UKBiobank LD matrix estimated for 2.8 million SNPs and with parameters: gamma=0.0,0.01,01,1, pi=0.95,0.02,0.02,0.01, burn-in=5000, chain-length=25000, exclude-mhc, impute-n. For BPD ^38^, MDD ^64^, and SCZ ^39^ we use large published GWAS that are independent of iPSYCH. For ADHD and ASD we use the results of mixed effects model GWAS^65^ from the complementary iPSYCH cohort (e.g., beta from GWAS in iPSYCH 2015i for PGS in iPSYCH 2012).

### Statistical model fitting to iPSYCH data

Variance in liability explained by combinations of genetic instruments was estimated with weighted multivariate generalized linear models incorporating a probit link function. A baseline model included age, gender, and the first ten principal components of an estimated genetic relatedness matrix with subsequent models including combinations of PGS, FH, and PA-FGRS. The liability scale variance for each model was estimated according to a modified approach presented in Lee et al ^27^ as,

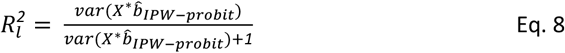

where 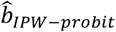 is a vector of coefficients from the inverse probability weighted probit regression of the disease status on the matrix of covariates *X*. For the weighting, two-stage sampling weights were computed as the product of the case-control sampling weights suggest by Lee et al ^27^ (unit weights in controls and 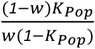 in cases) and a second, study specific, probability weight to reflect selection out of the sample due to relatedness pruning (Supplementary Note S1). *X*^∗^, a random subset of *X* such that *w* ∗= *K*_*Pop*_.

Reported liability variance explained is after subtracting that of a baseline covariates only model. Empirical p-values were used to test for significance of changes in liability variance explained in nested models as the proportion of times out of 100,000 that the empirical change associated with adding a variable to a model was larger than adding a random permutation of the variable. Significance adjusted to p < 0.05/110 model contrasts = 4.55 × 10-4. Unadjusted and adjusted odds ratios (OR) were estimated using multivariate generalized linear models with a logit link function. Pearson’s correlations among genetic instruments were estimated in the random subcohort of each iPSYCH cohort and not adjusted for covariates.

### Estimating the expected Accuracy of Pearson-Aitken Family Genetic Risk Scores (PA-FGRS)

#### Exact Solution

The expected accuracy of our PA-FGRS estimator was derived fully in the Supplementary Information S11 and is defined as the product of the inverse of the heritability and the variance of the unbiased estimator *Ĝ*_*PA-FGRS*_,

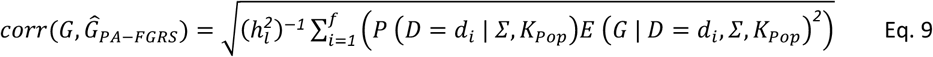

where D is *n*_*rel*_-variate thresholded gaussian, and the conditional expectation of G is the PA estimator above. Where {*d*_*1*_,*d*_*2*_,…,*d*_*f*_} is the set of 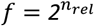 possible configurations of disease in the selected pedigree. For pedigrees with relatively few numbers of relatives we can solve this by a explicit calculation of the n_*rel*_-variate integral over the *f* configurations (see code availability for our function *fgrs_accuracy(…, method=”pa”,estimate= “theory”)*), but for larger pedigrees it is intractable. This full solution is used for the FGRS expectation results presented in Figures 2-4.

#### Linearly Approximated Solution

Alternatively, one may use more tractable expectations of accuracy from a linear estimator (i.e., expected breeding value) in place of the full liability scale estimator PA-FGRS (Supplementary Information S3-5), similarly defined as the product of the inverse of the heritability and the variance of *Ĝ*_*linear*_,

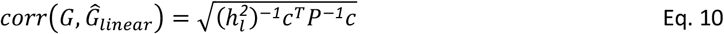

with *c* being a 1 x n_rel_ vector describing the expected covariance between the proband genetic liability and the true disease status of the i-th relative such that 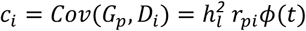. **P** is an n_rel_ x n_rel_ matrix describing the expected covariance in disease status among the relatives, which can be approximated such that 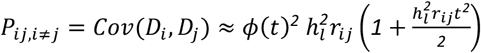 and *P*_*ij,i*=*j*_ = *Var(D*_*i*_) = *K*_*pop*_*(1* − *K*_*pop*_) following the second order Taylor approximation used by Golan et al^66^. Here, r_*pi*_ is the relatedness coefficient between the proband, p, and their i-th relative, *r*_*ij*_ is relatedness coefficient between the i-th and j-th relatives, and *t* is the liability threshold value, *t* = Ф^−*1*^(*1* – *K*_*Pop*_). We show that this is a good approximation of the full solution in smaller pedigrees (Supplementary Figures S14-S16) and use this to compute the expected accuracy of PA-FGRS for iPSYCH pedigrees as presented in Figure 5. When individuals are not fully observed, we use 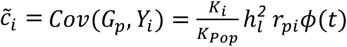 and 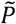 where and 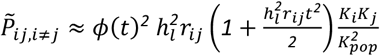 to estimate this accuracy as,

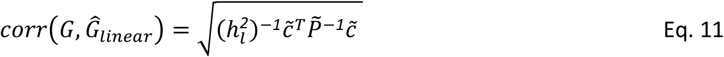

#### Linearly Approximated Number of Full Sibling Equivalents (*N*_*sibe*_)

In the special case of pedigree containing one relative type (e.g., Full siblings of the proband), the second order Taylor approximation of Eq. 10 reduces to,

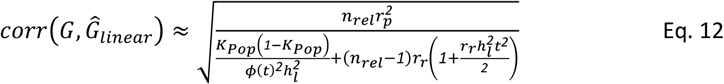

Here, *r*_*p*_ is the relatedness coefficient between the relatives and the proband, *r*_*r*_ is the relatedness coefficient among the relatives which may be different than *r*_*p*_ (e.g., as for half-sibling offspring where *r*_*p*_=0.5, *r*_*r*_=0.25). To derive the *N*_*sibe*_ for any predictor, we set this equation to its observed or expected accuracy, which for an empirical predictor can be recovered its performance as 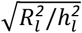, and solve this equation for the value of *n*_*rel*_ when *r*_*p*_ = *r*_*r*_ = *0*.*5*. This approximation was used to estimate *N*_*sibe*_ for iPSYCH pedigrees, simulated pedigrees, and to equate observed PGS sample size to equivalent PA-FGRS *N*_*sibe*_.

### Estimating the accuracy of polygenic scores (PGS)

The expected accuracy of polygenic scores has been described elsewhere by, e.g., Wu et al ^30^, Daetwyleer et al ^29^, and Dudbrige ^67^ under related, but not identical assumptions (see Supplementary Information S7-8). We use the formulas from Wu et al ^30^ to estimate the expected squared accuracy of a PGS as

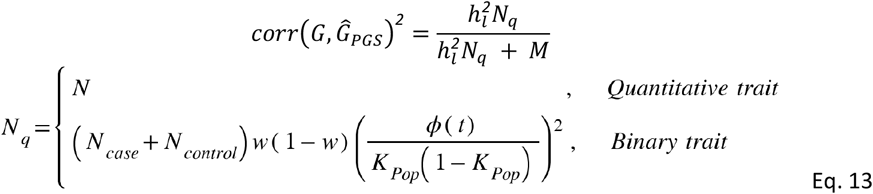

where *M* is the effective number of independent markers. Shrinkage-based estimators can gain power in the context of non-infinitesimal models (e.g., ldpred ^68^) and we can evaluate,

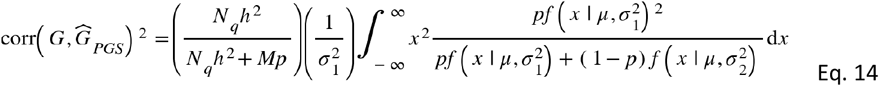

where a non-standardized Gaussian 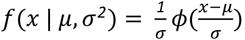, and with 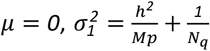, and 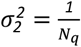

### Expected correlation between PA-FGRS and PGS

Under a simple liability framework as we propose, the *Ĝ*_*PA-FGRS*_ and *Ĝ*_*PGS*_ are statistically independent conditional on the true genetic value (G)^69^ (Supplementary Information S9) In this scenario, the expected correlation is

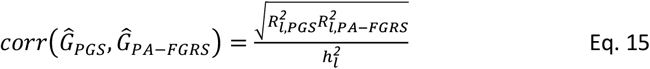

Where 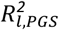 and 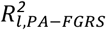 are the liability variance explained by the estimated genetic values and 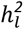 is the liability scale heritability.

### Expected joint performance of PA-FGRS and PGS

The expected semi-partial accuracy in liability of the PA-FGRS, or expected correlation between the PA-FGRS and liability after accounting for the PGS, can be written as,

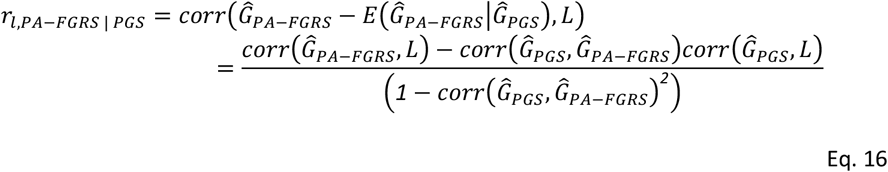

which allows us to predict the expected joint performance of a PGS and PA-FGRS fit in the same model as,

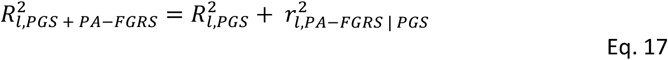

### Comparisons of expected and observed performance

Expected performance and relationships of genetic instruments depends on both assumed and estimated trait parameters that are described in Supplementary Table S6. The expected performance of PA-FGRS in iPYSCH were computed using the mean of the linear approximation for the expected accuracy of PA-FGRS for each iPSYCH pedigree using known values for 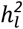 and *K*_*Pop*_.

The expected performance of the PGS in iPSYCH were calculated by first estimating, using the non-shrinkage estimator, the expected squared accuracy of the PGS in the training sample following the expectations of the non-shrinkage estimator above assuming M=60,000, and using training sample parameters N_case_, N_controls_, 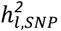, *K*_*Pop*_. The expected out of population test sample performance (i.e., in iPSYCH) was then estimated as,

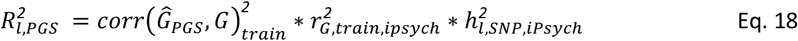

Where 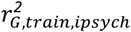 is the squared genetic correlation between the phenotype as defined in the training GWAS and in iPSYCH, and 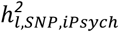 is the SNP-based heritability for the iPSYCH trait.

The expected correlation between the observed PGS and PA-FGRS were estimated as above using provided values for 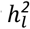.

The population GWAS sample size expected to result in a PGS with performance equal to that of the empirical PA-FGRS instruments, was calculated by setting the non-shrinkage based estimator (Eq. 13) equal to the observed value for squared accuracy of the PA-FGRs, (i.e., 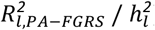) and solving for *N*_*q*_ assuming *w* = *K*_*Pop*_. The calculations assumed the training GWAS was performed in the same population that the PGS was applied within (i.e., 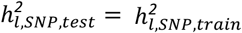 and *r*_*G,train,test*_ = 1), that m=60,000 and known *K*_*Pop*_.

The pedigree *N*_*sibe*_ expected to result in a PA-FGRS with performance equal to that of the empirical PGS instruments, was calculated by setting the linear approximation estimator for full sibling pedigrees above (Eq. 12) equal to the observed value for squared accuracy of the PGS, (i.e., 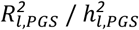) and solving for *n*_*rel*_ assuming *K*_*Pop*_ and 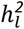.

### Code availability

PA-FGRS, predicted performance, and simulations are implemented in R code available at https://github.com/BioPsyk/PAFGRS

## Data Availability

All data produced in the present study are available upon reasonable request to the authors and in accordance with Danish legislation

## References

1. Falconer, D. S. Introduction to quantitative genetics. (Prentice Hall, 1996).

2. Lynch, M. & Walsh, B. Genetics and analysis of quantitative traits. https://www.invemar.org.co/redcostera1/invemar/docs/RinconLiterario/2011/febrero/AG_8.pdf.

3. Goddard, M. E. & Hayes, B. J. Genomic selection. Journal of Animal Breeding and Genetics vol. 124 323–330 Preprint at https://doi.org/10.1111/j.1439-0388.2007.00702.x (2007).

4. Meuwissen, T., Hayes, B. & Goddard, M. Genomic selection: A paradigm shift in animal breeding. Animal Frontiers vol. 6 6–14 Preprint at https://doi.org/10.2527/af.2016-0002 (2016).

5. Meuwissen, T. H., Hayes, B. J. & Goddard, M. E. Prediction of total genetic value using genome-wide dense marker maps. Genetics 157, 1819–1829 (2001).

6. Wray, N. R., Kemper, K. E., Hayes, B. J., Goddard, M. E. & Visscher, P. M. Complex Trait Prediction from Genome Data: Contrasting EBV in Livestock to PRS in Humans: Genomic Prediction. Genetics 211, 1131–1141 (2019).

7. Hayes, B. J., Lewin, H. A. & Goddard, M. E. The future of livestock breeding: genomic selection for efficiency, reduced emissions intensity, and adaptation. Trends Genet. 29, 206–214 (2013).

8. Wray, N. R., Goddard, M. E. & Visscher, P. M. Prediction of individual genetic risk to disease from genome-wide association studies. Genome Res. 17, 1520–1528 (2007).

9. Torkamani, A., Wineinger, N. E. & Topol, E. J. The personal and clinical utility of polygenic risk scores. Nat. Rev. Genet. 19, 581–590 (2018).

10. Khera, A. V. et al. Genome-wide polygenic scores for common diseases identify individuals with risk equivalent to monogenic mutations. Nat. Genet. 50, 1219–1224 (2018).

11. Schork, A. J., Schork, M. A. & Schork, N. J. Genetic risks and clinical rewards. Nature genetics vol. 50 1210–1211 (2018).

12. Dudbridge, F. Polygenic Epidemiology. Genet. Epidemiol. 40, 268–272 (2016).

13. Agerbo, E. et al. Risk of Early-Onset Depression Associated With Polygenic Liability, Parental Psychiatric History, and Socioeconomic Status. JAMA Psychiatry 78, 387–397 (2021).

14. Schendel, D. et al. Evaluating the interrelations between the autism polygenic score and psychiatric family history in risk for autism. Autism Res. 15, 171–182 (2022).

15. Agerbo, E. et al. Polygenic Risk Score, Parental Socioeconomic Status, Family History of Psychiatric Disorders, and the Risk for Schizophrenia: A Danish Population-Based Study and Meta-analysis. JAMA Psychiatry 72, 635–641 (2015).

16. Hujoel, M. L. A., Loh, P.-R., Neale, B. M. & Price, A. L. Incorporating family history of disease improves polygenic risk scores in diverse populations. Cell Genom 2, (2022).

17. Mars, N. et al. Systematic comparison of family history and polygenic risk across 24 common diseases. bioRxiv (2022) doi:10.1101/2022.07.06.22277333.

18. Kendler, K. S., Ohlsson, H., Bacanu, S., Sundquist, J. & Sundquist, K. Differences in genetic risk score profiles for drug use disorder, major depression, and ADHD as a function of sex, age at onset, recurrence, mode of ascertainment, and treatment. Psychol. Med. 53, 3448–3460 (2023).

19. Kendler, K. S., Ohlsson, H., Sundquist, J. & Sundquist, K. Impact of comorbidity on family genetic risk profiles for psychiatric and substance use disorders: a descriptive analysis. Psychol. Med. 53, 2389–2398 (2023).

20. Kendler, K. S., Ohlsson, H., Sundquist, J. & Sundquist, K. Family Genetic Risk Scores and the Genetic Architecture of Major Affective and Psychotic Disorders in a Swedish National Sample. JAMA Psychiatry 78, 735–743 (2021).

21. Dybdahl Krebs, M. et al. A novel method for estimating familial genetic risk complements whole genome genotyping in improving our understanding of the genetic architecture of major depressive disorder. In Preparation (2022).

22. Musliner, K. L. et al. Polygenic Risk and Progression to Bipolar or Psychotic Disorders Among Individuals Diagnosed With Unipolar Depression in Early Life. Am. J. Psychiatry 177, 936–943 (2020).

23. Musliner, K. L. et al. Polygenic Liability and Recurrence of Depression in Patients With First-Onset Depression Treated in Hospital-Based Settings. JAMA Psychiatry 78, 792–795 (2021).

24. Musliner, K. L. et al. Association of Polygenic Liabilities for Major Depression, Bipolar Disorder, and Schizophrenia With Risk for Depression in the Danish Population. JAMA Psychiatry 76, 516–525 (2019).

25. Hill, W. G., Goddard, M. E. & Visscher, P. M. Data and theory point to mainly additive genetic variance for complex traits. PLoS Genet. 4, e1000008 (2008).

26. Polderman, T. J. C. et al. Meta-analysis of the heritability of human traits based on fifty years of twin studies. Nat. Genet. 47, 702–709 (2015).

27. Lee, S. H., Goddard, M. E., Wray, N. R. & Visscher, P. M. A better coefficient of determination for genetic profile analysis. Genet. Epidemiol. 36, 214–224 (2012).

28. Mrode, R. A. & Thompson, R. Linear Models for the Prediction of Animal Breeding Values. (CABI, 2014).

29. Daetwyler, H. D., Villanueva, B. & Woolliams, J. A. Accuracy of predicting the genetic risk of disease using a genome-wide approach. PLoS One 3, e3395 (2008).

30. Wu, T., Liu, Z., Mak, T. S. H. & Sham, P. C. Polygenic power calculator: Statistical power and polygenic prediction accuracy of genome-wide association studies of complex traits. Front. Genet. 13, 989639 (2022).

31. Wray, N. R. et al. Pitfalls of predicting complex traits from SNPs. Nat. Rev. Genet. 14, 507–515 (2013).

32. Kendler, K. S., Ohlsson, H., Lichtenstein, P., Sundquist, J. & Sundquist, K. The Genetic Epidemiology of Treated Major Depression in Sweden. Am. J. Psychiatry 175, 1137–1144 (2018).

33. Kendler, K. S., Ohlsson, H., Sundquist, K. & Sundquist, J. Sources of Parent-Offspring Resemblance for Major Depression in a National Swedish Extended Adoption Study. JAMA Psychiatry 75, 194–200 (2018).

34. Lynskey, M. T. et al. Genetic and environmental contributions to cannabis dependence in a national young adult twin sample. Psychol. Med. 32, 195–207 (2002).

35. Baker, J. H., Maes, H. H. & Kendler, K. S. Shared environmental contributions to substance use. Behav. Genet. 42, 345–353 (2012).

36. Clarke, T.-K. et al. Genetic and shared couple environmental contributions to smoking and alcohol use in the UK population. Mol. Psychiatry 26, 4344–4354 (2019).

37. Silventoinen, K. et al. Genetic and environmental variation in educational attainment: an individual-based analysis of 28 twin cohorts. Sci. Rep. 10, 12681 (2020).

38. Mullins, N. et al. Genome-wide association study of more than 40,000 bipolar disorder cases provides new insights into the underlying biology. Nat. Genet. 53, 817–829 (2021).

39. Trubetskoy, V. et al. Mapping genomic loci implicates genes and synaptic biology in schizophrenia. Nature 604, 502–508 (2022).

40. Wang, X. et al. Polygenic risk prediction: why and when out-of-sample prediction R2 can exceed SNP-based heritability. Am. J. Hum. Genet. (2023) doi:10.1016/j.ajhg.2023.06.006.

41. Duncan, L. et al. Analysis of polygenic risk score usage and performance in diverse human populations. Nat. Commun. 10, 3328 (2019).

42. Schork, A. et al. Exploring contributors to variability in estimates of SNP-heritability and genetic correlations from the iPSYCH case-cohort and published meta-studies of major psychiatric disorders. bioRxiv 487116 (2019) doi:10.1101/487116.

43. Cai, N. et al. Minimal phenotyping yields genome-wide association signals of low specificity for major depression. Nat. Genet. 52, 437–447 (2020).

44. Yang, J., Wray, N. R. & Visscher, P. M. Comparing apples and oranges: equating the power of case-control and quantitative trait association studies. Genet. Epidemiol. 34, 254–257 (2010).

45. Yengo, L. et al. A saturated map of common genetic variants associated with human height. Nature 610, 704–712 (2022).

46. Pingault, J.-B. et al. Genetic sensitivity analysis: Adjusting for genetic confounding in epidemiological associations. PLoS Genet. 17, e1009590 (2021).

47. Pedersen, C. B. et al. The iPSYCH2012 case-cohort sample: new directions for unravelling genetic and environmental architectures of severe mental disorders. Mol. Psychiatry 23, 6–14 (2018).

48. Bybjerg-Grauholm, J. et al. The iPSYCH2015 Case-Cohort sample: updated directions for unravelling genetic and environmental architectures of severe mental disorders. bioRxiv (2020) doi:10.1101/2020.11.30.20237768.

49. Nørgaard-Pedersen, B. & Hougaard, D. M. Storage policies and use of the Danish Newborn Screening Biobank. J. Inherit. Metab. Dis. 30, 530–536 (2007).

50. Mors, O., Perto, G. P. & Mortensen, P. B. The Danish Psychiatric Central Research Register. Scand. J. Public Health 39, 54–57 (2011).

51. Lynge, E., Sandegaard, J. L. & Rebolj, M. The Danish National Patient Register. Scand. J. Public Health 39, 30–33 (2011).

52. Pedersen, C. B. The Danish Civil Registration System. Scand. J. Public Health 39, 22–25 (2011).

53. Appadurai, V. et al. Accuracy of haplotype estimation and whole genome imputation affects complex trait analyses in complex biobanks. bioRxiv 2022.06.27.497703 (2022) doi:10.1101/2022.06.27.497703.

54. Browning, B. L., Zhou, Y. & Browning, S. R. A One-Penny Imputed Genome from Next-Generation Reference Panels. Am. J. Hum. Genet. 103, 338–348 (2018).

55. Browning, S. R. & Browning, B. L. Rapid and accurate haplotype phasing and missing-data inference for whole-genome association studies by use of localized haplotype clustering. Am. J. Hum. Genet. 81, 1084–1097 (2007).

56. McCarthy, S. et al. A reference panel of 64,976 haplotypes for genotype imputation. Nat. Genet. 48, 1279–1283 (2016).

57. Manichaikul, A. et al. Robust relationship inference in genome-wide association studies. Bioinformatics 26, 2867–2873 (2010).

58. Price, A. L. et al. Principal components analysis corrects for stratification in genome-wide association studies. Nat. Genet. 38, 904–909 (2006).

59. Athanasiadis, G. et al. A comprehensive map of genetic relationships among diagnostic categories based on 48.6 million relative pairs from the Danish genealogy. Proc. Natl. Acad. Sci. U. S. A. 119, (2022).

60. Rainer, J. et al. FamAgg: an R package to evaluate familial aggregation of traits in large pedigrees. Bioinformatics 32, 1583–1585 (2016).

61. Sinnwell, J. P., Therneau, T. M. & Schaid, D. J. The kinship2 R package for pedigree data. Hum. Hered. 78, 91–93 (2014).

62. Wright, S. Coefficients of Inbreeding and Relationship. Am. Nat. 56, 330–338 (1922).

63. Lloyd-Jones, L. R. et al. Improved polygenic prediction by Bayesian multiple regression on summary statistics. Nat. Commun. 10, 5086 (2019).

64. Howard, D. M. et al. Genome-wide meta-analysis of depression identifies 102 independent variants and highlights the importance of the prefrontal brain regions. Nat. Neurosci. 22, 343–352 (2019).

65. Zhou, W. et al. Efficiently controlling for case-control imbalance and sample relatedness in large-scale genetic association studies. Nat. Genet. 50, 1335–1341 (2018).

66. Golan, D., Lander, E. S. & Rosset, S. Measuring missing heritability: inferring the contribution of common variants. Proc. Natl. Acad. Sci. U. S. A. 111, E5272–81 (2014).

67. Dudbridge, F. Power and predictive accuracy of polygenic risk scores. PLoS Genet. 9, e1003348 (2013).

68. Vilhjálmsson, B. J. et al. Modeling Linkage Disequilibrium Increases Accuracy of Polygenic Risk Scores. Am. J. Hum. Genet. 97, 576–592 (2015).

69. Wright, S. The Method of Path Coefficients. aoms 5, 161–215 (1934).

