## Supplementary Figures for "The relationship between genotype- and phenotype-based estimates of genetic liability to psychiatric disorders, in practice and in theory"

**Supplementary Figure S1.** Common liability scale estimators give qualitatively different estimates of liability scale variance explained for phenotype-based genetic instruments.
**Supplementary Figure S2.** Common estimates of variance explained on the liability scale can show large bias in simulations**.**
**Supplementary Figure S3.** Nagelkerke's pseudo r2 from logistic model fitting.
**Supplementary Figure S4.** Impact of mutual adjustment on odds ratios for PGS, FGRS and FH in iPSYCH-2012
**Supplementary Figure S5.** Impact of mutual adjustment on odds ratios for PGS, FGRS and FH in iPSYCH-2015i
**Supplementary Figure S6.** Full Correlation among all predictors - iPSYCH 2012 random cohort.
**Supplementary Figure S7.** Full Correlation among all predictors - iPSYCH 2015i random cohort.
**Supplementary Figure S8.** Expected accuracy of family genetic risk scores from theory and in simulated data based on records of full-siblings.
**Supplementary Figure S9.** Expected accuracy of family genetic risk scores from theory and in simulated data based on records of offspring with different mates.
**Supplementary Figure S10.** Expected accuracy of family genetic risk scores from theory and in simulated data based on records of siblings of one parent.
**Supplementary Figure S11.** Expected accuracy of family genetic risk scores from theory and in simulated data based on records of half-sibs that are also half-sibs of each other.
**Supplementary Figure S12.** Expected accuracy of family genetic risk scores from theory and in simulated data based on records of first cousins that are also first cousins of each other.
**Supplementary Figure S13.** Estimators of the Pearson correlation of disease status given liability correlation and prevalence.
**Supplementary Figure S14.** Empirical and expected reliability of a linear predictor
**Supplementary Figure S15.** Empirical and expected reliability of the PA-FGRS
**Supplementary Figure S16.** Exact and approximate asymptotes of accuracy of a linear predictor based on sibling information.
**Supplementary figure S17.** Sibling equivalents for disease traits with different prevalence and heritability.
**Supplementary Figure S18.** Impact of shared environment on risk of phenotypic recurrence.
**Supplementary Figure S19.** Impact of shared environment on squared accuracy and performance of PA-FGRS relative to expectations.
**Supplementary Figure S20.** Impact of shared environment on performance of PGS relative to expectations.
**Supplementary Figure S21.** Impact of shared environment on correlation between PGS and PA-FGRS relative to expectations.
**Supplementary Figure S22.** Choice of “M” and FGRS-PGS relationship.
**Supplementary Figure S23.** Choice of h2_snp / h2 and FGRS-PGS relationship
**Supplementary Figure S24.** Choice of p and FGRS-PGS relationship.
**Supplementary Figure S25.** A diagram depicting our generative liability model and the implied relationships among PGS, PA-FGRS, and liability.
**Supplementary Figure S26.** Choice of M in PGS-FGRS correlation
**Supplementary Figure S27.** Choice of $h^2_{SNP}$ in PGS-FGRS correlation
**Supplementary Figure S28.** Choice of p in PGS, PA-FGRS correlation
**Supplementary Figure S29.** Choice of M in PGS-FGRS joint prediction
**Supplementary Figure S30.** Choice of $h^2_{SNP}$ in PGS-FGRS joint prediction
**Supplementary Figure S31.** Choice of p in PGS-FGRS joint prediction

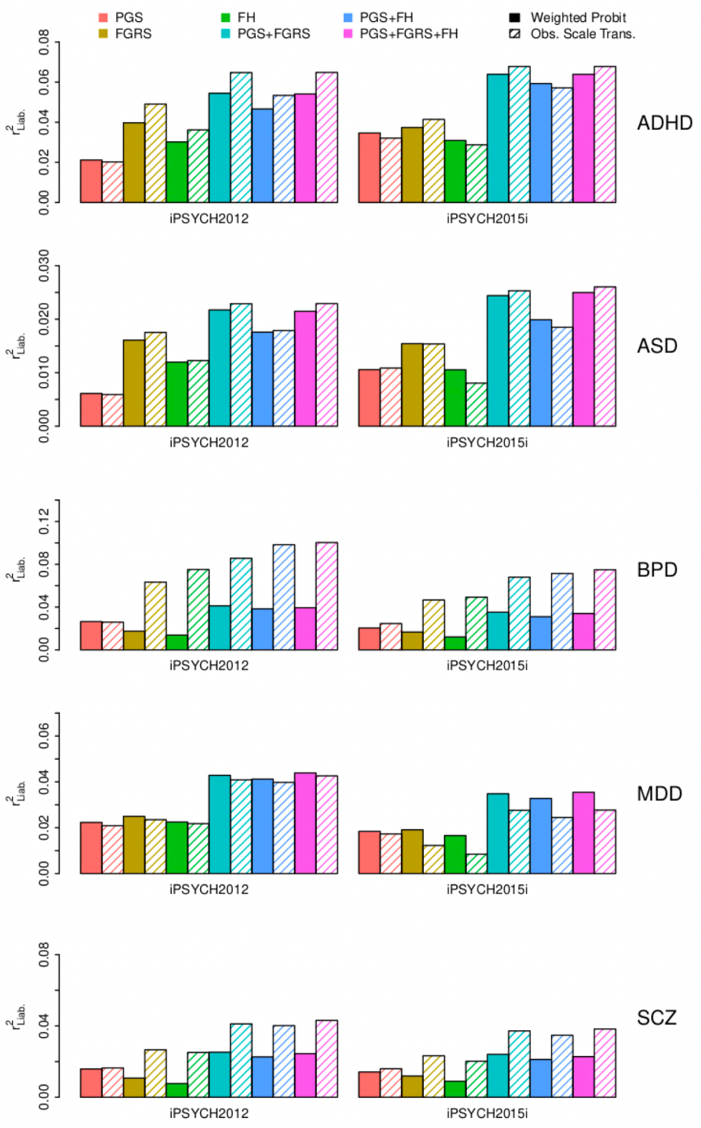


**Supplementary Figure S1. Common liability scale estimators give qualitatively different estimates of liability scale variance explained for phenotype-based genetic instruments.**

The estimated liability explained by polygenic score (PGS), family genetic risk score (FGRS), the indicator variable for having an affected parent or sibling (FH), or their combinations (PGS+FGRS, PGS+FH, PGS+FGRS+FH) can vary depending on estimation approach. The solid bars are estimates of liability explained by a two-stage weighted probit model whereas the striped bars are estimated using transformations from linear regression estimates on the observed scale. With the exception of the PGS alone model, the estimated liability scale variance tends to be larger when applying the observed scale transformations, which we believe to be incorrect.

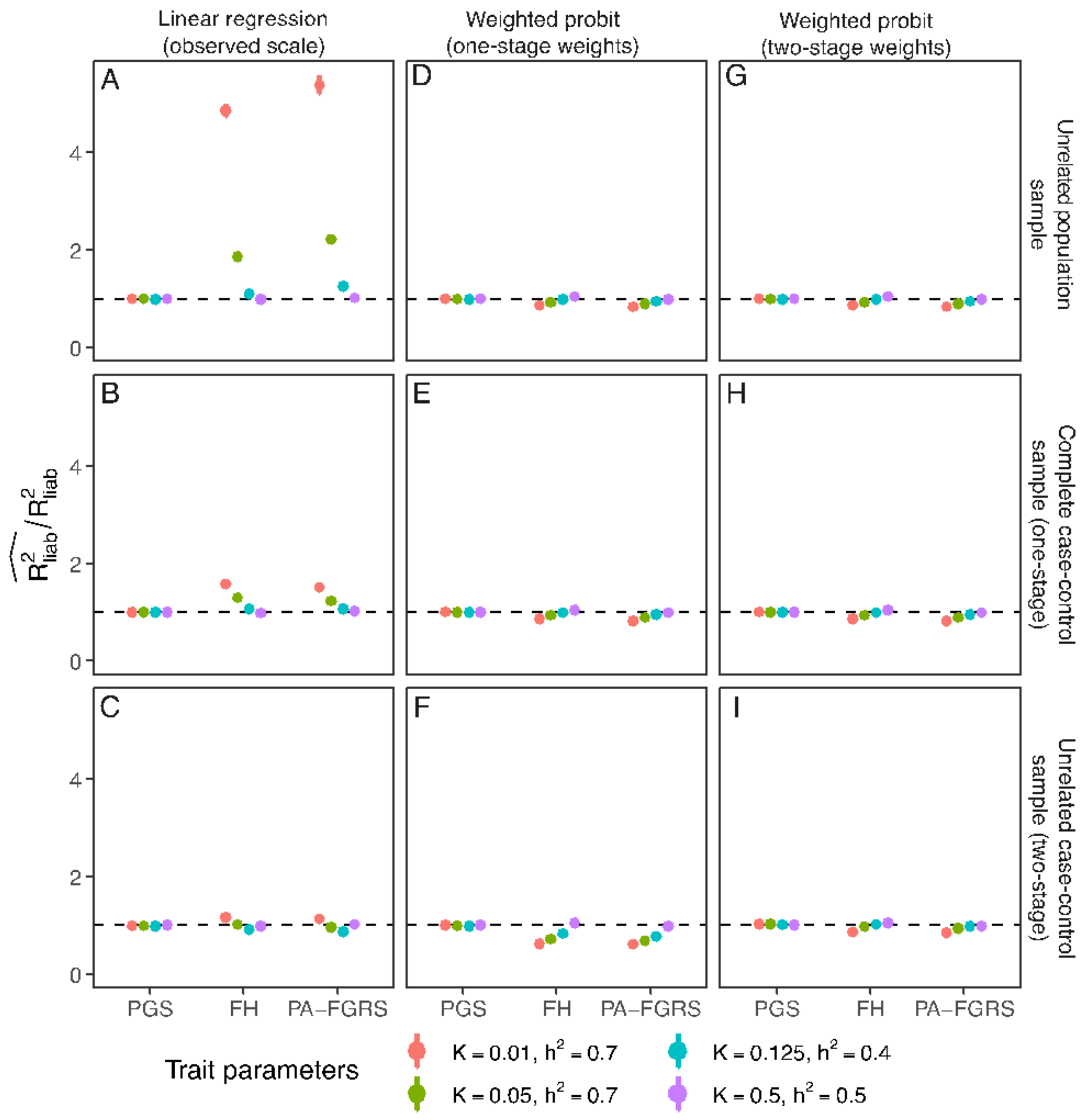


**Supplementary Figure S2. Common estimates of variance explained on the liability scale are biased for phenotype-based genetic instruments.** In a simulated population of families, we compare different strategies for estimating liability scale variance explained by PGS, FH, and PA-FGRS, under different sampling schemes. The dashed line in each plot represents the ratio of estimated to true liability scale variance explained where values deviating from 1 indicate biased estimates. Transforming variance explained from the observed scale (A,B,C) is only appropriate for PGS, but produces biased estimates for FH and PA-FGRS across nearly all sampling and trait configurations tested. Directly estimating liability scale variance using a weighted probit regression to account for one-stage case-control sampling (D,E,F) corrects for this bias, but overcorrects when two-stage sampling (e.g., pruning of relatives) is applied (F). The most robust estimates were found by directly estimating liability scale variance with a weighted probit regression that accounts for the two-stage sampling associated with case oversampling followed by relatedness pruning (G,H,I). When estimating variance explained on the liability scale for phenotype-based instruments, accounting for sampling beyond case-control proportion may be critical. Confidence intervals are generally narrower than and thus contained within each circle. Plotted simulation estimates are presented in Supplementary Table S3. PGS, polygenic score; FH, indicator of first-degree family history; PA-FGRS, Pearson-Aitken Family Genetic Risk Score; $R^2_{liab}$, Liability scale variance explained; K, lifetime prevalence; $h^2$, narrow-sense heritability.

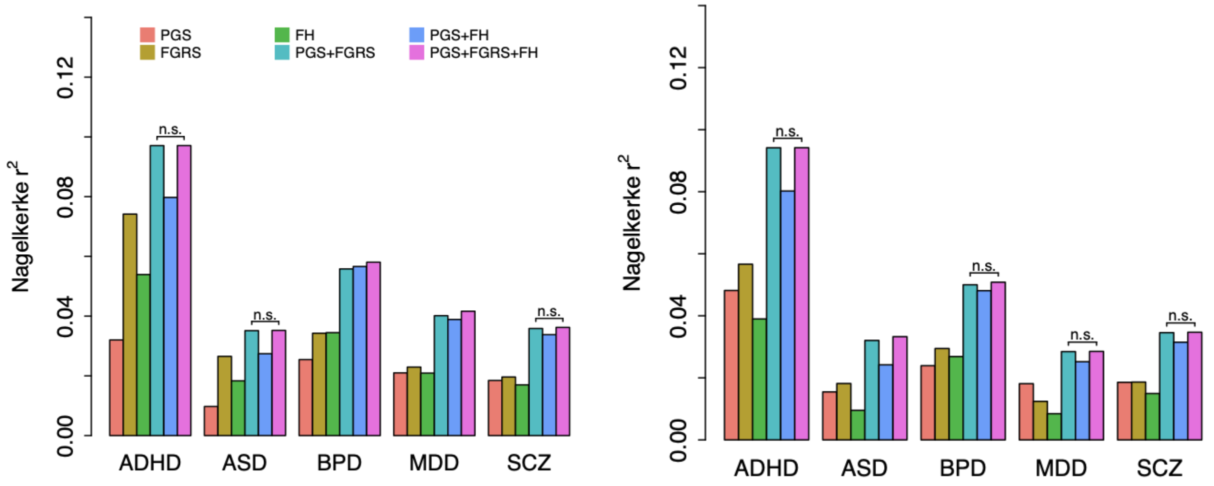


**Supplementary Figure S3. Nagelkerke's pseudo r2 from logistic model fitting.**

The figure displays Nagelkerke's pseudo r2 from logistic regression for each of the five mental disorders (attention deficit hyperactivity disorder (ADHD), autism spectrum disorder (ASD), bipolar disorder (BPD), major depressive disorder (MDD) and schizophrenia (SCZ) in the iPSYCH-2012 case-cohort and the non-overlapping iPSYCH-2015i case-cohort, by the disorder specific polygenic score (PGS), family genetic risk score (FGRS), the indicator variable for having an affected parent or sibling (FH), or their combinations (PGS+FGRS, PGS+FH, PGS+FGRS+FH) estimated as multiple regression.

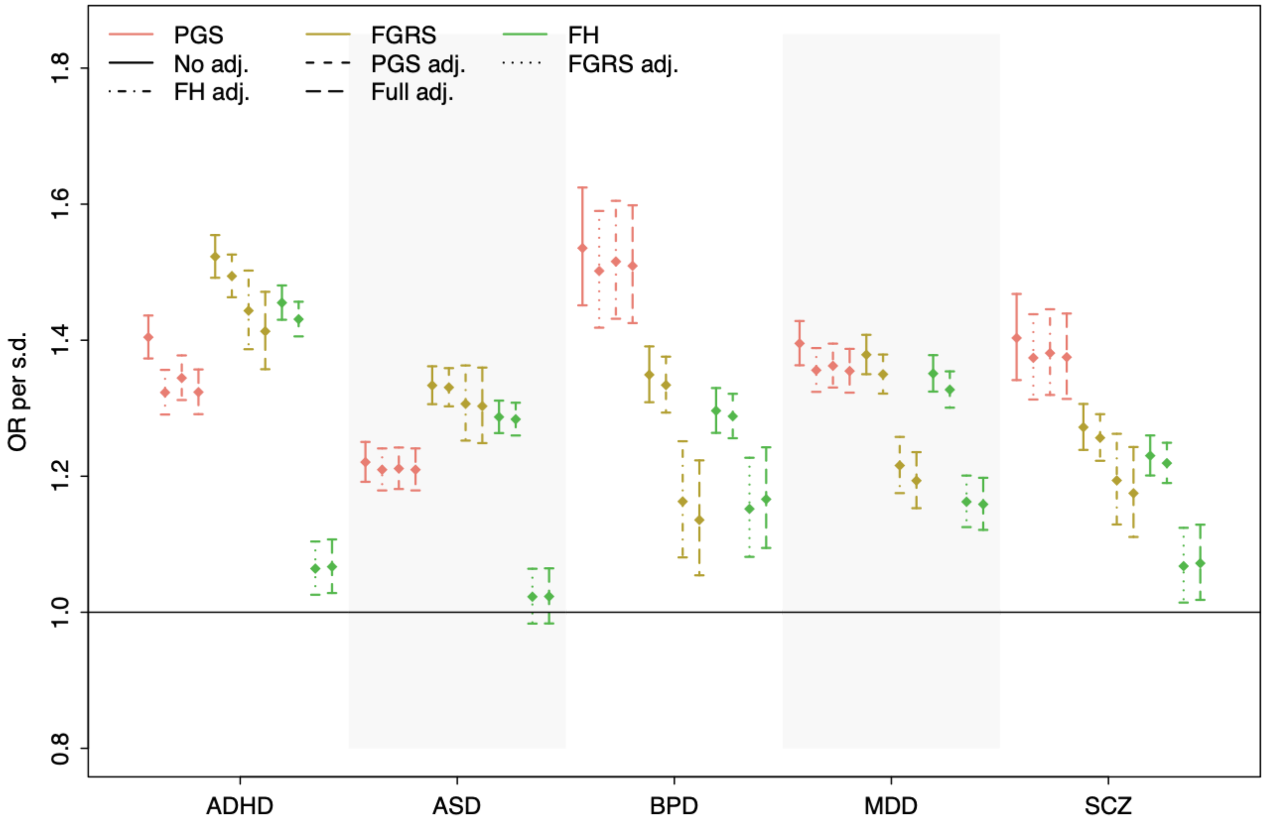


**Supplementary Figure S4. Impact of mutual adjustment on odds ratios for PGS, FGRS and FH in iPSYCH-2012**

The figure shows the odds ratio for the five different mental disorders associated with one standard deviation change in the predictor, when the predictor is the disorder specific PGS (red), PA-FGRS (yellow) or family history indicator (FH; green). The odds ratios are presented in an unadjusted model (solid lines), and with adjustment for each or both of the two other predictors. Each estimate is based on the disorder specific case cohort from the iPSYCH 2012 data.

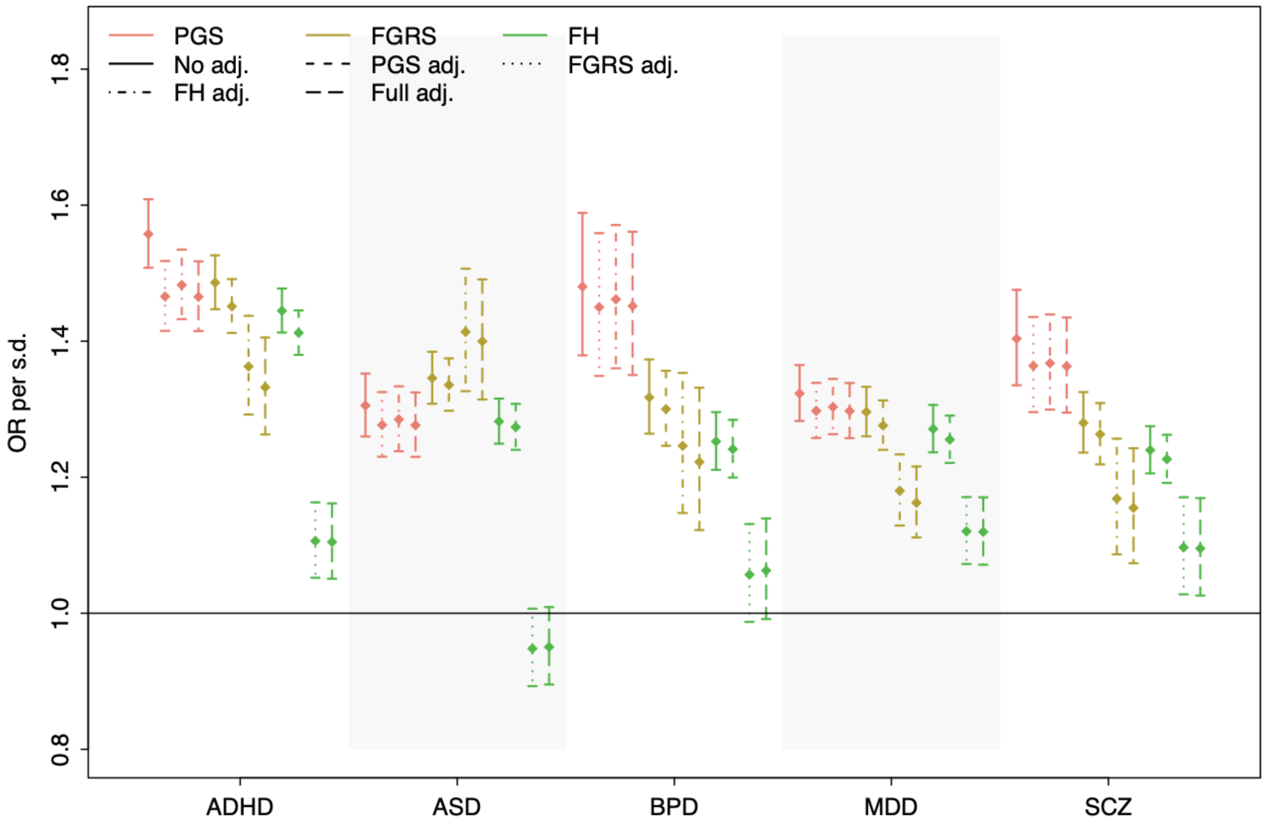


**Supplementary Figure S5. Impact of mutual adjustment on odds ratios for PGS, FGRS and FH in iPSYCH-2015i**

The figure shows the odds ratio for the five different mental disorders associated with one standard deviation change in the predictor, when the predictor is the disorder specific PGS (red), PA-FGRS (yellow) or family history indicator (FH; green). The odds ratios are presented in an unadjusted model (solid lines), and with adjustment for each or both of the two other predictors. Each estimate is based on the disorder specific case control sample from the iPSYCH 2015i data.

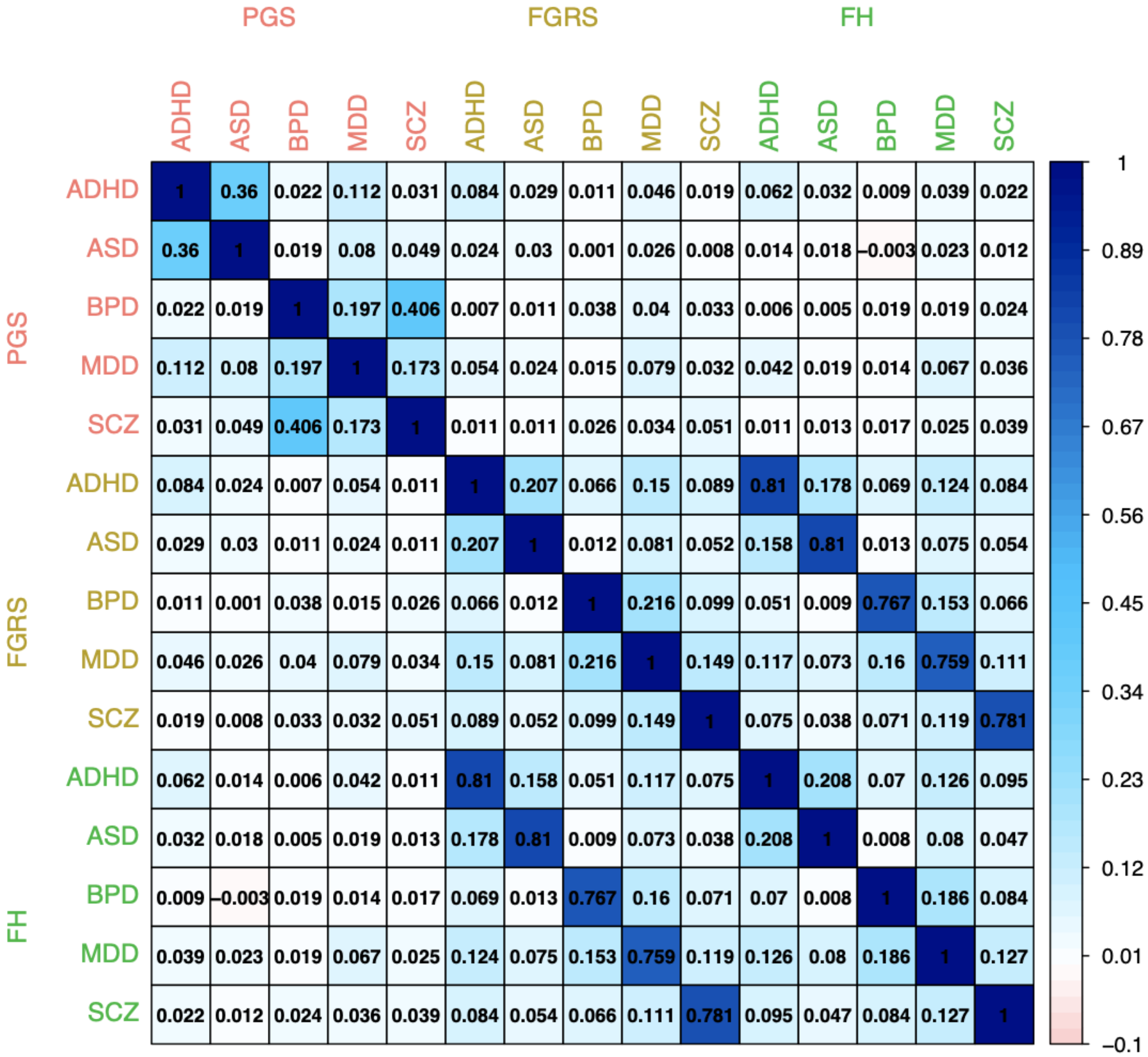


**Supplementary Figure S6. Full Correlation among all predictors - iPSYCH 2012 random cohort.**
Estimated Pearson correlation coefficient between the polygenic score (PGS), the Family Genetic risk score (FGRS) and the family history indicator (FH) in the random population cohort of iPSYCH-2012 (N=24,266).
Notes: liab221015, FH = fgrs_ps > 0

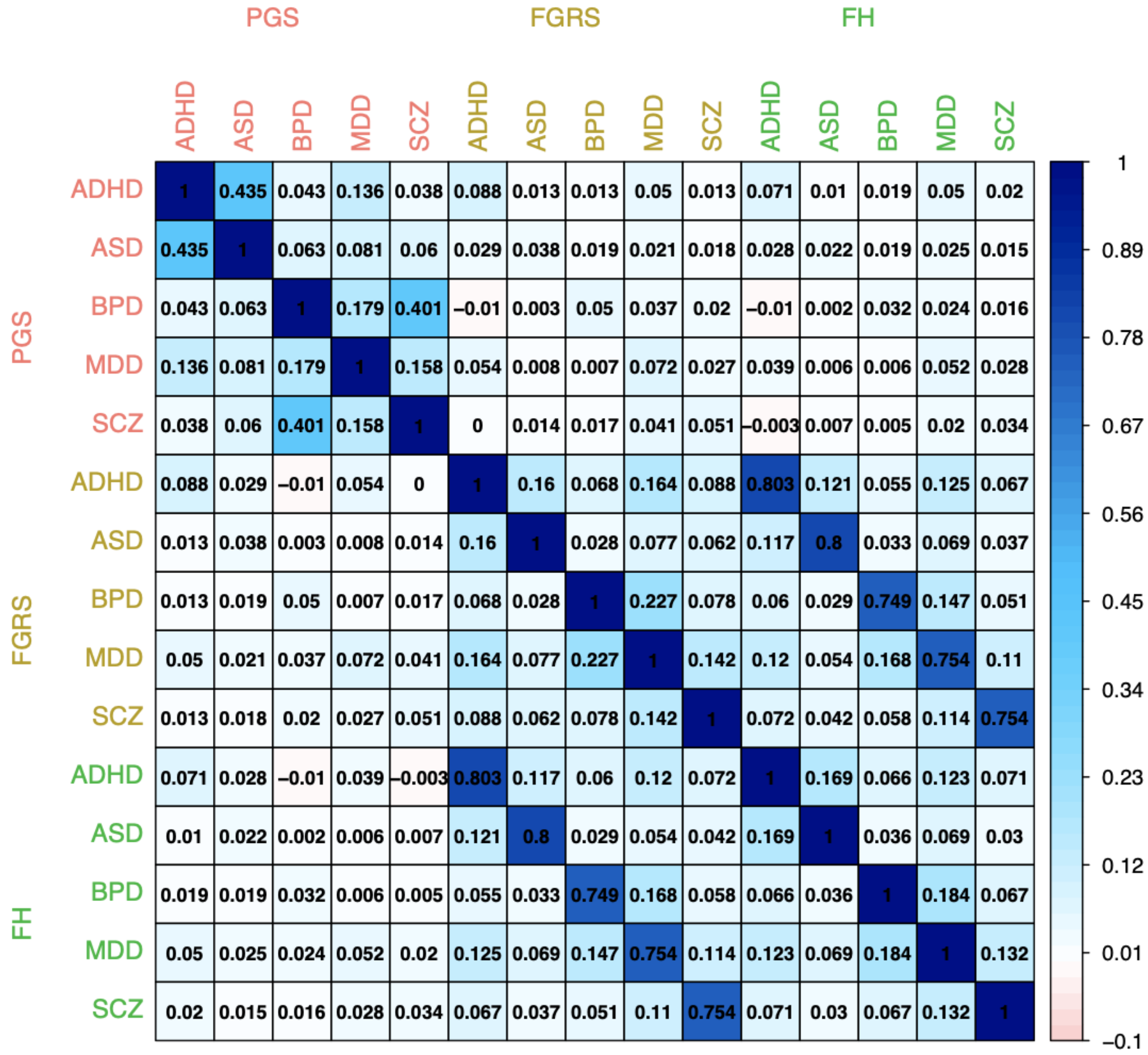


**Supplementary Figure S7. Full Correlation among all predictors - iPSYCH 2015i random cohort.**
Estimated Pearson correlation coefficient between the polygenic score (PGS), the Family Genetic risk score (FGRS) and the family history indicator (FH) in the random population cohort of iPSYCH-2015i (N=15,381).
Notes: liab221015, FH = fgrs_ps > 0

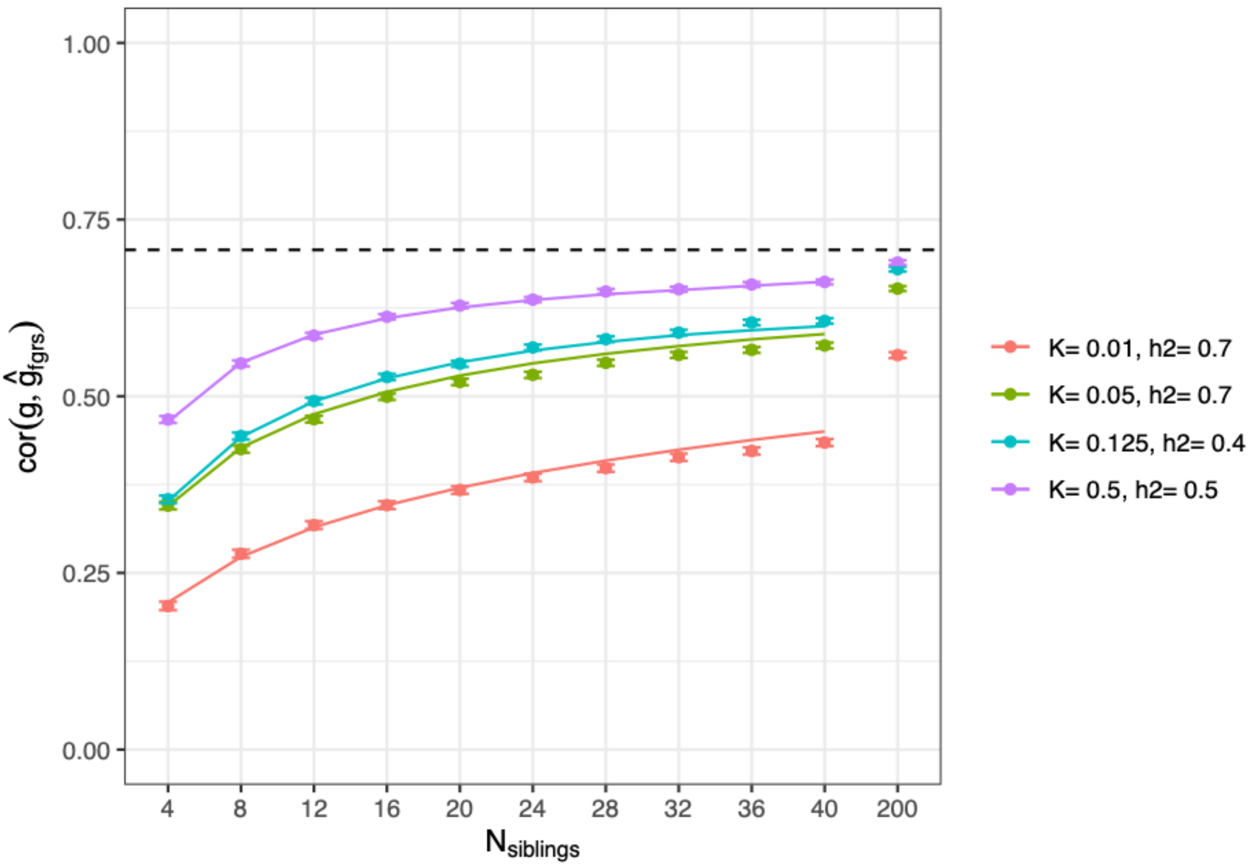


**Supplementary Figure S8. Expected accuracy of family genetic risk scores from theory and in simulated data based on records of full-siblings.**

We simulate 100 000 index individuals and a varying number of relatives and assess the correlation between the predicted and the true liability (dots with error bars for 95%-confidence intervals). We compare this to the expected accuracy given *Eq.2* (solid line). Colors indicate the prevalence and heritability of the phenotype.

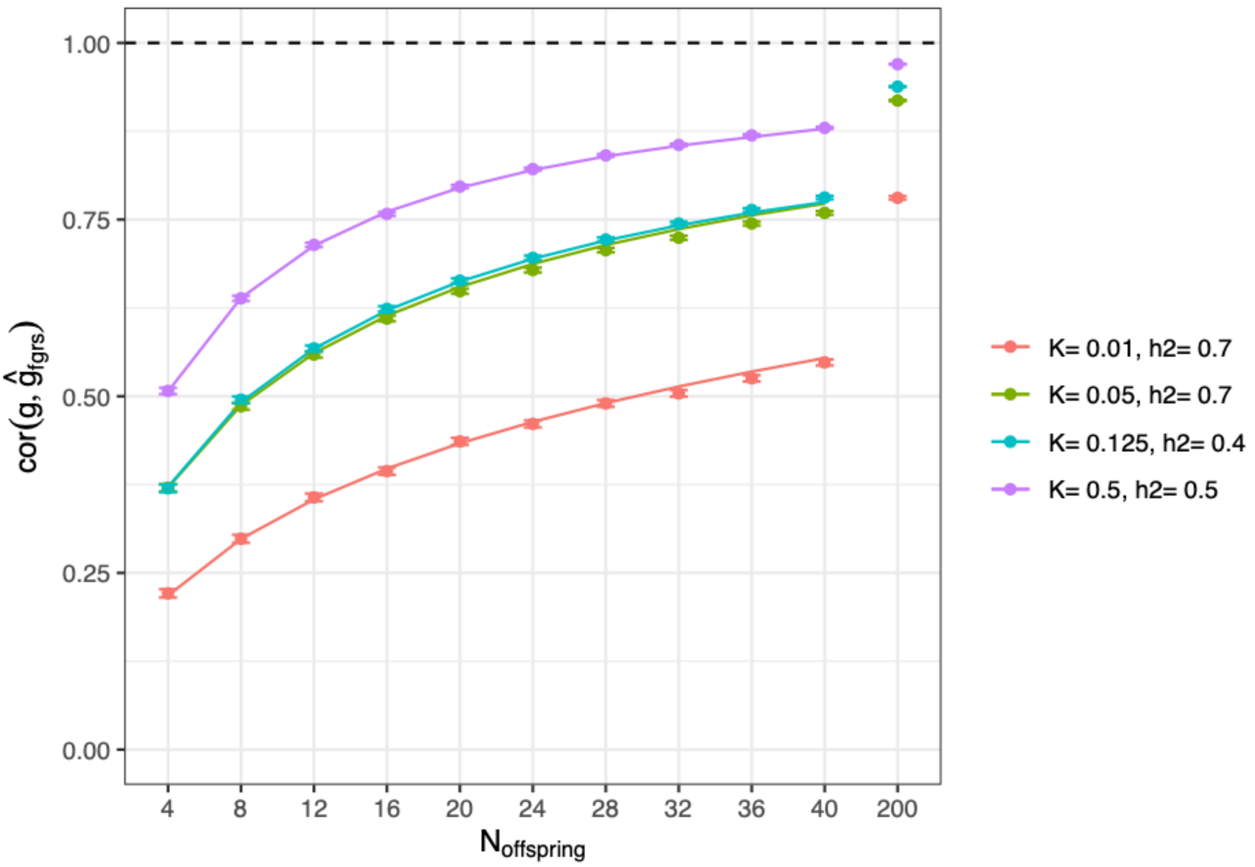


**Supplementary Figure S9. Expected accuracy of family genetic risk scores from theory and in simulated data based on records of offspring with different mates.**
We simulate 100 000 index individuals and a varying number of relatives and assess the correlation between the predicted and the true liability (dots with error bars for 95%-confidence intervals). We compare this to the expected accuracy given *Eq.2* (solid line). Colors indicate the prevalence and heritability of the phenotype.

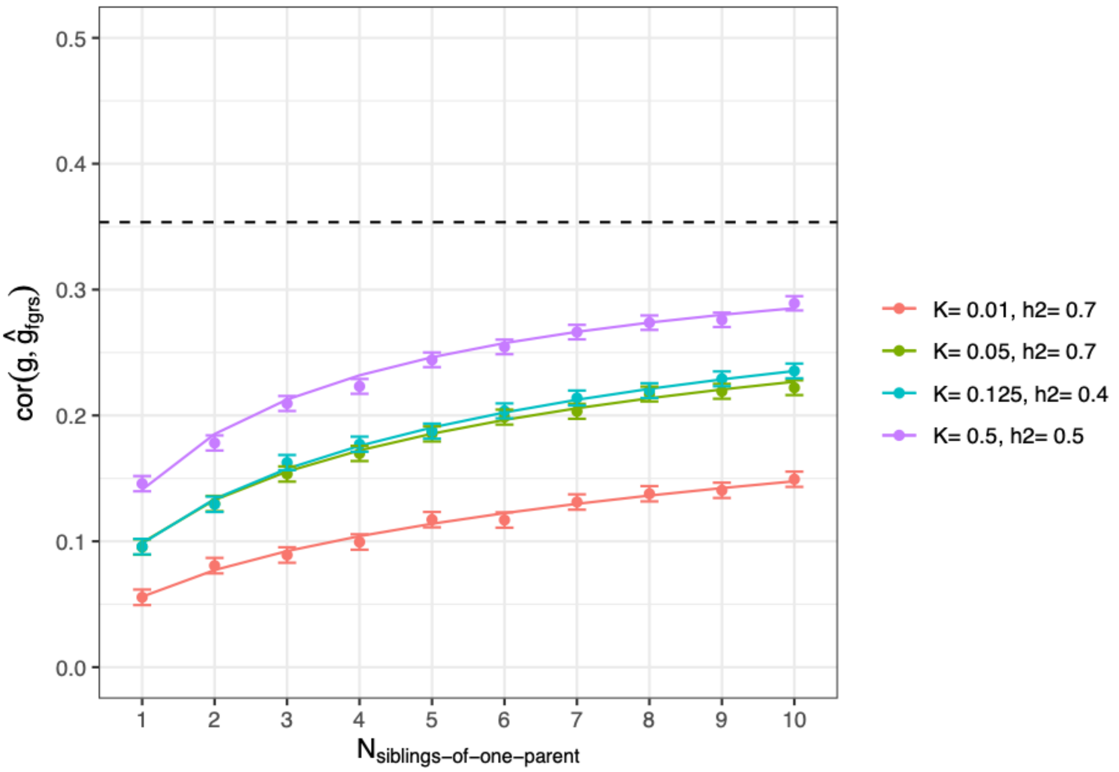


**Supplementary Figure S10. Expected accuracy of family genetic risk scores from theory and in simulated data based on records of siblings of one parent.**

We simulate 100 000 index individuals and a varying number of relatives and assess the correlation between the predicted and the true liability (dots with error bars 95%-confidence intervals). We compare this to the expected accuracy given *Eq.2* (solid line). Colors indicate the prevalence and heritability of the phenotype.

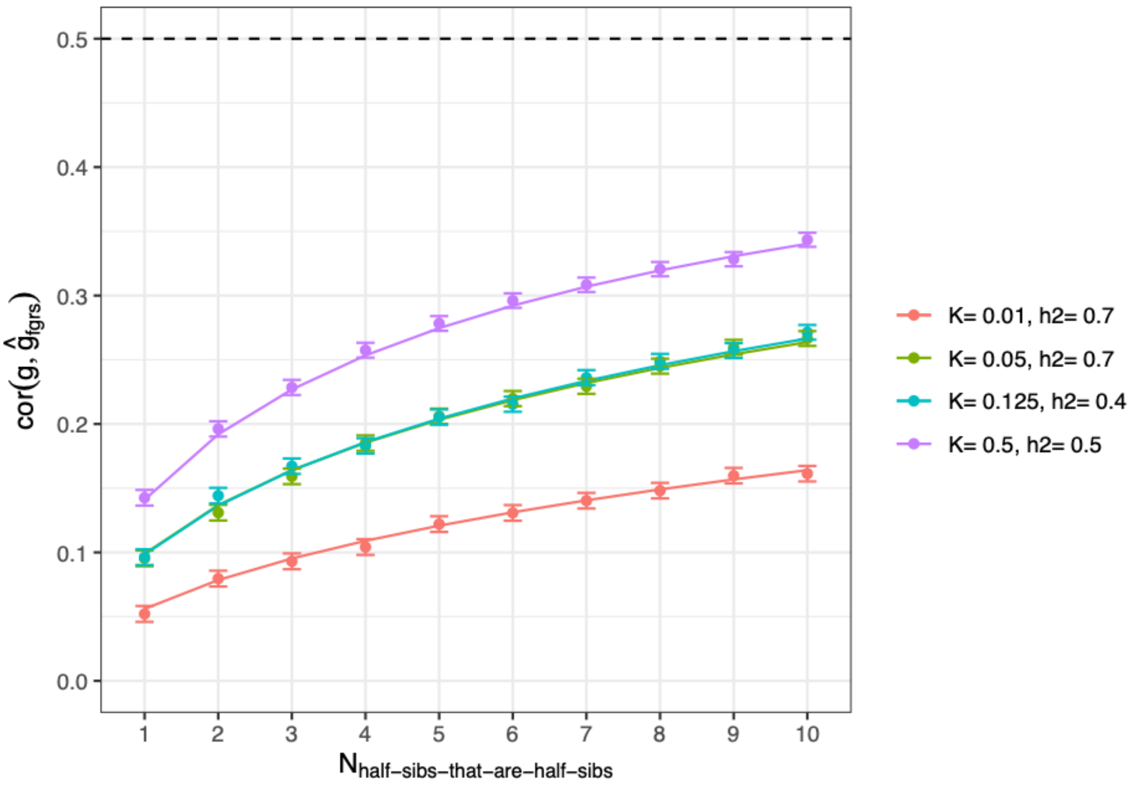


**Supplementary Figure S11. Expected accuracy of family genetic risk scores from theory and in simulated data based on records of half-sibs that are also half-sibs of each other.**
We simulate 100 000 index individuals and a varying number of relatives and assess the correlation between the predicted and the true liability (dots with error bars 95%-confidence intervals). We compare this to the expected accuracy given *Eq.2* (solid line). Colors indicate the prevalence and heritability of the phenotype.

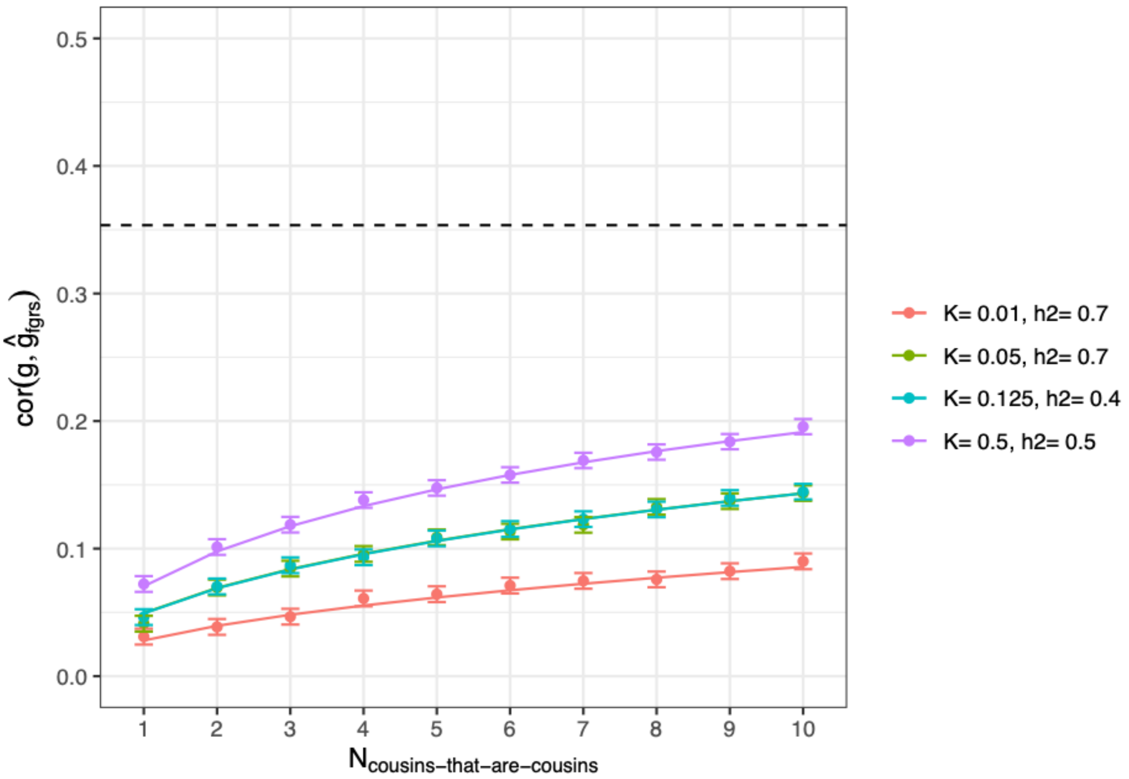


**Supplementary Figure S12. Expected accuracy of family genetic risk scores from theory and in simulated data based on records of first cousins that are also first cousins of each other.**

We simulate 100 000 index individuals and a varying number of relatives and assess the correlation between the predicted and the true liability (dots with error bars 95%-confidence intervals). We compare this to the expected accuracy given *Eq.2* (solid line). Colors indicate the prevalence and heritability of the phenotype.

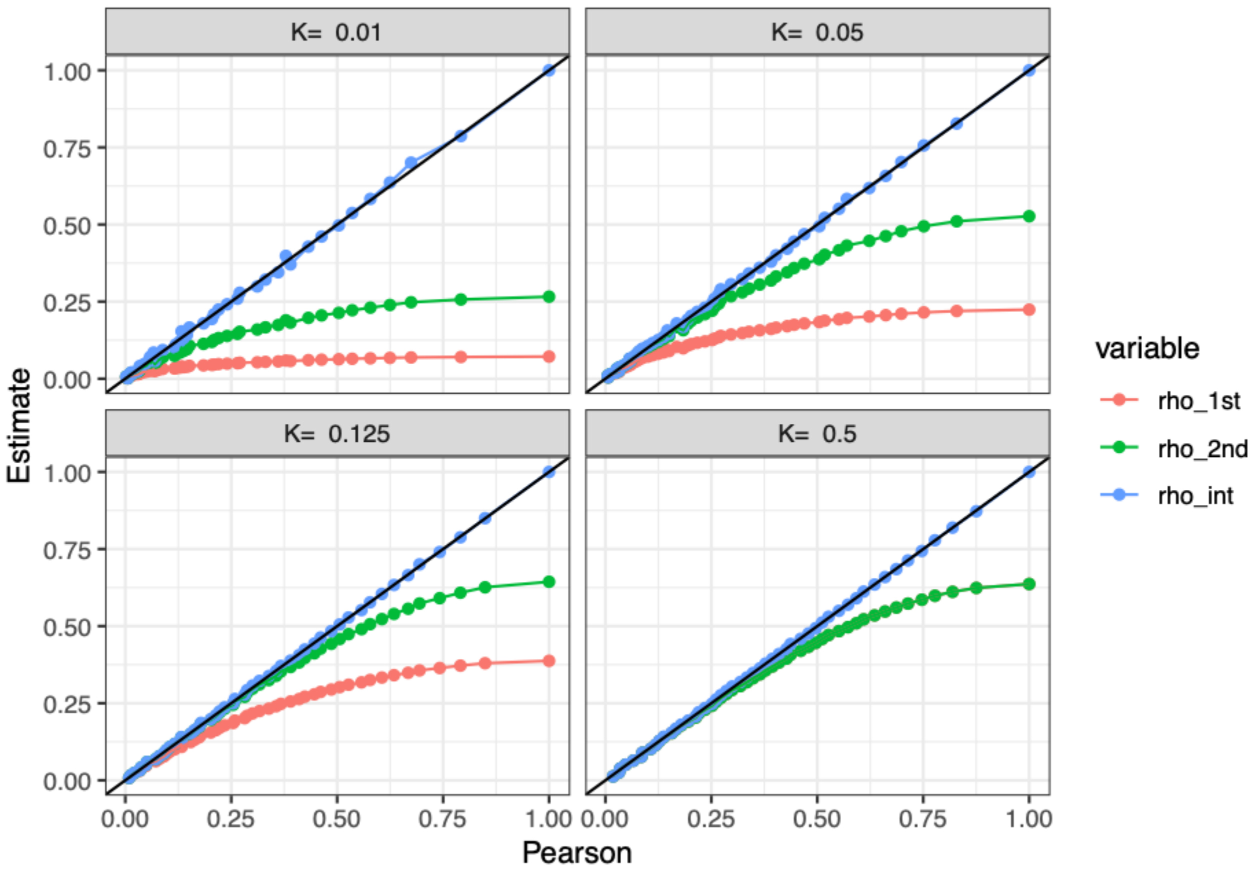


**Supplementary Figure S13. Estimators of the Pearson correlation of disease status given liability correlation and prevalence.**

The figure displays the relationship between the empirical pearson correlation (x-axis) and the expected pearson correlation (y-axis). The empirical pearson correlations are obtained by generating 100K samples from a standard bivariate normal distribution with correlation varying from 0 to 1 (at 0.02 increments) , and dichotomizing into disease status at threshold $t = \Phi^{-1}(1 - K)$ setting K to 0.01, 0.05, 0.125 or 0.5, and the expected pearson correlation (y-axis) are obtained by the three different approximations: the first order taylor approximation (rho_1st, $\rho_{1st} = \frac{\phi(t)^2 r_{rr} h^2}{K(1-K)}$) , the second order taylor approximation (rho_2nd, $\rho_{2nd} = \frac{\phi(t)^2 r_{rr} h^2}{K(1-K)} + \frac{t^2 \phi(t)^2 h^4 r_{rr}^2}{2K(1-K)}$) and the approximation obtained by the multivariate integration (rho_int, $\rho_{exact} = \frac{\int_t^{\infty}\int_t^{\infty} \phi_2(x, y, r_{rr} h^2)\, dx\, dy - K^2}{K(1-K)}$). Note that the first and second order approximations become identical at K=0.5 since t=0.

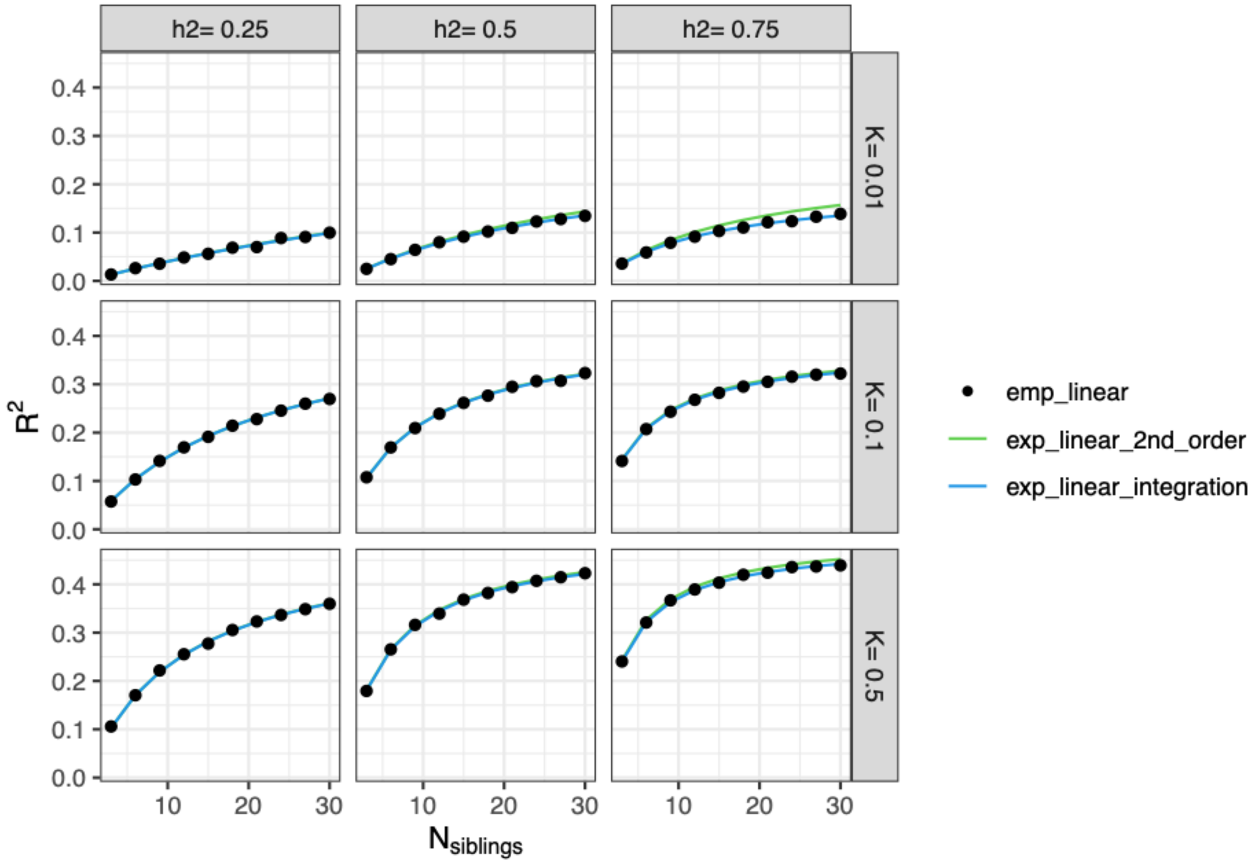


**Supplementary Figure S14. Empirical and expected reliability of a linear predictor**

The figure displays the empirical reliability of a linear predictor of genetic liability (dots) obtained by simulating 100K individuals with 1-30 siblings and the expected reliability given by the second order approximation $R^2 = \frac{n_{rel} r_{ir}^2}{\frac{K(1-K)}{\phi(t)^2 h^2} + (n_{rel}-1) r_{rr}\left(1+\frac{r_{rr} h^2 t^2}{2}\right)}$ (from Equation S5; green line) and the estimator based on multivariate integration $R^2 = \frac{n_{rel} h^2 r_{ir}^2 \phi(t)^2}{K(1-K) + (n_{rel}-1)\left(\int_t^{\infty}\int_t^{\infty} \phi_2(x,y,r_{rr}h^2)\, dx\, dy - K^2\right)}$ (from Equation S6; green line). The panels denote different combinations of heritability (0.25,0.5 and 0.75) and prevalence (0.01,0.1 and 0.5).

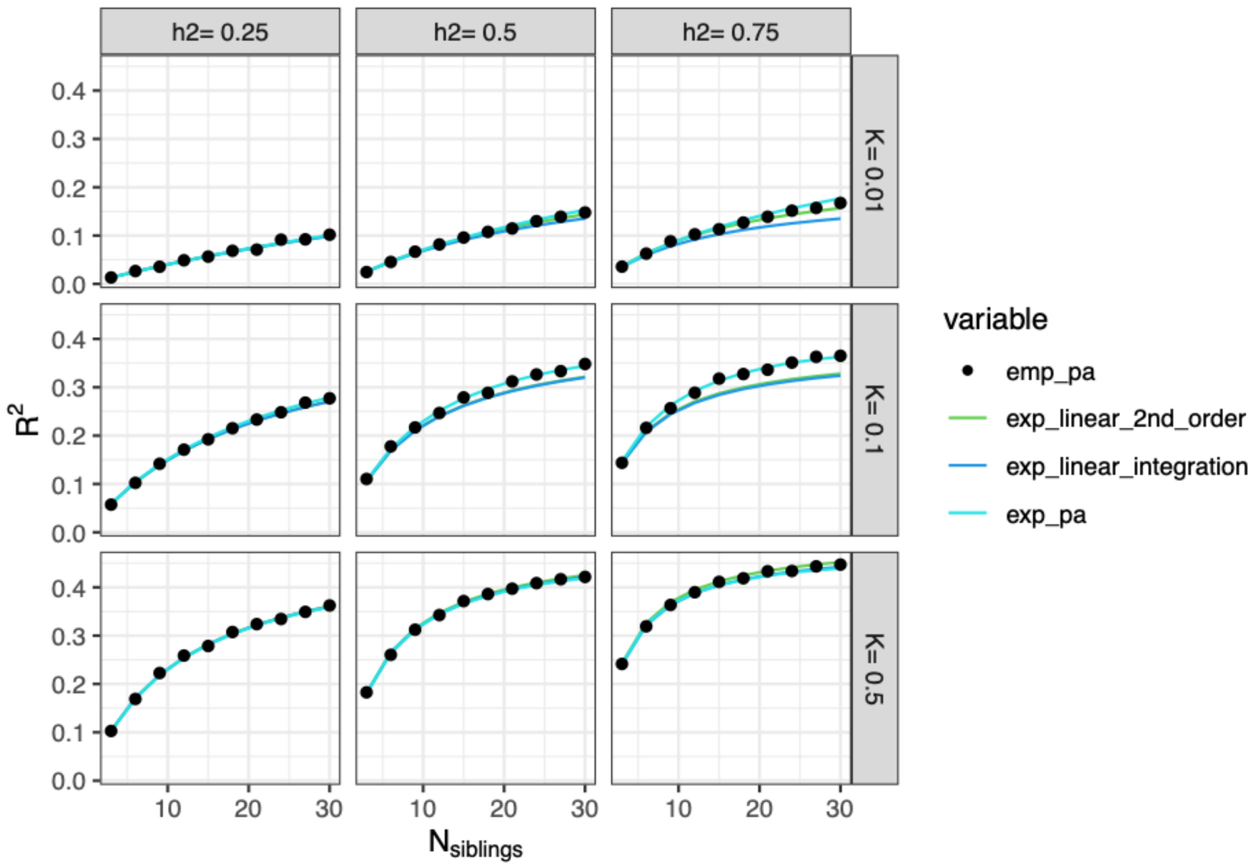


**Supplementary Figure S15. Empirical and expected reliability of the PA-FGRS**

The figure displays the empirical reliability of a pa-fgrs of genetic liability (dots) obtained by simulating 100K individuals with 1-30 siblings and the expected reliability given by the second order approximation $R^2 = \frac{n_{rel} r_{ir}^2}{\frac{K(1-K)}{\phi(t)^2 h^2} + \left(n_{rel}-1\right) r_{rr}\left(1+\frac{r_{rr} h^2 t^2}{2}\right)}$ (from Equation S5; green line), the estimator based on multivariate integration $R^2 = \frac{n_{rel} h^2 r_{ir}^2 \phi(t)^2}{K(1-K) + \left(n_{rel}-1\right)\left(\int_t^\infty \int_t^\infty \Phi_2(x, y, r_{rr} h^2)\, dx\, dy - K^2\right)}$ (from Equation S6; green line), and the exact estimator of the expected accuracy of the non-linear predictor $\frac{\sum_{n_{aff}=0}^{n_{rel}} \left(\frac{n_{rel}}{n_{aff}} P\left(\sum_{j=1}^{n_{rel}} D_j = n_{aff}, n_{rel}, \Sigma, K\right) \mathrm{E}\left(G \mid \sum_{j=1}^{n_{rel}} D_j = n_{aff}, n_{rel}, \Sigma, K\right)^2\right)}{h^2}$ (from Equation S13; turquoise line). The panels denote different combinations of heritability (0.25,0.5 and 0.75) and prevalence (0.01,0.1 and 0.5).

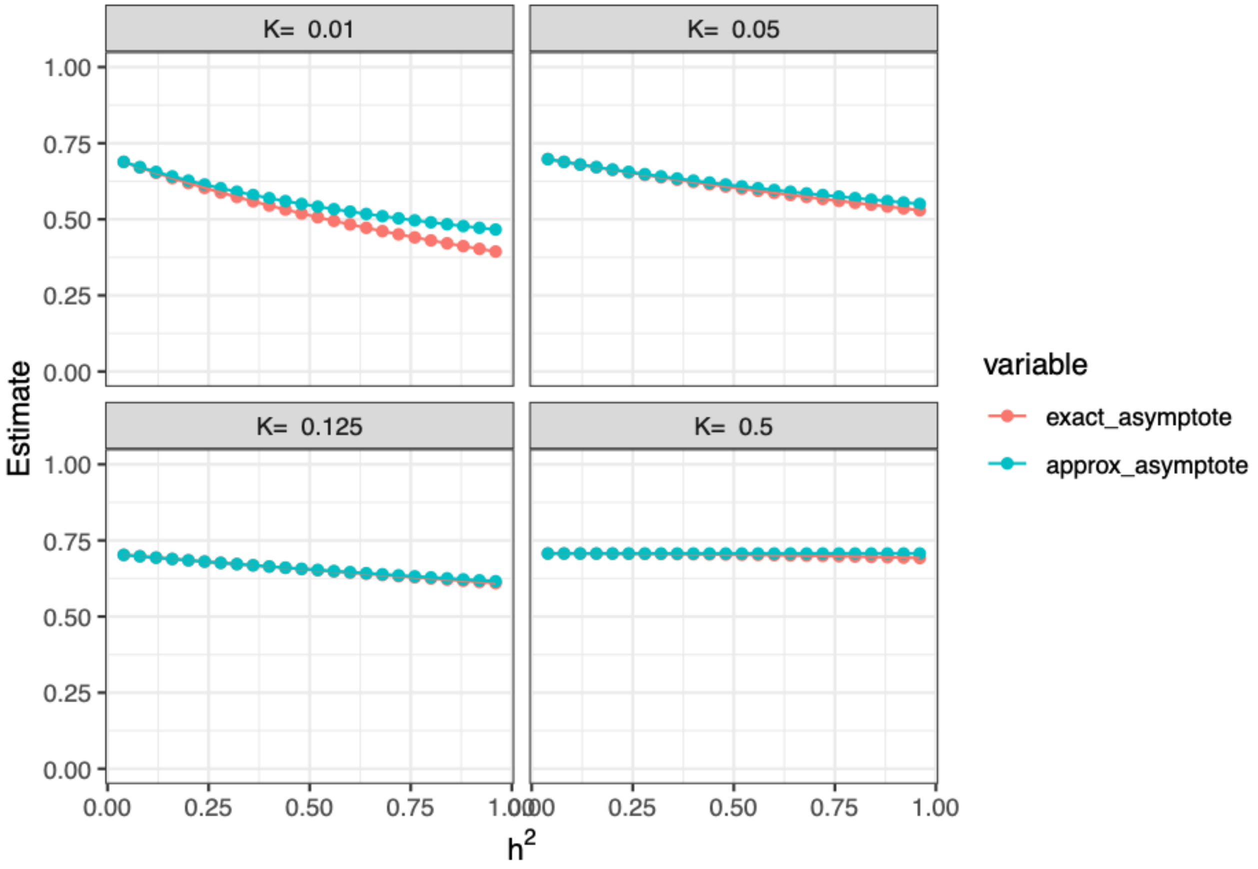


**Supplementary Figure S16. Exact and approximate asymptotes of accuracy of a linear predictor based on sibling information.**

The figure displays how the asymptote of the accuracy of a linear predictor depends on both the heritability ($h^2$; x-axis) and the prevalence (K; panels). The red line shows the exact asymptote of a linear predictor given by $\sqrt{\frac{h^2 r_{ir}^2 \phi(t)^2}{\int_t^{\infty}\int_t^{\infty} \Phi_2(x,y,r_{rr}h^2)\,dx\,dy - K^2}}$ (Equation S7), while the turquoise line shows the approximate asymptote given by the simpler expression $\frac{r_{ir}}{\sqrt{r_{rr}}}\frac{1}{\sqrt{1+\frac{r_{rr}h^2t^2}{2}}}$ (Equation S8).

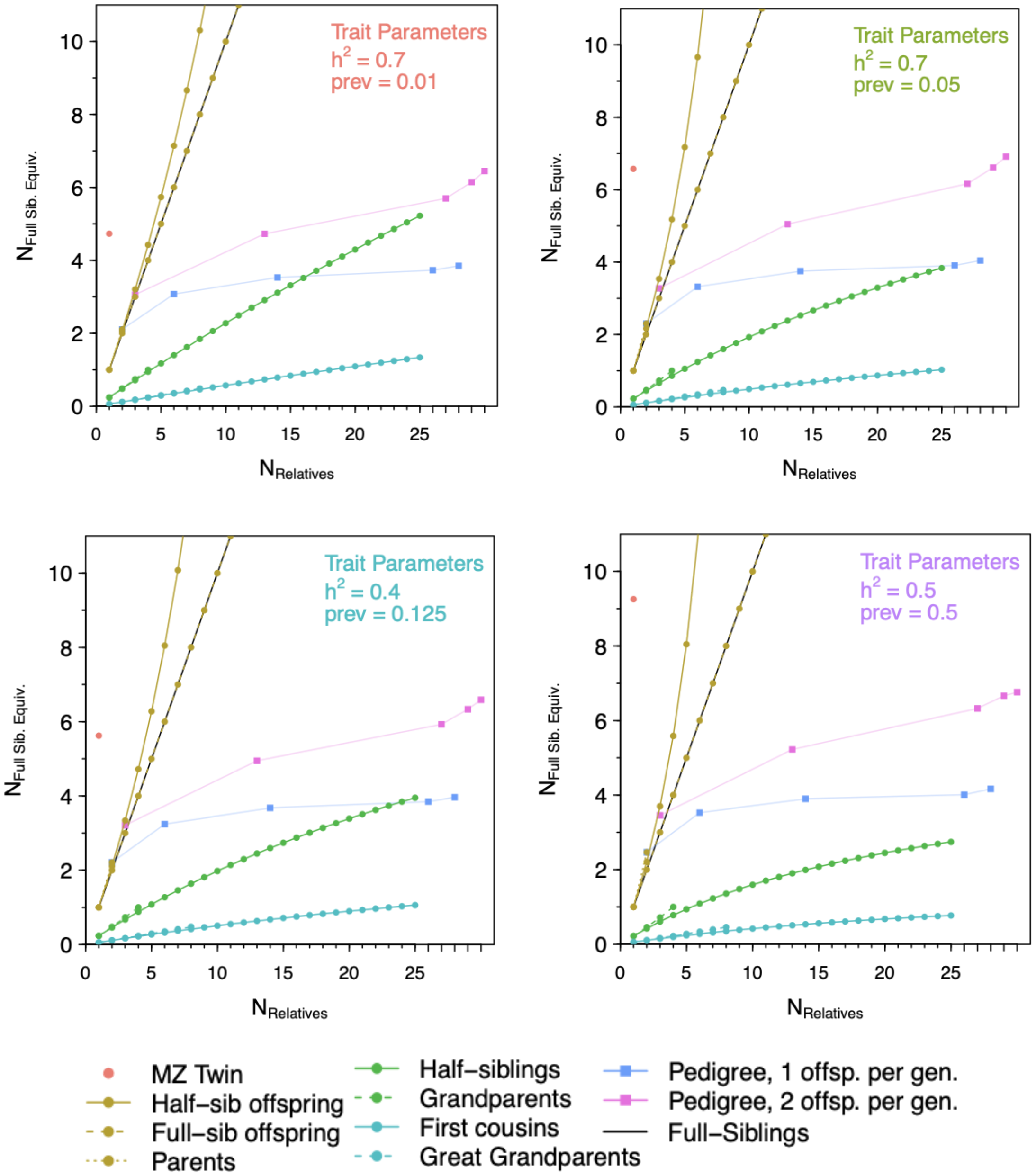


**Supplementary figure S17. Sibling equivalents for disease traits with different prevalence and heritability.**

The figure displays the number of fully siblings that would be required to obtain the same accuracy as obtained by 1 to 10 siblings, 1 or 2 parents, 1 to 5 half sibling offspring, one monozygotic twin, 1 to 4 grandparents (g.parents), 1 to 8 great grandparents, 1 to 16 cousins (cousins) and eight examples *n*-generational pedigrees with *n* varying from 1 (parents as founders) to 5 (gr. gr. gr. grandparents as founders) when all relatives mate and have either one child (child_pr_gen=1) or two children (child_pr_gen=2). The x-axis shows the number of relatives, while the y-axis shows the estimated number of sibling equivalents as estimated by the second order approximation. The four panel shows the relationship under different heritabilities (h2) and prevalence (K).

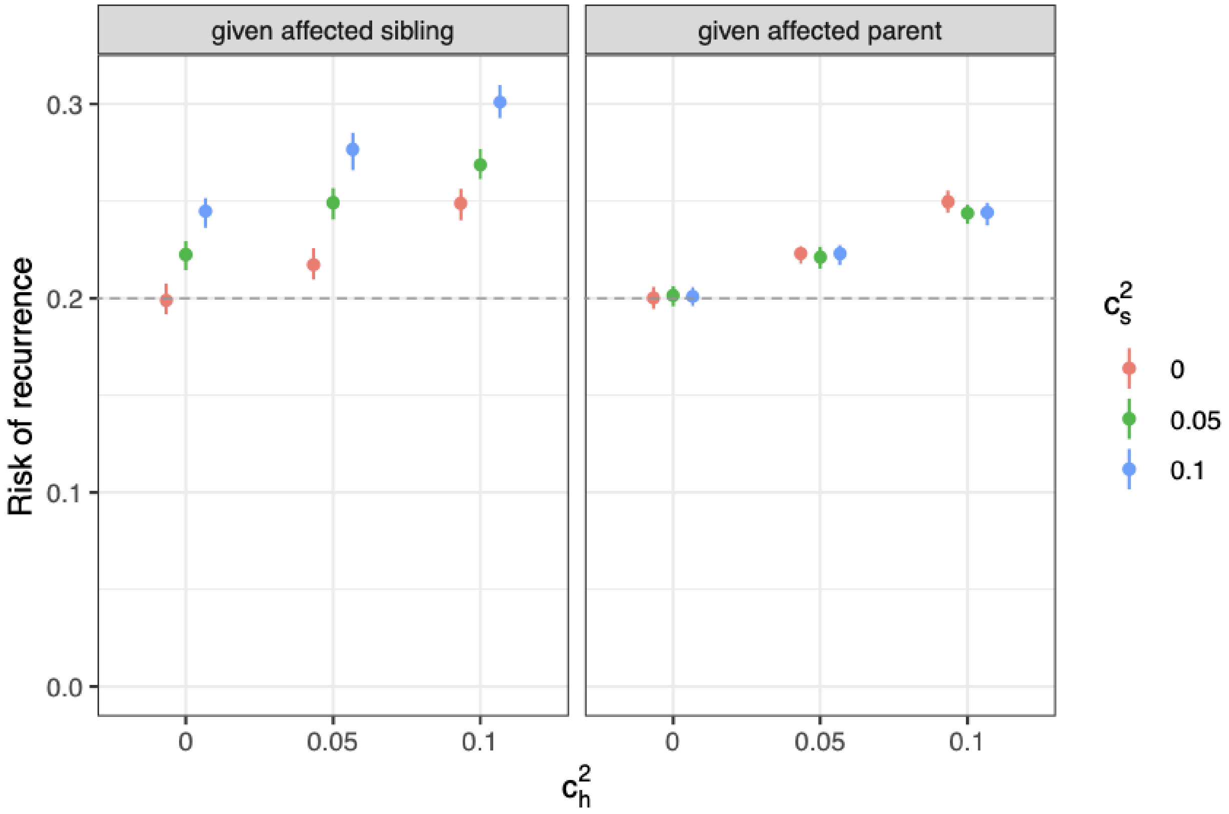


**Supplementary Figure S18. Impact of shared environment on risk of phenotypic recurrence.**
To test how effects of shared environment would impact on our results we performed simulation under a model $L_i = G_i + e_i$ where for a set of relatives $i, ..., n$ with $G \sim MVN([0,..., 0]^T, \Sigma_g)$ where $\sigma_{g,i,j} = cov(G_i, G_j) = r_{ij}h^2$ and $e \sim MVN([0,..., 0]^T, \Sigma_e)$, where

$$\sigma_{e,\ ij} = \mathrm{cov}\left(e_i, e_j\right) = \begin{cases} 1 - h^2 & i=j \\ c_h^2 + c_s^2 & \text{siblings} \\ c_h^2 & \text{parent offspring} \\ 0 & \text{otherwise} \end{cases}$$

Under this model we simulated 100,000 families of 14 individuals (a proband, a sibling, two parents, four grandparents, two avunculars, and four cousins) by sampling random draws from two 14-variate normal distributions with covariance matrices outlines above setting $h^2 = 0.4$, $c_h^2$ to 0, 0.05 or 0.1 and $c_s^2$ to 0, 0.1 or 0.2. We declared individuals as cases if $L_i = G_i + e_i > t = \Phi^{-1}(1 - K_{pop})$ with $K_{pop} = 0.125$

Here we display the expected impact of various levels of shared environmental effects within sibships ($c_s^2$) and/or within households ($c_h^2$) on the recurrence risk of the disease outcome. Under a model with no common environmental effects, we expect a recurrence risk associated with either an affected sibling or an affected parent to be ~0.2. Increasing levels of common environmental effects, increases the recurrence risk associated with an affected family member. Using family history information to predict outcomes in a proband with incorporate both additive genetics and aspects of the familial environment.

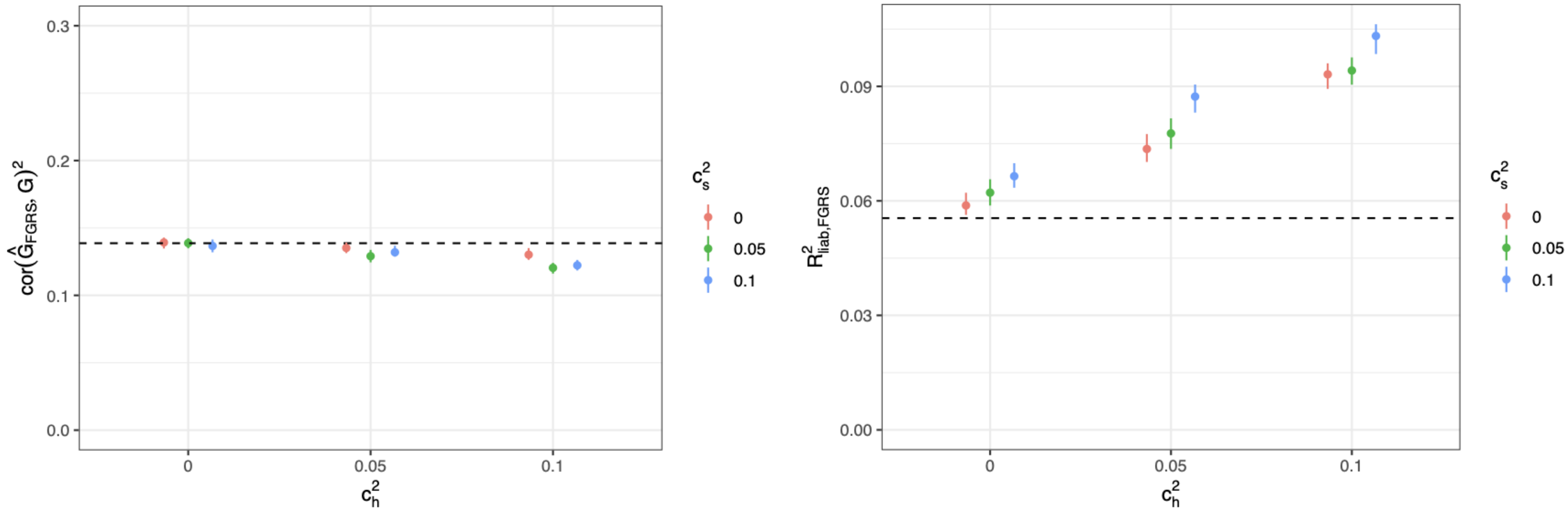


**Supplementary Figure S19. Impact of shared environment on squared accuracy and performance of PA-FGRS relative to expectations.**
To test how effects of shared environment would impact on our results we performed simulation under a model $L_i = G_i + e_i$ where for a set of relatives $i, ..., n$ with $G \sim MVN([0,..., 0]^T, \Sigma_g)$ where $\sigma_{g,i,j} = cov(G_i, G_j) = r_{ij}h^2$ and $e \sim MVN([0,..., 0]^T, \Sigma_e)$, where

$$\sigma_{e,\ ij} = \mathrm{cov}(e_i, e_j) = \begin{cases} 1 - h^2 & i = j \\ c_h^2 + c_s^2 & \text{siblings} \\ c_h^2 & \text{parent offspring} \\ 0 & \text{otherwise} \end{cases}$$

Under this model we simulated 100,000 families of 14 individuals (a proband, a sibling, two parents, four grandparents, two avunculars, and four cousins) by sampling random draws from two 14-variate normal distributions with covariance matrices outlines above setting $h^2 = 0.4$, $c_h^2$ to 0, 0.05 or 0.1 and $c_s^2$ to 0, 0.1 or 0.2. We declared individuals as cases if $L_i = G_i + e_i > t = \Phi^{-1}(1 - K_{pop})$ with $K_{pop} = 0.125$.

Here we display, via simulations (Supplementary Information), the observed squared accuracy (left panel) and performance (right panel) of PA-FGRS when various levels of shared environmental effects within sibships ($c_s^2$) and/or within households ($c_h^2$). In the left panel, we see that the squared correlation between $\hat{G}_{FGRS}$ and $G$ is modestly reduced by $c_h^2$ and $c_s^2$ and modestly lower than expected assuming $c_h^2 = c_s^2 = 0$. In the right panel, as $c_h^2$ or $c_s^2$ increases, the squared correlation between $\hat{G}_{FGRS}$ and $L$ increases and deviates from the performance expected assuming $c_h^2 = c_s^2 = 0$

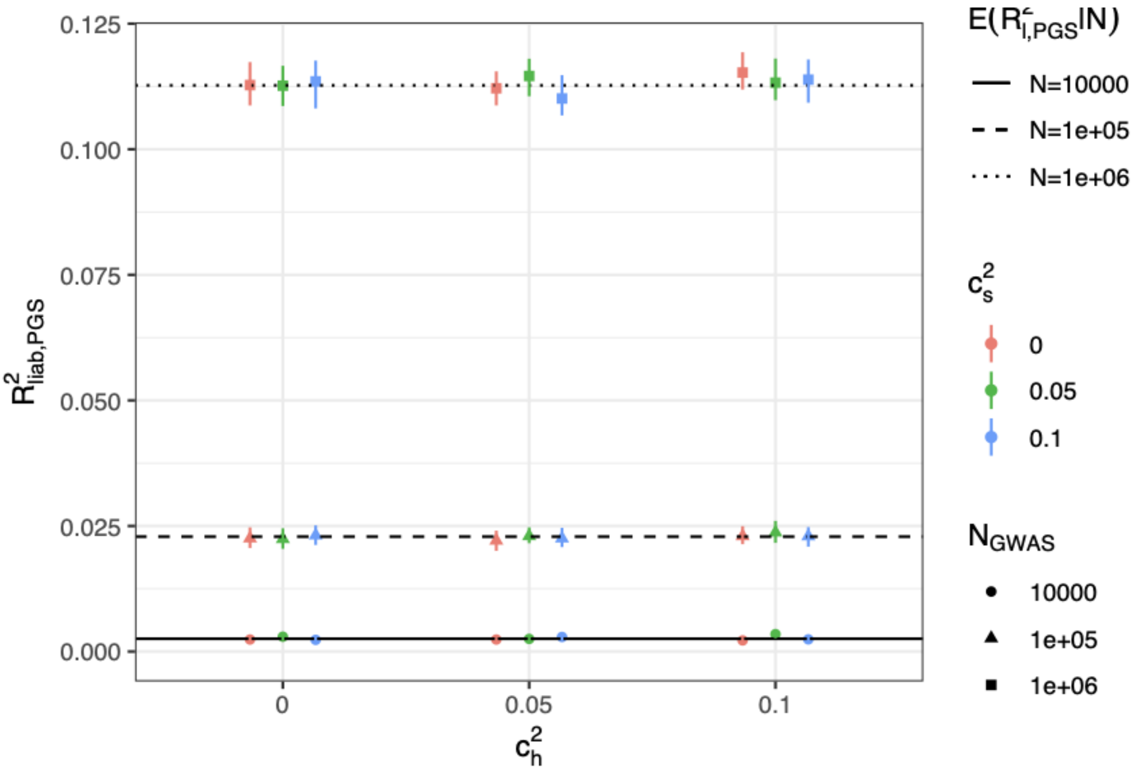


**Supplementary Figure S20. Impact of shared environment on performance of PGS relative to expectations.**

To test how effects of shared environment would impact on our results we performed simulation under a model $L_i = G_i + e_i$ where for a set of relatives $i, ..., n$ with $G \sim MVN([0,..., 0]^T, \Sigma_g)$ where $\sigma_{g,i,j} = cov(G_i, G_j) = r_{ij}h^2$ and $e \sim MVN([0,..., 0]^T, \Sigma_e)$, where

$$\sigma_{e,\,ij} = \mathrm{cov}(e_i, e_j) = \begin{cases} 1 - h^2 & i = j \\ c_h^2 + c_s^2 & \text{siblings} \\ c_h^2 & \text{parent offspring} \\ 0 & \text{otherwise} \end{cases}$$

Under this model we simulated 100,000 families of 14 individuals (a proband, a sibling, two parents, four grandparents, two avunculars, and four cousins) by sampling random draws from two 14-variate normal distributions with covariance matrices outlines above setting $h^2 = 0.4$, $c_h^2$ to 0, 0.05 or 0.1 and $c_s^2$ to 0, 0.1 or 0.2. We declared individuals as cases if $L_i = G_i + e_i > t = \Phi^{-1}(1 - K_{pop})$ with $K_{pop} = 0.125$. We simulated PGS for the probands as $\hat{G}_{i,PGS} = \sqrt{f}G_i + \epsilon$ where $\epsilon \sim N(0, 1 - f)$. Where $f$ is the fraction of $h^2$ expected to be explained by a PGS trained in a population study (i.e., the case proportion is equal to the prevalence) of $N$= 10,000, 100,000 or 1,000,000 samples.

Here we display, via simulations (Supplementary Information), the observed performance of PGS in scenarios with various levels of shared environmental effects on within sibships ($c_s^2$) and/or within households ($c_h^2$). In our simulations, $c_h^2$ and $c_s^2$ have no effect on the squared correlation between $\hat{G}_{PGS}$ and $L$.

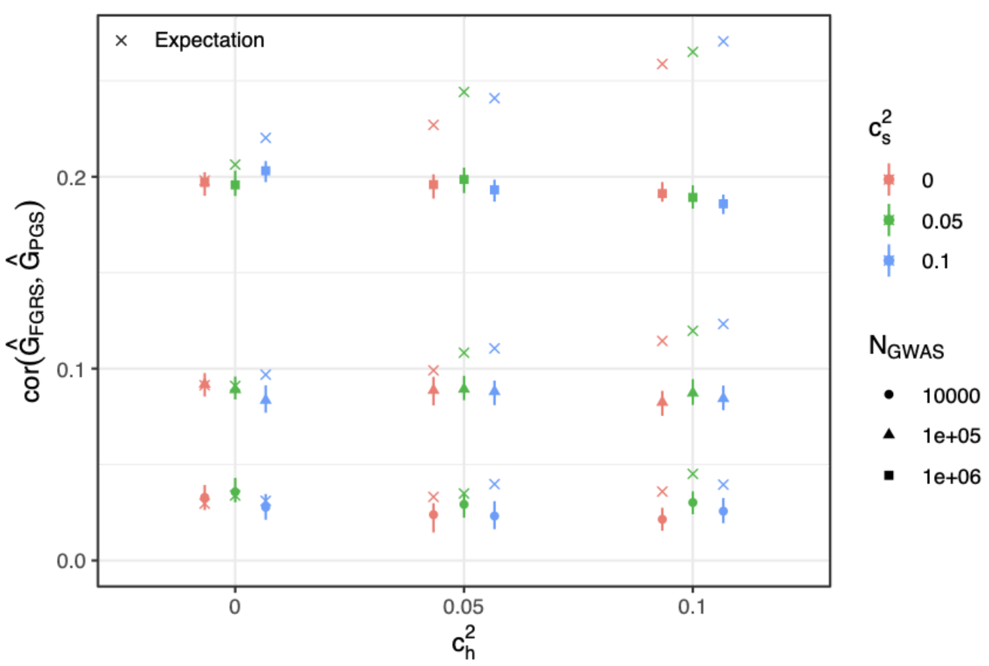


**Supplementary Figure S21. Impact of shared environment on correlation between PGS and PA-FGRS relative to expectations.**

To test how effects of shared environment would impact on our results we performed simulation under a model $L_i = G_i + e_i$ where for a set of relatives $i, ..., n$ with $G \sim MVN([0,..., 0]^T, \Sigma_g)$ where $\sigma_{g,i,j} = cov(G_i, G_j) = r_{ij}h^2$ and $e \sim MVN([0,..., 0]^T, \Sigma_e)$, where

$$\sigma_{e,\,ij} = \mathrm{cov}(e_i, e_j) = \begin{cases} 1 - h^2 & i = j \\ c_h^2 + c_s^2 & \text{siblings} \\ c_h^2 & \text{parent offspring} \\ 0 & \text{otherwise} \end{cases}$$

Under this model we simulated 100,000 families of 14 individuals (a proband, a sibling, two parents, four grandparents, two avunculars, and four cousins) by sampling random draws from two 14-variate normal distributions with covariance matrices outlines above setting $h^2 = 0.4$, $c_h^2$ to 0, 0.05 or 0.1 and $c_s^2$ to 0, 0.1 or 0.2. We declared individuals as cases if $L_i = G_i + e_i > t = \Phi^{-1}(1 - K_{pop})$ with $K_{pop} = 0.125$. We simulated PGS for the probands as $\hat{G}_{i,PGS} = \sqrt{f}G_i + \epsilon$ where $\epsilon \sim N(0, 1 - f)$. Where $f$ is the fraction of $h^2$ expected to be explained by a PGS trained in a population study (i.e., the case proportion is equal to the prevalence) of $N$= 10,000, 100,000 or 1,000,000 samples.

Here we display, via simulations, the observed (filled shapes) and expected (x symbols) correlation between PA-FGRS and PGS trained with different sample size GWAS, across scenarios with various levels of shared environmental effects on within sibships ($c_s^2$) and/or within households ($c_h^2$). In our simulations, $c_h^2$ and $c_s^2$ reduce slightly the correlation between $\hat{G}_{PGS}$ and $\hat{G}_{FGRS}$ (filled shapes), but the observed correlations are much lower than what would be expected from their marginal performances, assuming no familial effects (x symbols).

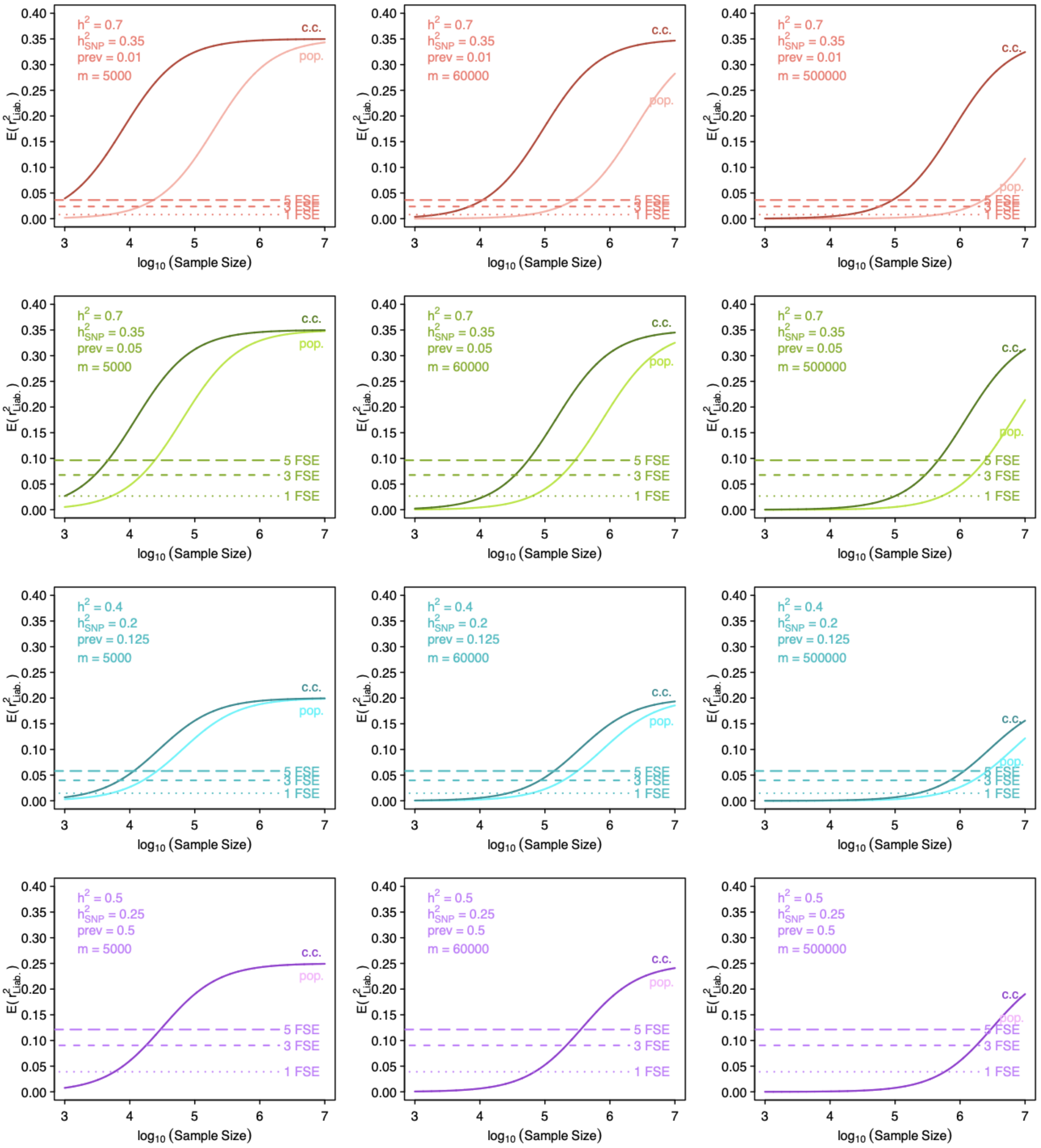


**Supplementary Figure S22. Choice of "M" and FGRS-PGS relationship.**

Assuming trait $h^2_{SNP,test} = h^2_{SNP,training} = 0.5\,h^2$ , $r_{G,test-training} = 1$ and the effective proportion of causal markers ($p$) is 1, we compare the expected performance of PGS to the expected performance of PA-FGRS with $N_{sibe}$=1,3, or 5 (dashed lines) under different choices of number of independent genetic factors (M). Compared to the usual choice of M= 60 000 decreasing M to 10 000 decreases the sample size required to obtain the sample size as PA-FGRS predictors. Whereas increasing M to 1 000 000 increases the required sample size. PA-FGRS, Pearson-Aitken Family Genetic Risk Scores; PGS, polygenic score; $h^2$, narrow-sense heritability; $h^2_{SNP}$, SNP heritability; $r_G$, genetic correlation; prev, lifetime prevalence; $N_{sibe}$, number of full sibling equivalents; c.c., case-control sampling; pop., population sampling.

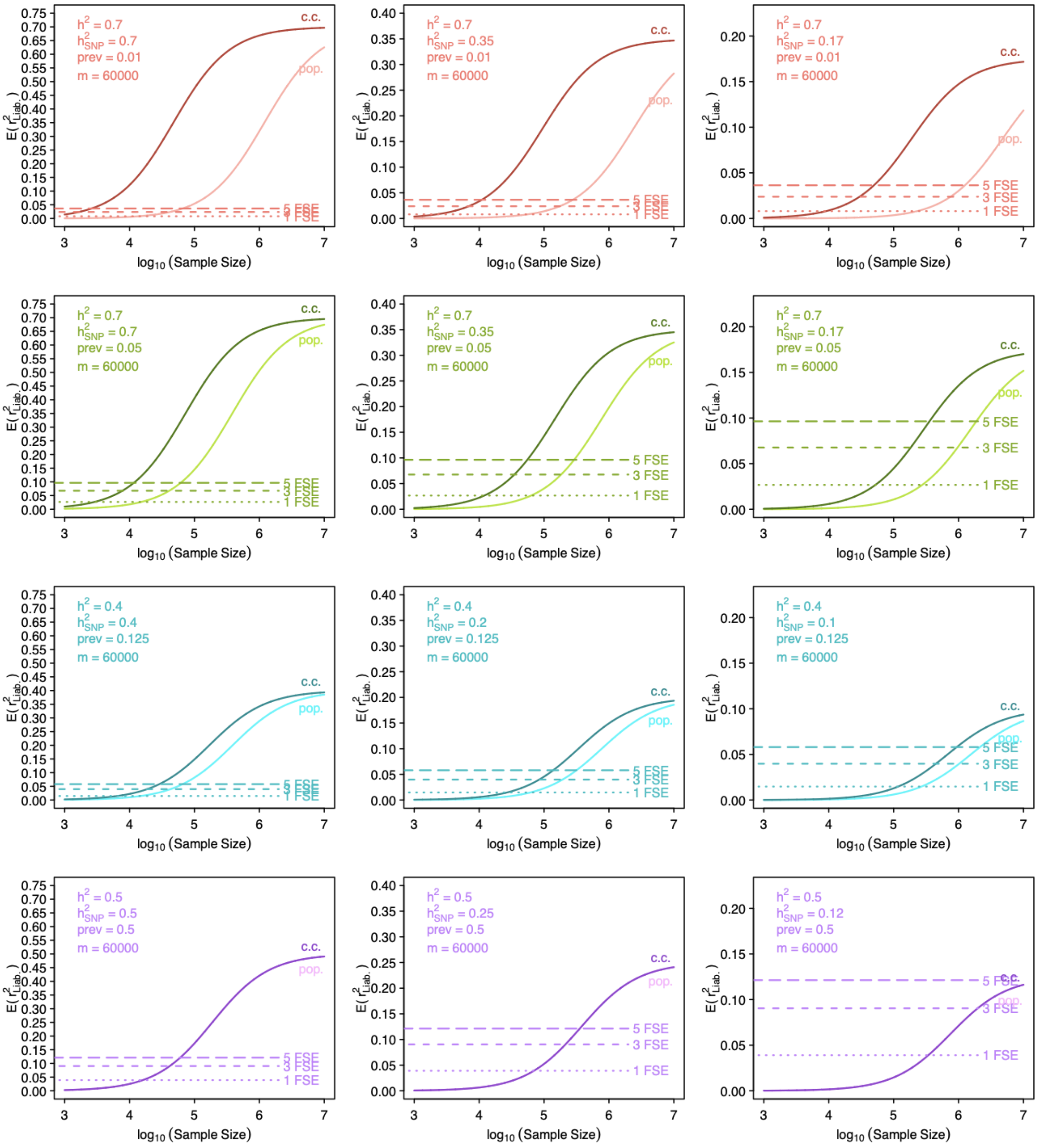


**Supplementary Figure S23. Choice of h2_snp / h2 and FGRS-PGS relationship**

Assuming trait $h^2_{SNP,test} = h^2_{SNP,training}$, $r_{G,test-training} = 1$ and the effective proportion of causal markers ($p$) is 1 and M= 60 000, we compare the expected performance of PGS to the expected performance of PA-FGRS with $N_{sibe}$=1,3, or 5 (dashed lines) under different choices of $h^2_{SNP}$. PA-FGRS, Pearson-Aitken Family Genetic Risk Scores; PGS, polygenic score; $h^2$, narrow-sense heritability; $h^2_{SNP}$, SNP heritability; $r_G$, genetic correlation; prev, lifetime prevalence; $N_{sibe}$, number of full sibling equivalents; c.c., case-control sampling; pop., population sampling.

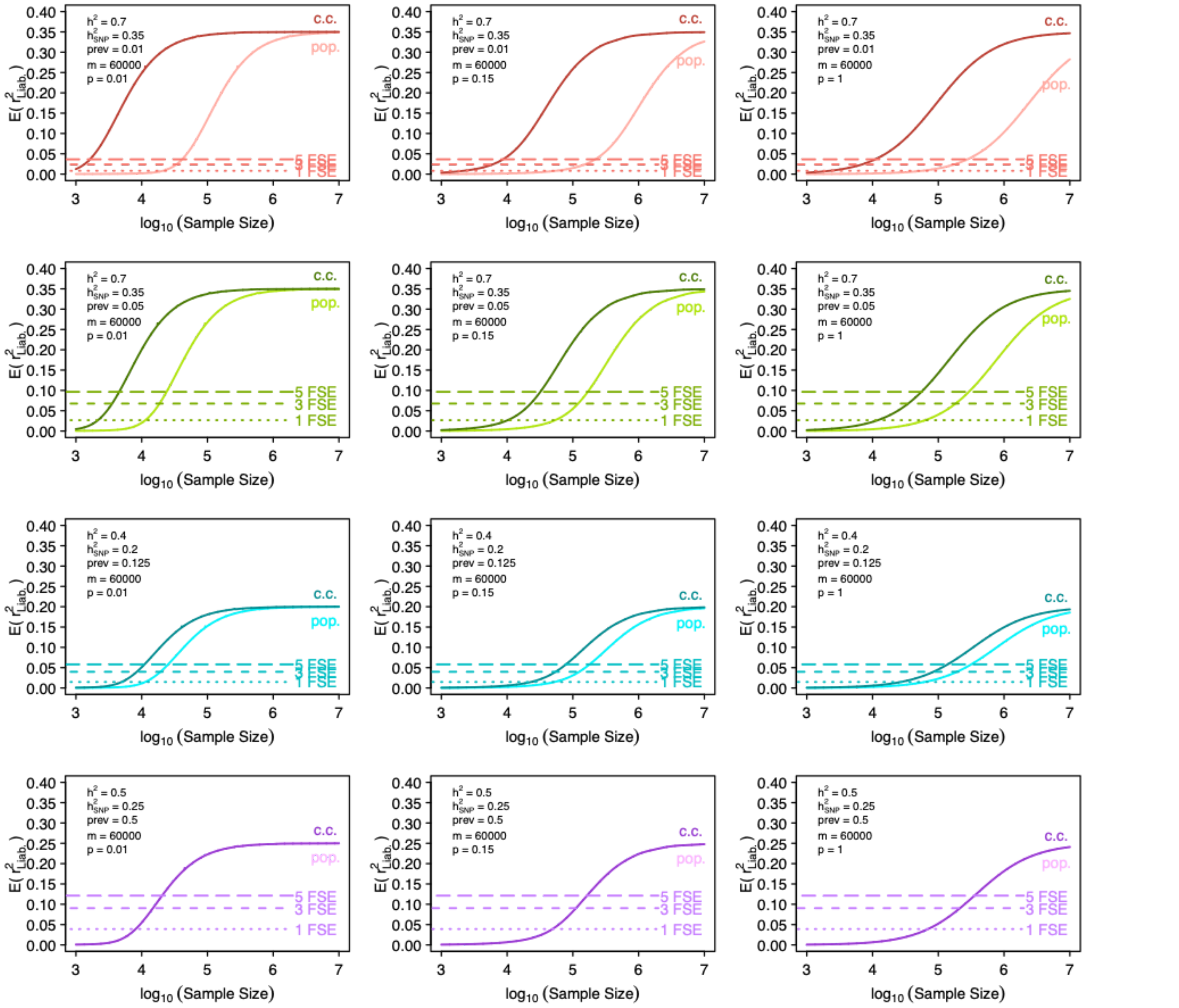


**Supplementary Figure S24. Choice of p and FGRS-PGS relationship.**

Assuming trait $h^2_{SNP,test} = h^2_{SNP,training}$ , $r_{G,test-training} = 1$, $h^2_{SNP} = 0.5\, h^2$, and M= 60 000, we compare the expected performance of PGS to the expected performance of PA-FGRS with $N_{sibe}$=1,3, or 5 (dashed lines) under different choices of *p*. Compared to our standard choice of p=1 (right column), decreasing *p* decreases the sample size required to obtain the sample size as PA-FGRS predictors (i.e., increases the efficiency of thee PGS). PA-FGRS, Pearson-Aitken Family Genetic Risk Scores; PGS, polygenic score; $h^2$, narrow-sense heritability; $h^2_{SNP}$, SNP heritability; $r_G$, genetic correlation; prev, lifetime prevalence; $N_{sibe}$, number of full sibling equivalents; c.c., case-control sampling; pop., population sampling.

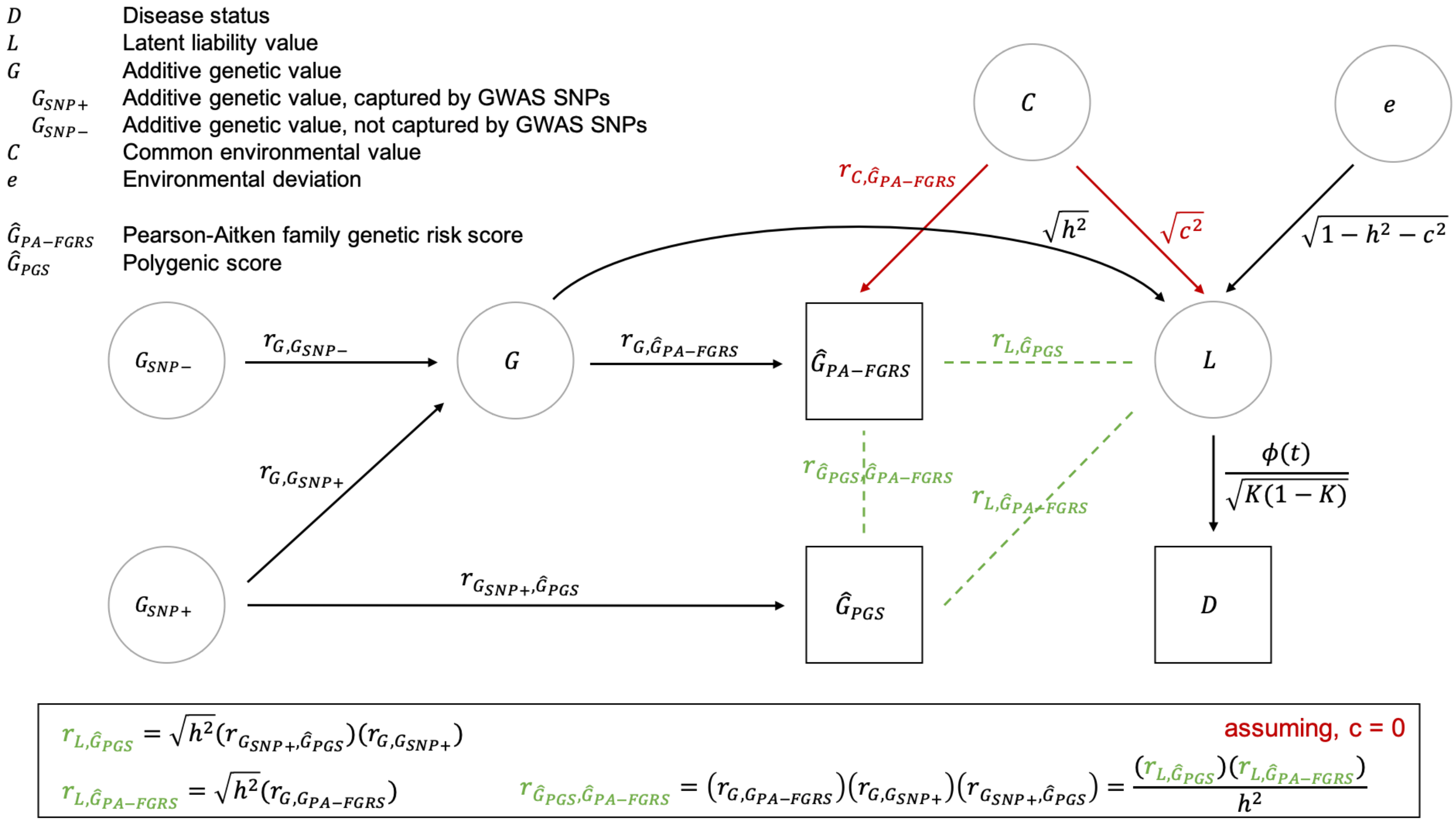


**Supplementary Figure S25. A diagram depicting our generative liability model and the implied relationships among PGS, PA-FGRS, and liability.**
The relationships of interest (green dashed lines) among PGS, PA-FGRS, and disease liability can be inferred from a path diagram that depicts our assumed generative liability model. PA-FGRS its expected relationships to PGS and liability are derived assuming no confounding by familial environment. In other sensitivity analyses we relax this assumptions to describe how these relationships are affected if this assumption is not valid.

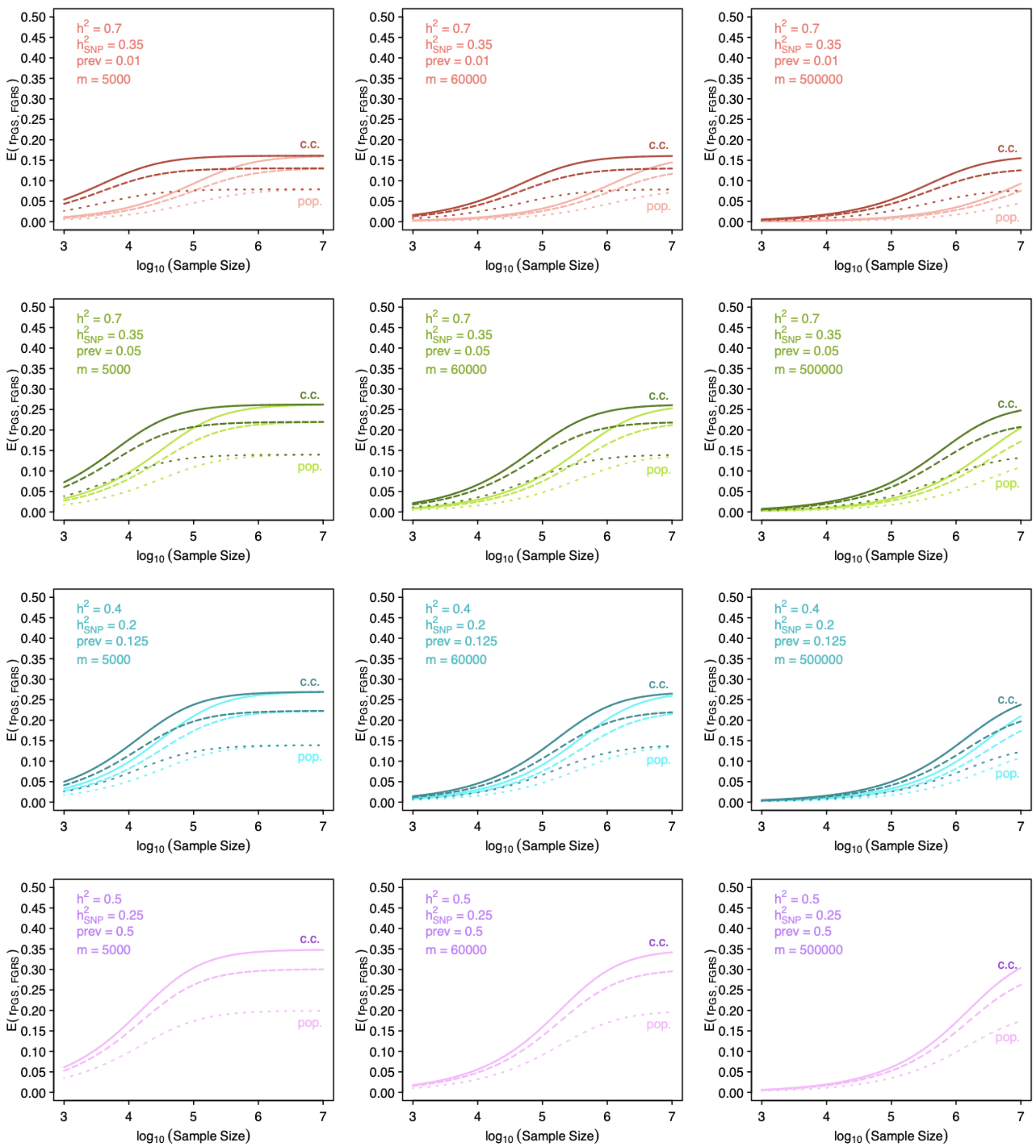


**Supplementary Figure S26. Choice of M in PGS-FGRS correlation**

Assuming trait $h^2_{SNP,test} = h^2_{SNP,training}$ , $r_{G,test-training} = 1$, $h^2_{SNP} = 0.5* h^2$, and p=1, we compare the expected correlation of a PGS to PA-FGRS with $N_{sibe}$=1,3, or 5 (dotted, dashed and solid lines) under different choices of $m$, the number of independent markers in a PGS that can explain the full $h^2_{SNP}$. PA-FGRS, Pearson-Aitken Family Genetic Risk Scores; PGS, polygenic score; $h^2$, narrow-sense heritability; $h^2_{SNP}$, SNP heritability; $r_G$, genetic correlation; prev, lifetime prevalence; $N_{sibe}$, number of full sibling equivalents; c.c., case-control sampling; pop., population sampling.

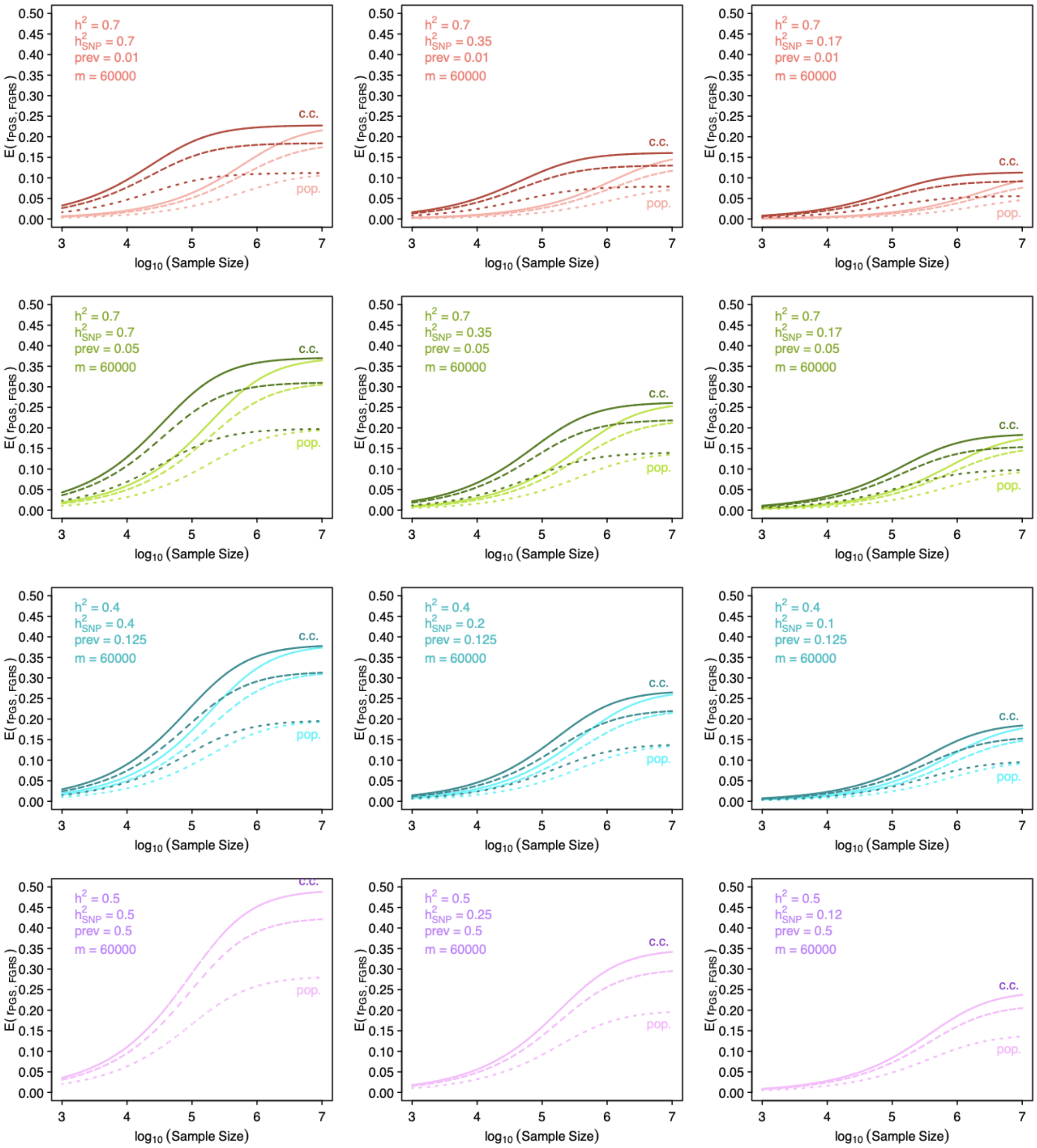


**Supplementary Figure S27. Choice of $h^2_{SNP}$ in PGS-FGRS correlation**

Assuming trait $h^2_{SNP,test} = h^2_{SNP,training}$ , $r_{G,test-training} = 1$, m=60,000, and p=1, we compare the expected correlation of a PGS to PA-FGRS with $N_{sibe}$=1,3, or 5 (dotted, dashed and solid lines) under different choices of $h^2_{SNP}$ defined either 100%, 50%, or 25% of the $h^2$. Increasing the proportion of $h^2$ covered by $h^2_{SNP}$ increases the efficiency of the PGS and this increases the expected correlation across the range of training sample sizes. Importantly, even as the PGS efficiency increases, the PA-FGRs efficiency stays the same which limits the maximum correlation of the two instruments. PA-FGRS, Pearson-Aitken Family Genetic Risk Scores; PGS, polygenic score; $h^2$, narrow-sense heritability; $h^2_{SNP}$, SNP heritability; $r_G$, genetic correlation; prev, lifetime prevalence; $N_{sibe}$, number of full sibling equivalents; c.c., case-control sampling; pop., population sampling.

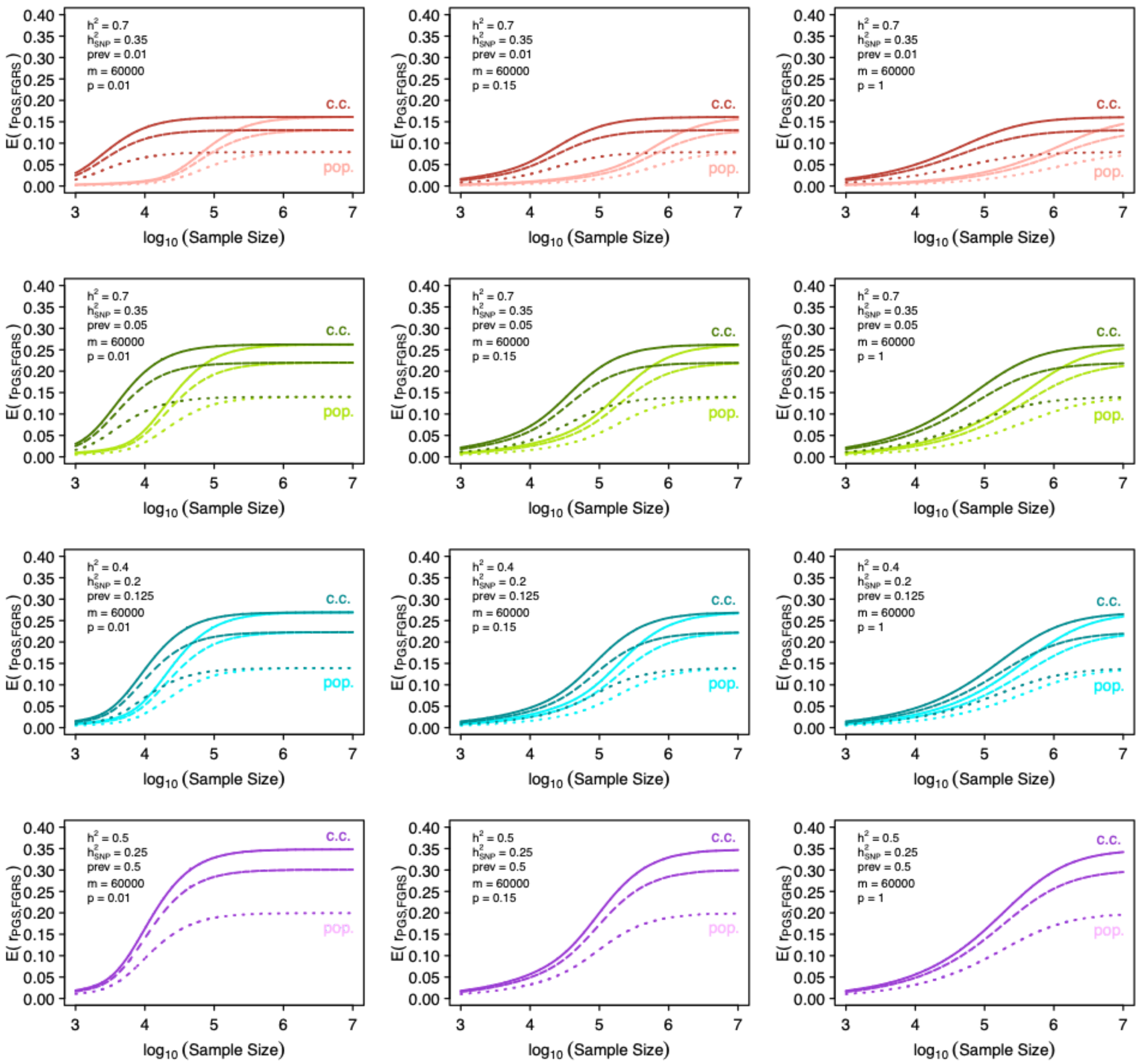


**Supplementary Figure S28. Choice of p in PGS, PA-FGRS correlation**

Assuming trait $h^2_{SNP,test} = h^2_{SNP,training}$ , $r_{G,test-training} = 1$, m=60,000, and $h^2_{SNP} = 0.5 * h^2$, we compare the expected correlation of a PGS to PA-FGRS with $N_{sibe}$=1,3, or 5 (dotted, dashed and solid lines) under different choices of p, the polygenicity factor among the m markers. Decreasing p, the proportion of m with an effect, increases the efficiency of the PGS and this increases the expected correlation across the range of training sample sizes. Importantly, even as the PGS efficiency increases, the PA-FGRs efficiency stays the same which limits the maximum correlation of the two instruments. PA-FGRS, Pearson-Aitken Family Genetic Risk Scores; PGS, polygenic score; $h^2$, narrow-sense heritability; $h^2_{SNP}$, SNP heritability; $r_G$, genetic correlation; prev, lifetime prevalence; $N_{sibe}$, number of full sibling equivalents; c.c., case-control sampling; pop., population sampling.

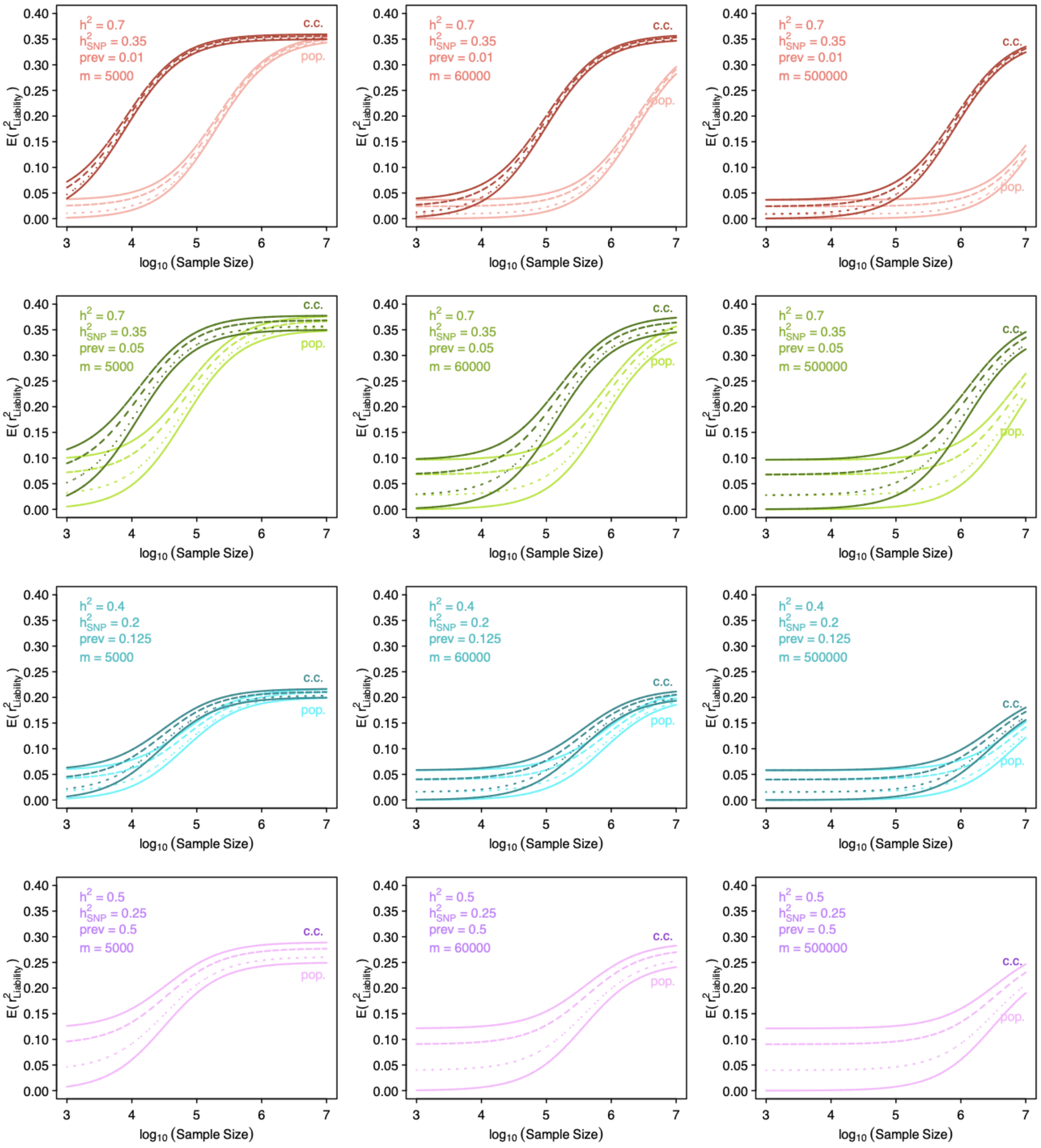


**Supplementary Figure S29. Choice of M in PGS-FGRS joint prediction**

Assuming trait $h^2_{SNP,test} = h^2_{SNP,training}$ , $r_{G,test-training} = 1$, $h^2_{SNP} = 0.5* h^2$, and p=1, we compare the expected performance of a PGS (lower solid line) and a joint predictor of PGS and PA-FGRS with $N_{sibe}$=1,3, or 5 (dotted, dashed and upper solid lines) under different choices of *m*, the number of independent markers in a PGS that can explain the full $h^2_{SNP}$. PA-FGRS, Pearson-Aitken Family Genetic Risk Scores; PGS, polygenic score; $h^2$, narrow-sense heritability; $h^2_{SNP}$, SNP heritability; $r_G$, genetic correlation; prev, lifetime prevalence; $N_{sibe}$, number of full sibling equivalents; c.c., case-control sampling; pop., population sampling.

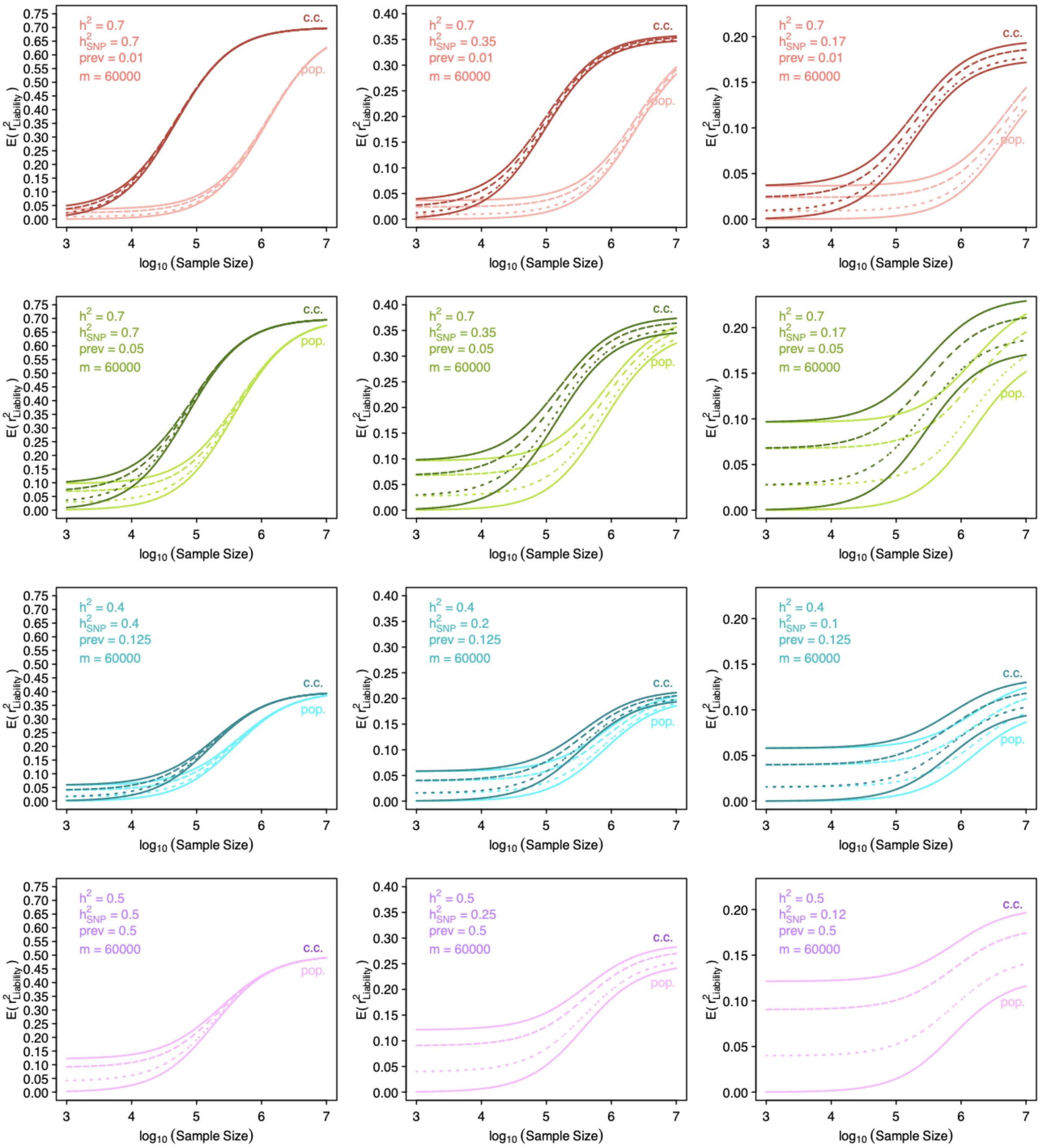


**Supplementary Figure S30. Choice of $h^2_{SNP}$ in PGS-FGRS joint prediction**

Assuming trait $h^2_{SNP,test} = h^2_{SNP,training}$, $r_{G,test-training} = 1$, m=60,000, and p=1, we compare the expected performance of a PGS (lower solid line) and a joint predictor of PGS and PA-FGRS with $N_{sibe}$=1,3, or 5 (dotted, dashed and upper solid lines) under different choices of $h^2_{SNP}$ defined either 100%, 50%, or 25% of the $h^2$. PA-FGRS, Pearson-Aitken Family Genetic Risk Scores; PGS, polygenic score; $h^2$, narrow-sense heritability; $h^2_{SNP}$, SNP heritability; $r_G$, genetic correlation; prev, lifetime prevalence; $N_{sibe}$, number of full sibling equivalents; c.c., case-control sampling; pop., population sampling.

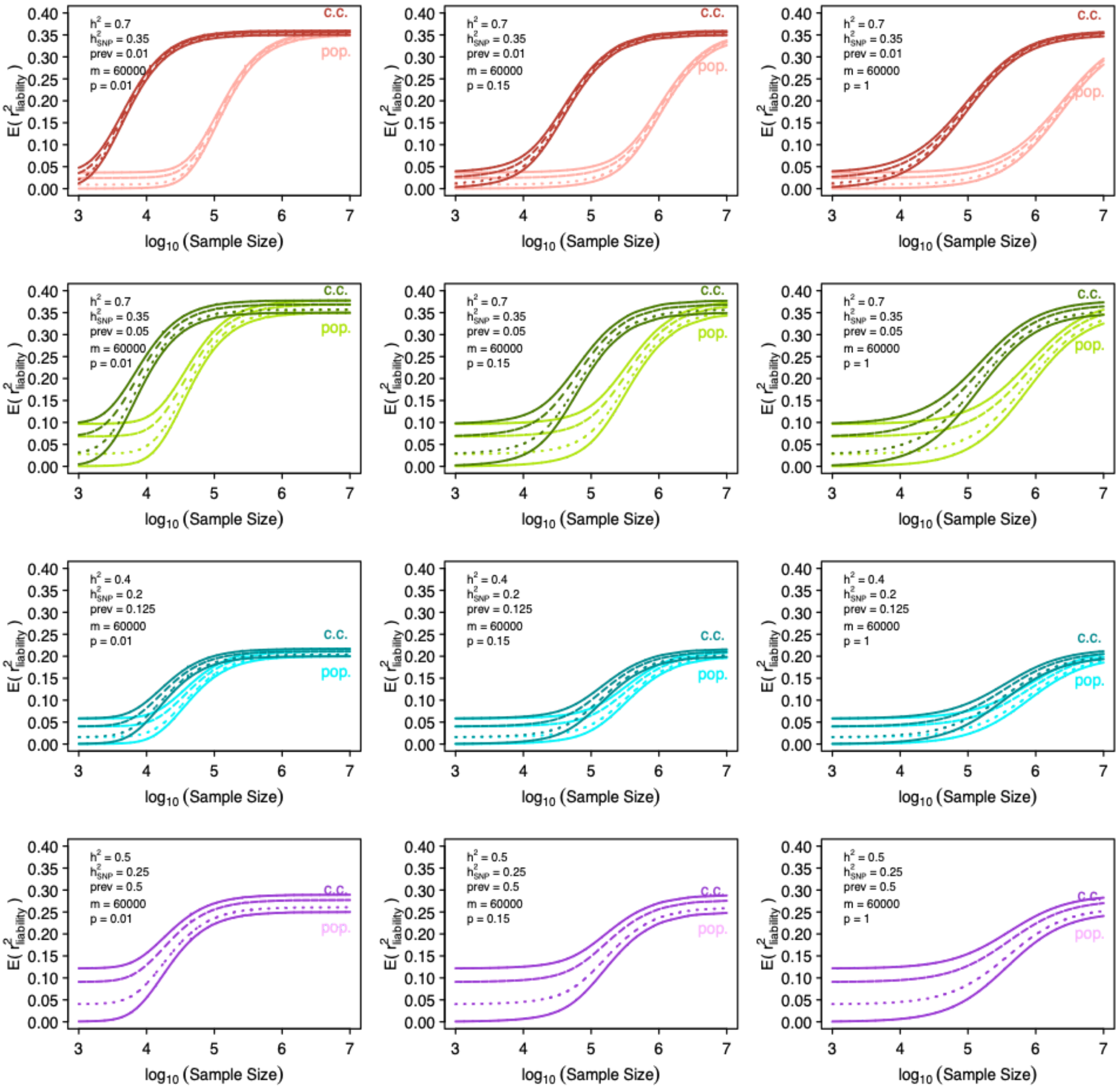


**Supplementary Figure S31. Choice of p in PGS-FGRS joint prediction**

Assuming trait $h^2_{SNP,test} = h^2_{SNP,training}$ , $r_{G,test-training} = 1$, m=60,000, and $h^2_{SNP} = 0.5h^2$, we compare the expected performance of a PGS (lower solid line) and a joint predictor of PGS and PA-FGRS with $N_{sibe}$=1,3, or 5 (dotted, dashed and upper solid lines) under different choices of p. PA-FGRS, Pearson-Aitken Family Genetic Risk Scores; PGS, polygenic score; $h^2$, narrow-sense heritability; $h^2_{SNP}$, SNP heritability; $r_G$, genetic correlation; prev, lifetime prevalence; $N_{sibe}$, number of full sibling equivalents; c.c., case-control sampling; pop., population sampling.
