## Supplementary Information for "The relationship between genotype- and phenotype-based estimates of genetic liability to psychiatric disorders, in practice and in theory"

S.1 A note on liability transformations of variance explained 3
S.2 Defining accuracy, reliability, and liability variance explained 5
S.3 The accuracy of PA-FGRS 7
S.4 The asymptotic accuracy of Glinear from one class of relatives 11
S.5 Accuracy in an arbitrarily structured pedigree 12
S.6 Accuracy of linear predictor under censoring 14
S.7 The expected accuracy of PGS 14
S.8 The expected accuracy of shrinkage PGS 15
S.9 Expected correlation of PGS and PA-FGRS assuming conditional independence 18
S.10 On the asymptotic difference between linear predictors and PA-FGRS 18
S.11 Exact estimator for PA-FGRS accuracy 23
Supplementary References: 25

**Definitions:**

| | |
|---|---|
| $G$ | random variable of true genetic liability of the probands |
| $L$ | random variable of true total liability of the probands |
| $\hat{G}_{PA-FGRS}$ | random variable of estimated genetic liability. Estimated from phenotypes in relatives. |
| $\hat{G}_{PGS}$ | random variable of estimated genetic liability. Estimated from genotypes. |
| $h_l^2$ | heritability on the liability scale. |
| $K_{pop}$ | prevalence of the disease in the population |
| $t$ | liability threshold |
| $\mathbf{g}$ | vector of of true genetic liabilities of the probands |
| $\mathbf{L}$ | an $(m \times n)$ matrix with the liabilities of the $n$ relatives of each of the $m$ probands |
| $\mathbf{D}$ | an $(m \times n)$ matrix with the disease status of the $n$ relatives of each of the $m$ probands<br>(i.e. $d_{ij} = 1, l_{ij} > t; 0, l_{ij} < t$ ) |
| $\mathbf{R}$ | an $(m \times m)$ genetic relatedness matrix. |
| $\mathbf{P}$ | an $(n - 1 \times n - 1)$ phenotypic covariance matrix of the relatives.<br>$p_{ij} = \mathrm{Cov}(d_{,i}, d_{,j}) \{ \rho, i \neq j ; K(1 - K), i = j$ |
| $\mathbf{c}$ | an $(n - 1)$ vector denoting the covariance between the genetic value of the proband and the liability of each of $n - 1$ relatives (i.e. $c_i = \mathrm{Cov}(\mathbf{g}, \mathbf{L}_{,i})$ ) |

#### S.1 A note on liability transformations of variance explained

In order to model expectations and compare magnitudes across instruments and studies, genetic effects must be estimated accurately and on an objective scale. In human genetics, this means reporting effects on the liability scale[37,38], typically by mathematical transformations of effects estimated on other scales (e.g., from linear or logistic regression)[38]. These common transformations are based on assumptions of one-stage case-control sampling (i.e., *complete* ascertainment) and well-behaved (e.g., Gaussian, exogenous error) genetic instruments like PGS, but have not been shown to be valid under two-stage case-control sampling typical in molecular genetics studies (e.g., sampling *unrelated* cases; akin to observing relatives conditional on their relationship to an affected proband as common in twin register studies[39]) and less well-behaved (i.e., non-Gaussian, endogenous error) genetic instruments (e.g., FH, PA-FGRS). In Supplementary Figure S1, we observed qualitatively different estimates of variance explained by our instruments, depending on transformation. In our simulations (described in detail below) we observe that proposed transformations of variance explained by a linear model (observed scale) to the liability scale are biased for FH and PA-FGRS, but not for PGS (Supplementary Figures 2A-C; Supplementary Table S3). This resulted in overestimating the variance explained by more than 500% in some scenarios and produced bias in all scenarios where the trait prevalence was less than 50% (Supplementary Figures 2A-C; Supplementary Table S3). Directly estimating variance explained on the liability scale using a weighted probit regression as described by Lee et al[38] reduced this bias under population sampling (Supplementary Figures 2D; Supplementary Table S3) and complete case-control sampling (Supplementary Figures 2E; Supplementary Table S3), but introduced a downward bias of nearly 40% when unrelated case-control sampling was applied (Supplementary Figures 2F; Supplementary Table S3). A weighted probit regression with two-stage sampling weights to account for this second selection, namely, removing all but one member of a relative cluster, produces the most reliable estimates of variance explained on the liability scale, but could not eliminate completely small downward bias (5–15%) for rarer traits (Supplementary Figures 2G-I; Supplementary Table S3). Estimating objective, liability scale variance components when sampling is complex (i.e., multi-stage) and instruments are not well-behaved (e.g., FH, PA-FGRS, or other liability scores) is difficult and direct approaches incorporating appropriate sampling weights producing the most reliable estimates.

Lee et al.[1] proposed several ways to estimate liability variance explained ($R_l^2$) including a transformation of the variance explained in a linear regression $R_{obs}^2$ onto the liability scale, $R_{l_{cc}}^2$, given the population prevalence of the trait and case sampling proportion. Briefly, Dempster and Lerner[4] showed that for population studies, where the proportion of cases, $w$, corresponds to the prevalence, $K$, $R_l^2$ can be estimated as $R_l^2 = R_{obs}^2 \frac{K(1-K)}{\phi(t)^2}$, where $\phi(t)$ is the density of the standard normal distribution af the threshold, $t$. Lee et al.[1] proposed an extension for case control studies ( $w > K$), such that:

$$R_{l_{cc}}^2 = \frac{R_{obs}^2 C}{1+R_{obs}^2 \theta C}, \qquad \text{(Eq. S1)}$$

where $C = \frac{K(1-K)}{\phi(t)^2} \frac{K(1-K)}{w(1-w)}$ and $\theta = \frac{\phi(t)(w-K)}{K(1-K)} \left( \frac{\phi(t)(w-K)}{K(1-K)} - t \right)$

Alternatively, they proposed that $R_l^2$ can be estimated using a weighted probit model with the weights $\frac{K(1-w)}{w(1-K)}$ for cases and 1 for controls. To see this, let $S$ denote an indicator variable of being in the case control set, $Y$ denote the case status, $w$ is the proportion of cases in the study, and $K$ the prevalence in the population.

By Bayes theorem we have:

$$P(S = 1|Y = 0) = \frac{P(Y=0|S=1)P(S=1)}{P(Y=0)} = \frac{(1-w)P(S=1)}{1-K}$$

and similarly,

$$P(S = 1|Y = 1) = \frac{P(Y=1|S=1)P(S=1)}{P(Y=1)} = \frac{wP(S=1)}{K}$$

Thus the ratio between $P(S = 1|Y = 1)$ and $P(S = 1|Y = 0)$ is:

$$\frac{P(S=1|Y=1)}{P(S=1|Y=0)} = \frac{w(1-K)}{K(1-w)}$$

rearranging gives us:

$$P(S = 1|Y = 1) = \frac{w(1-K)}{K(1-w)} P(S = 1|Y = 0) \qquad \text{(Eq. S2)}$$

Lee et al.[1] used simulated data to show this was robust when the predictor was a Polygenic Score (PGS), however, the equations were not validated for phenotype-based genetic instruments such as family history indicators (FH) or family genetic risk scores (FGRS), which can have very different distributional properties.

In Supplementary Figure S1, we show how the choice of model and scale-transformation impacts on our real results such that the estimated and transformed variance in liability are not consistent across models, particularly for the FH and PA-FGRS estimates. This is critical because, e.g., the transformation of the Pearson correlation used by Hujoel et al.[2] and Lee et al[1] can provide nonsensical values ($R_l^2 > 1$) if applied when $R_{obs}^2 > \frac{\phi(t)^2}{K(1-K)}$.

An additional issue that can arise then estimating $R_l^2$ is if relatives have been excluded from a case control study. To see this, we can can introduce a second selection $S_2$ which forms a subset of $S$ (i.e. $P(S_{2,i} = 0|S_i = 0)$). If $S_2$ is independent of both outcome and predictors, it can be ignored, but in other cases, including the case of 'relative pruning' to obtain a case control study of unrelated individuals, it may impact on our estimate of variance explained. For this we introduce a second set of weights. Let $Z_i = 1$ denote that the *i*th individual has a relative in *S*, and that only one relative per family is included in $S_2$, the the conditional probability of being sampled in $S_2$ if sampled in $S$ is:

$$P(S_{2,i} = 1|S_i = 1, Z_i = 1) = \frac{\sum_i^n S_{2_i} Z_i}{\sum_i^n Z_i} = \mu_{(S_2, Z=1)}$$

and

$$P(S_{2,i} = 1|S_i = 1,\ Z_i = 0) = 1 \qquad \text{(Eq.S3)}$$

thus our two stage weights are the inverse of product of the case control probabilities (Eq S2), and the relatedness probabilities (Eq.S3):

$$w_{2stage,i} = \begin{cases} 1 & \text{if } Y_i = 0 \text{ and } Z_i = 0 \\ \frac{K(1-w)}{w(1-K)} & \text{if } Y_i = 1 \text{ and } Z_i = 0 \\ \mu_{(S_2|Z=1)} & \text{if } Y_i = 0 \text{ and } Z_i = 1 \\ \frac{K(1-w)}{w(1-K)\,\mu_{(S_2|Z=1)}} & \text{if } Y_i = 1 \text{ and } Z_i = 1 \end{cases} \qquad \text{(Eq.S4)}$$

To investigate the properties of the two transformation approaches suggested by Lee et al[1] and our two-stage weighting, when estimating $R_l^2$ for PA-FGRS, FH and PGS, we performed a simulation experiment. First, we simulated a simplified population consisting of 500,000 families each with three children and two parents. For each family, the liabilities were drawn from a multivariate normal distribution, $MVN([0, 0, 0, 0, 0]^T, \Sigma_l)$, where

$$\Sigma_g = \begin{bmatrix} h^2 & 0.5h^2 & 0.5h^2 & 0.5h^2 & 0.5h^2 \\ 0.5h^2 & h^2 & 0.5h^2 & 0.5h^2 & 0.5h^2 \\ 0.5h^2 & 0.5h^2 & h^2 & 0.5h^2 & 0.5h^2 \\ 0.5h^2 & 0.5h^2 & 0.5h^2 & h^2 & 0 \\ 0.5h^2 & 0.5h^2 & 0.5h^2 & 0 & h^2 \end{bmatrix}$$

We defined cases if $L_i = G_i + e_i > t$ and controls otherwise, where $t = \Phi^{-1}(1 - K)$. We investigated three different scenarios: (1) an *unrelated population sample* where we randomly selected one child from each family (e.g. 5K cases and 495K controls if K=0.01), (2) a *complete case-control sample* where we sampled all the cases and the same number of controls (e.g. 15K cases and 15K controls for K=0.01), or (3) an *unrelated case control sample* where only one child was retained from each family (e.g. ~14K cases and ~15K controls for K=0.01). We computed FH variables and PA-FGRS (see Online Methods and below) from the case status of the four relatives and we generated PGS-like predictor generated as the sum of the true genetic value noise sampled from $MVN([0, 0, 0, 0, 0]^T, \Sigma_\epsilon)$ where:

$$\Sigma_\varepsilon = \begin{bmatrix} 1 & 0.5 & 0.5 & 0.5 & 0.5 \\ 0.5 & 1 & 0.5 & 0.5 & 0.5 \\ 0.5 & 0.5 & 1 & 0.5 & 0.5 \\ 0.5 & 0.5 & 0.5 & 1 & 0 \\ 0.5 & 0.5 & 0.5 & 0 & 1 \end{bmatrix}$$

We then compared the true $R_l^2$ to the estimated and transformed $\hat{R}_l^2$ using the transformations ( $R_{l_{cc}}^2$ and $R_{l_{probit}}^2$ ) mentioned by Lee et al.[1] , and our proposed two-stage weighted extension $R_{l_{probit,2stage}}^2$ . This was repeated for four classes of traits and 10 times for each set of parameters. The mean and standard error of the estimates are reported in Supplementary Figure S2 and Supplementary Table S4.

#### S.2 Defining accuracy, reliability, and performance

We use the term accuracy to refer the correlation between an estimated genetic value, $\hat{G}$, and the true genetic value, $G$, and reliability to refer to the square of the accuracy.

$$accuracy = \mathrm{corr}\left(\hat{G}, G\right)$$
$$reliability = \mathrm{corr}\left(\hat{G}, G\right)^2$$

We define the performance to be the liability variance explained, $R_l^2$, or the squared correlation between the *total* liability, $L$, and the estimated genetic value which we can write as :

$$R_l^2 = \mathrm{corr}\left(\hat{G}, L\right)^2 = \frac{\mathrm{Cov}\left(\hat{G},L\right)^2}{\mathrm{Var}\left(\hat{G}\right)\mathrm{Var}(L)} = \frac{\mathrm{Cov}\left(\hat{G},G\right)^2}{\mathrm{Var}\left(\hat{G}\right)} = \mathrm{corr}\left(\hat{G}, G\right)^2 \mathrm{Var}(G) = \mathrm{corr}\left(\hat{G}, G\right)^2 h_l^2$$

This reminds us that the liability scale heritability, $h_l^2$, and the reliability of our genetic value estimate will, under an additive polygenic liability model, describe the asymptote for $R_l^2$, which will be equal to $h_l^2$ when there is no error in $\hat{G}$ (i.e., $\hat{G} = G$). While $R_l^2 = h_l^2$ can be obtained for monogenic traits and from phenotype information on an identical twins for perfectly heritable phenotypes, $\hat{h}_l^2$ is only a theoretical limit of the maximal achievable value of $R_l^2$ and, as we show, is typically unachievable in practice. In this paper we do not consider the performance of binary classifiers based on the PGS or FGRS (such as sensitivity, specificity, or AUC), but note that these are expected to improve monotonically with accuracy[5].

#### S.3 The accuracy of PA-FGRS

The expected accuracy of PA-FGRS has not previously been described. However, estimated breeding values (EBV)[6] for quantitative traits, i.e., *linear* combinations of phenotypes of proband relatives, are well studied in animal breeding and can be thought of as achieving the same goal as PA-FGRS in the hypothetical scenario where latent liability was directly observable. We can find guidance by considering $(y_{ij} - \mu_{ij})$ as the residual phenotypic value of the $j$th relative of individual $i$ after subtraction of fixed effects ($\mu_{ij}$) and define a simple EBV as the weighted sum,

$$\hat{G}_{i,EBV} = b_{1,EBV}(y_{i1} - \mu_{i1}) + \cdots + b_{n,EBV}(y_{in} - \mu_{in})$$

Where $b_{j,EBV}$ are the weights of the measurement on the $j$th relative, defined as $b_{EBV} = \mathrm{Cov}(y_1, \cdots, y_n)^{-1}\mathrm{Cov}([y_1, \cdots, y_n], y_i)$. For simplicity, we can consider the expected accuracy of $\hat{G}_{i,EBV}$ when all relatives are equally related to the index person and to each other, e.g., when considering a large sibship (See Hazel 1941[6], equation 16) the expected accuracy of this $\hat{G}_{EBV}$:

$$\mathrm{corr}\left(\hat{G}_{EBV}, G\right) = \sqrt{\frac{n_{rel}r_{pr}^2}{\left(\frac{1}{h^2} + \left(n_{rel} - 1\right)r_{rr}\right)}} \qquad \text{(Eq. S5)}$$

For PA-FGRS, which is a non-linear combination of binary traits observed in relatives, we need a new equation, and what we are looking for is an estimate of:

$$\mathrm{corr}\left(\hat{G}_{PA-FGRS}, G\right) = \frac{\mathrm{Cov}\left(\hat{G}_{PA-FGRS}, G\right)}{\sqrt{\mathrm{Var}\left(\hat{G}_{PA-FGRS}\right)\mathrm{Var}(G)}} = \frac{\mathrm{Cov}\left(\hat{G}_{PA-FGRS}, G\right)}{\sqrt{\mathrm{Var}\left(\hat{G}_{PA-FGRS}\right)h_l^2}} \qquad \text{(Eq. S6)}$$

Under the simple polygenic liability threshold model, we have a vector of of true genetic liabilities of the probands, $\mathbf{g}$, an $(m \times n)$ matrix $\mathbf{L}$ with the liabilities of the $n$ relatives of each of the $m$ probands and a corresponding $(m \times n)$ matrix $\mathbf{D}$ with the disease status of the relatives (i.e. $d_{ij} = 1, l_{ij} > t;\ 0, l_{ij} < t$ ). The covariance among the liabilities of the relatives is given by $\mathrm{Cov}(\mathbf{L}_{,i}, \mathbf{L}_{,j}) = \{r_{ij}h_l^2,\ i \neq j;\ 1,\ i = j$ , where $r_{ij}$ is the relatedness coefficient for $i$th and the $j$ the relative and $h^2 = \mathrm{Var}(G)$, and the covariance between the liabilities of the relatives and the genetic liability of the proband is given by $\mathrm{Cov}(\mathbf{L}_{,i}, \mathbf{g}) = r_{pi}h_l^2;\ i \neq p$, where $r_{pi}$ is relatedness coefficient for $i$th relative and the proband ($p$).

We could propose a simple linear predictor of $\mathbf{g}$ from the observed diseases in relatives. That could be $\hat{G}_{linear} = \mathbf{Db}$ , where $\mathbf{b} = \mathbf{P}^{-1}\mathbf{c}$, in which $\mathbf{P}_{ij} = \mathrm{Cov}(D_i, D_j)$ and

$\mathbf{c}_i = \mathrm{Cov}(G_p, D_i) = h_l^2 r_{pi} \phi(t)$ . When, as following the intuition in equation Eq. S5, all relatives are equally related to the index person, and equally related to each other, (i.e. $\mathbf{b}$ is constant) then we can define a random variable $\bar{D} = \frac{1}{n}\mathbf{D}\,\mathbf{1}'$ , where $\mathbf{1}$ is the unit 1-vector of size $n$, such that $\bar{D}$ is a vector with corresponds to the mean status of $n$ relatives. We can take the correlation between the mean of disease statuses in the relatives ($\bar{D}$) and $G$ as one potential (simplified) estimator for the accuracy of PA-FGRS.

$$\mathrm{corr}\left(\hat{G}_{PA-FGRS}, G\right) \approx \mathrm{corr}\left(\hat{G}_{linear}, G\right) = \mathrm{corr}\left(\bar{D}, G\right) = \frac{\mathrm{Cov}\left(\bar{D}, G\right)}{\sqrt{\mathrm{Var}\left(\bar{D}\right) h_l^2}} \qquad \text{(Eq. S7)}$$

We note that this is an approximation since $\mathrm{corr}\left(\hat{G}_{PA-FGRS}, \bar{D}\right) < 1$ when $n_{rel} > 1$. For a detailed discussion of this difference see Supplementary Note S10 below.

Treating $G$ and $\bar{D}$ as random variables and $L$ and $D$ as vectors of random variables we can estimate the expected accuracy, $\mathrm{corr}\left(\bar{D}, G\right)$, using that in $i$th relative :the covariance between the random variables of disease status $D_i$ and liability $L_i$ is:

$$\mathrm{Cov}\left(L_i, D_i\right) = \mathrm{E}(L_i D_i) - \mathrm{E}(L_i)\mathrm{E}(D_i) = \mathrm{E}(D_i L_i) = K\mathrm{E}(L_i | L_i > t) = \varphi(t)$$

Then by the additive law of covariance we have

$$\mathrm{Cov}\left(\bar{D}, G\right) = \mathrm{Cov}\left(G, \frac{1}{n_{rel}} \sum_{i=1}^{n_{rel}} D_i\right) = \frac{1}{n_{rel}} \sum_{i=1}^{n_{rel}} \mathrm{Cov}\left(G, D_i\right) =$$

$$\frac{1}{n_{rel}} \sum_{i=1}^{n_{rel}} \mathrm{Cov}\left(G, L_i\right)\mathrm{Cov}\left(L_i, D_i\right) = h_l^2 r_{pr} \varphi(t)$$

Then

$$\begin{aligned}\mathrm{Var}\left(\bar{D}\right) &= \mathrm{Var}\left(\frac{1}{n_{rel}} \sum_{i=1}^{n_{rel}} \left(D_i\right)\right) \\ &= \frac{1}{n_{rel}^2}\left(n_{rel} K(1 - K) + n_{rel}\left(n_{rel} - 1\right)\rho\, K(1 - K)\right) \\ &= \frac{1+\left(n_{rel}-1\right)\rho}{n_{rel}} K(1 - K)\end{aligned}$$

where ρ denotes the Pearson correlation of the disease status of the relatives.

Inserting these into *Eq.S7* we have:

$$\mathrm{corr}\left(\bar{D}_{r'}, G\right) = \sqrt{\frac{n_{rel} r_{pr}^2 \varphi(t)^2}{\left(1+\left(n_{rel}-1\right)\rho\right) K(1-K)}}$$

ρ can be estimated by a first order approximation (see Golan et al[3] as the expected intraclass correlation $\rho_{1st}$ of disease status among relatives as:

$$\rho \approx \rho_{1st} = \frac{\varphi(t)^2 r_{rr} h_l^2}{K(1-K)}$$

Plugging this in provides a 'first order' estimator of the expected accuracy for a PA-FGRS computed from equally related relatives as,

$$\mathrm{corr}\left(\bar{D}_{r'}, G\right) \approx \sqrt{\frac{n_{rel} h_l^2 r_{pr}^2 \varphi(t)^2}{\left(1+\left(n_{rel}-1\right)\frac{\varphi(t)^2 r_{rr} h_l^2}{K(1-K)}\right) K(1-K)}} = \sqrt{\frac{n_{rel} r_{pr}^2}{\frac{K(1-K)}{\varphi(t)^2 h_l^2} + \left(n_{rel}-1\right) r_{rr}}} \qquad \text{(Eq. S8)}$$

A better approximation can be obtained plugging in a second order Taylor approximation for ρ , as described by Golan et al[3]:

$$\rho \approx \rho_{2nd} = \frac{\varphi(t)^2 r_{rr} h_l^2}{K(1-K)} + \frac{t^2 \varphi(t)^2 h_l^4 r_{rr}^2}{2K(1-K)}$$

Rearranging this gives us:

$$\rho_{2nd} = \frac{\varphi(t)^2 r_{rr} h_l^2}{K(1-K)} \left(1 + \frac{r_{rr} h_l^2 t^2}{2}\right)$$

Providing a 'second order' estimator of the expected accuracy for a PA-FGRS computed from equally related relatives,

$$\mathrm{corr}\left(\bar{D}_r, G\right) \approx \sqrt{\frac{n_{rel} h_l^2 r_{pr}^2 \varphi(t)^2}{\left(1+\left(n_{rel}-1\right)\frac{\varphi(t)^2 r_{rr} h_l^2}{K(1-K)}\left(1+\frac{r_{rr} h_l^2 t^2}{2}\right)\right)K(1-K)}} = \sqrt{\frac{n_{rel} r_{pr}^2}{\frac{K(1-K)}{\varphi(t)^2 h_l^2}+\left(n_{rel}-1\right)r_{rr}\left(1+\frac{r_{rr} h_l^2 t^2}{2}\right)}} \qquad \text{(Eq. S9)}$$

Where, $n_{\mathrm{rel}}$ is the number of relatives, $r_{pr}$ is the relatedness between the index individual and the relatives, $r_{rr}$ is the fixed relatedness among relatives, $K$ the population prevalence of the disease, $h_l^2$ the heritability, $t$, the liability threshold, and $\varphi(t)$ the density of the standard normal distribution at the threshold. In simulations this second order estimator adequately approximates expected accuracy (Supplementary Figure S14). However, in the case of $r_{rr} = 0.5, h^2 > 0.75,\ K = 0.01$ , the approximation becomes inaccurate indicating that the equation will not hold for highly heritable, rare phenotypes with many related individuals in the pedigree which can also be seen from the accuracy of the approximation $\rho \approx \rho_{2nd}$ (Supplementary Figure S13).

An exact estimator for ρ can be obtained by taking the double integral of the bivariate normal distribution.

$$\rho = \rho_{exact} = \frac{\int_t^\infty \int_t^\infty \phi_2(x, y, r_{rr} h_l^2)\, dx\, dy - K^2}{K(1-K)}$$

where $\int_t^\infty \int_t^\infty \varphi_2(x, y,\ r_{rr} h^2)\, dx\, dy$ is the pdf for the bivariate standard normal distribution with correlation $r_{rr} h^2$.

Inserting this gives us,

$$\mathrm{corr}\left(\hat{G}_{linear}, G\right) = \sqrt{\frac{n_{rel} h_l^2 r_{pr}^2 \varphi(t)^2}{K(1-K)+\left(n_{rel}-1\right)\left(\int_t^\infty \int_t^\infty \phi_2(x, y, r_{rr} h_l^2)\, dx\, dy - K^2\right)}} \qquad \text{(Eq. S10)}$$

Eq. S10 suggests several intuitions about the expected accuracy of PA-FGRS.

First, if based on single relative, e.g., a family history indicator, the expected accuracy reduces to,

$$\mathrm{corr}\left(\hat{G}_{Linear-1rel}, G\right) = \frac{r_{pr}\varphi(t)\sqrt{h_l^2}}{\sqrt{K(1-K)}}$$

Second, if the relatives are not related to each other, the reliability (squared accuracy) increases linearly with the number of relatives (i.e. two parents explain twice the variance of one parent). Third, the same accuracy is achieved by *one* parent ($0.5\sqrt{\frac{h_l^2\varphi(t)^2}{K(1-K)}}$) and *four* unrelated ($\rho = 0$) grandparents ($0.25\sqrt{4}\sqrt{\frac{h_l^2\varphi(t)^2}{K(1-K)}}$), illustrating that the higher relatedness (of a parent) outweighs the higher number of individuals (grandparents). Fourth, the accuracy will increase with increasing $h_l^2$ and prevalence up to 0.5 where $\frac{\varphi(t)}{K(1-K)}\left(= \frac{\varphi(\Phi^{-1}(K))}{K(1-K)}\right)$ is maximized.

#### S.4 The asymptotic accuracy of $\hat{G}_{linear}$ from one class of relatives

For certain types of relatives, i.e., those where there is no constraint on the number an index individual can have (where $r_{rr} > 0$ ), such as when estimating liabilities only from siblings ( $r_{pr} = r_{rr} = 0.5$), only from cousins ($r_{pr} = r_{rr} = 0.125$), or only from off-spring ($r_{pr} = r_{rr} = 0.5$ or $r_{pr} = 0.5$ , $r_{rr} = 0.25$ ) , we can get a theoretical upper limit of accuracy by:

$$\begin{aligned}
\lim_{n_{rel}\to\infty} \mathrm{Var}\left(\bar{D}\right) &= \lim_{n_{rel}\to\infty} \frac{1+\left(n_{rel}-1\right)\rho}{n_{rel}} K(1-K) \\
&= \lim_{n_{rel}\to\infty} \left(\frac{1}{n_{rel}} + \frac{\left(n_{rel}-1\right)\rho}{n_{rel}}\right) K(1-K) \\
&= \lim_{n_{rel}\to\infty} \frac{\left(n_{rel}-1\right)\rho}{n_{rel}} K(1-K) \\
&= \rho K(1-K)
\end{aligned}$$

$$\begin{aligned}
\lim_{n_{rel}\to\infty} \mathrm{corr}\left(\bar{D}, G\right) &\approx \lim_{n_{rel}\to\infty} \frac{h_l^2 r_{pr}\varphi(t)}{\sqrt{\frac{1+\left(n_{rel}-1\right)\rho}{n_{rel}} K(1-K)h_l^2}} \\
&= \frac{h_l^2 r_{pr}\varphi(t)}{\sqrt{\rho\, K(1-K)h_l^2}}
\end{aligned}$$

$$= \sqrt{\frac{h_l^2 r_{pr}^2 \varphi(t)^2}{\rho \; K(1-K)}}$$

$$= \sqrt{\frac{h_l^2 r_{pr}^2 \varphi(t)^2}{\int_t^\infty \int_t^\infty \varphi_2(x, y, r_{rr} h_l^2)\, dx\, dy - K^2}} \qquad \text{(Eq. S11)}$$

replacing the integral with the second order Taylor approximation ($\rho \approx \rho_{2nd}$) gives us a closed form approximation:

$$\approx \frac{h_l^2 r_{pr} \varphi(t)}{\sqrt{\frac{\phi(t)^2 r_{rr} h_l^2}{K(1-K)} \left(1 + \frac{r_{rr} h_l^2 t^2}{2}\right) h_l^2 K(1-K)}}$$

$$= \frac{r_{pr}}{\sqrt{r_{rr}}} \frac{1}{\sqrt{1 + \frac{r_{rr} h_l^2 t^2}{2}}} \qquad \text{(Eq. S12)}$$

Supplementary Figure S16 show that Eq.S8 provides reasonable estimates of the asymptotic accuracy of a linear predictor $\mathrm{corr}\left(\bar{D}, G\right)$. Demonstrating that the further the prevalence is from 0.5 and the higher the heritability, the more will the asymptotic accuracy at an infinite number of relatives be lower than $\frac{r_{pr}}{\sqrt{r_{rr}}}$, which is the maximum accuracy that can be achieved by a non-linear predictor (see section S.10).

#### S.5 Accuracy in an arbitrarily structured pedigree

As in *Eq.S6* above, we are looking for is an estimate of

$$\mathrm{corr}\left(\hat{G}_{FGRS}, G\right) = \frac{\mathrm{Cov}\left(\hat{G}_{FGRS}, G\right)}{\sqrt{\mathrm{Var}\left(\hat{G}_{FGRS}\right)\mathrm{Var}(G)}} = \frac{\mathrm{Cov}\left(\hat{G}_{FGRS}, G\right)}{\sqrt{\mathrm{Var}\left(\hat{G}_{FGRS}\right) h_l^2}}$$

But one that does not rely on the simplifying assumption of one relative class. If do, however, again consider the linear predictor of $\mathbf{g}$ defined as $\hat{G}_{linear} = \mathbf{Db}$, where $\mathbf{b} = \mathbf{P}^{-1}\mathbf{c}$, in which

$$\mathbf{P}_{ij, i \neq j} = \mathrm{Cov}(d_i, d_j) = \rho_{ij} K(1 - K)$$

$$\mathbf{P}_{ij,\, i=j} = Var(D_i) = K_{Pop}\left(1 - K_{Pop}\right)$$

$$\mathbf{c}_j = \mathrm{Cov}(G_p, D_j) = E(D_j) E(G_p | D_j = 1) = h_l^2 r_{pj} \varphi(t) .$$

and where ρ can be estimated as described in section S.3 ($\rho = \rho_{exact}$). Then we can write,

$$\mathrm{Cov}\left(\hat{G}_{linear}, G\right) = \mathbf{c}'\,\mathbf{b} = \mathbf{c}'\ \mathbf{P}^{-1}\mathbf{c}$$

We can also get the variance of $\hat{G}_{linear}$ as,

$$\mathrm{Var}\left(\hat{G}_{linear}\right) = \mathrm{Var}\left(\sum_{j=1}^{n_{rel}}\left(b_j D_j\right)\right) = \sum_{j=1}^{n_{rel}}\left(b_j^2 p_{jj}\right) + \sum_{j=1}^{n_{rel}}\sum_{k=1}^{n_{rel}}\left(b_j b_k p_{ij};\ j\neq k\right)$$

Which can be equivalently written in matrix notation,

$$\mathrm{Var}\left(\hat{G}_{linear}\right) = \mathbf{b}'\,\mathbf{P}\,\mathbf{b} = (\mathbf{P}^{-1}\mathbf{c})'\ \mathbf{P}\,(\mathbf{P}^{-1}\mathbf{c}) = \mathbf{c}'\ \mathbf{P}^{-1}\mathbf{c}$$

Plugging this back into equation *Eq.S7*, gives

$$\mathrm{corr}\left(\hat{G}_{linear}, G\right) = \frac{\mathbf{c}'\ \mathbf{P}^{-1}\mathbf{c}}{\sqrt{\mathbf{c}'\ \mathbf{P}^{-1}\mathbf{c}}\sqrt{h_l^2}} = \sqrt{\frac{\mathbf{c}'\ \mathbf{P}^{-1}\mathbf{c}}{h_l^2}} \qquad \text{(Eq. S9)}$$

where $\mathbf{c}$ is a column vector of covariances between the phenotype of the relatives and the genetic value of the index individual, and $\mathbf{P}$ is the phenotypic covariance matrix of the relatives. Since *Eq. S13* involves the inverse of $\mathbf{P}$, it is harder to get an intuitive sense of this expression than Eq. S9, but this can be used to estimate the expected accuracy in more complex pedigrees.

#### S.6 Accuracy of linear predictor under censoring

If the phenotype status for the relatives is not the true disease status $D$, but the observed disease status $Y$ (as defined in Online Methods). This changes $\mathbf{c}$ to $\tilde{\mathbf{c}}$ where

$$\tilde{\mathbf{c}}_i = \mathrm{Cov}(G, Y_i) = E(Y_i)E(D_i|Y_i = 1)E(G|D_i = 1) + (1 - E(Y_i))E(D_i|Y_i = 0)E(G|D_i = 0) =$$

$$= E(Y_i)E(G|D_i = 1) = \frac{K_i}{K_{pop}} h_l^2 r_{pi} \varphi(t)$$

then we can define,

$$\mathrm{Cov}\left(\hat{G}_{linear-censored}, G\right) = \tilde{\mathbf{c}}' \tilde{\mathbf{P}}^{-1} \tilde{\mathbf{c}}$$

Where $\tilde{\mathbf{P}}$ is ($n_{rel}$ by $n_{rel}$) covariance matrix of the observed phenotypes in the relatives such that,

$$\tilde{p}_{ij} = \mathrm{Cov}(Y_i, Y_j) = \begin{cases} \mathrm{Cov}(D_i, D_j) \dfrac{K_i K_j}{K_{pop}^2} & i \neq j \\ K_i\left(1 - K_i\right) & i = j \end{cases}$$

#### S.7 The expected accuracy of PGS

The accuracy of a PGS, has previously been shown to be a function of the $h_{SNP}^2$ and the statistical power of the discovery study [7]. Given a discovery study $N_{case}$ cases and $N_{ctrl}$ controls and $M$ independent predictors (genotypes):

$$\mathrm{corr}\left(\hat{G}_{PGS}, G\right)^2 = \frac{\lambda w h_l^2 (i_q - \bar{i})^2}{\lambda w h_l^2 (i_q - \bar{i})^2 + (1-w)\frac{var^*(x_j)}{var(x_j)}} \qquad \text{(Eq. S14)}$$

in which $\lambda = \frac{N_{case} + N_{ctrl}}{M}$, $w = \frac{N_{case}}{N_{case} + N_{ctrl}}$ , $i_q = \frac{\varphi(t)}{K}$, $\bar{i} = w\frac{\varphi(t)}{K} - (1 - w)\frac{-\varphi(t)}{1-K}$. $\frac{\mathrm{Var}^*(x_j)}{\mathrm{Var}(x_j)}$ is the ratio of the expected variance of a genotype in the ascertained sample to that in a population sample.

$$\mathrm{Var}^*(x_j) = \mathrm{Var}(x_j) - \frac{h_l^2}{M}\bar{i}(\bar{i} - t)\mathrm{Var}(x_j)$$

Dudbrigde et al.[8] ignored this difference, as for polygenic traits $\frac{h_l^2}{M}$ can be assumed to be very close to 0. Note that the derivations in Daewtyler [7] suggest $var*(x_j) = var(x_j) - h_l^2 i(\bar{i} - t)var(x_j)$, omitting the factor M and thereby implying the contribution of ascertainment would have a non-negligible effect on the genetic variance, but we believe this to be an error.

This expectation can be written equivalently, as shown in Wu et al [9], by defining,

$$N_q = (N_{case} + N_{ctrl})w(1-w)\left(\frac{\varphi(t)}{K*(1-K)}\right)^2$$

which gives,

$$\mathrm{corr}\left(\hat{G}_{PGS}, G\right)^2 = \frac{h_l^2 N_q}{h_l^2 N_q + M} \qquad \text{(Eq.15)}$$

#### S.8 The expected accuracy of shrinkage PGS

*Eq. S14* and *Eq. S15* work under the assumption that PGS includes all markers (i.e. no p-value thresholding). Recently PGS methods that apply a bayesian shrinkage to the effect size estimates have been shown to be more efficient. If the true effect sizes have a pointnormal distribution:

$$\beta \sim (1-p)\,N(0, 0) + p\,N(0, \frac{h_{l,SNP}^2}{Mp})$$

Typically $\hat{\beta}$, will be estimated on the observed scale such that[8] $\hat{\beta}_i \approx \left(\beta_i + \sum_j^M r_{ij}\beta_j\right)\frac{w(1-w)}{K(1-K)\varphi(t)} + \epsilon$, where $\mathrm{Var}(\epsilon) \approx \frac{w(1-w)}{N}$. This gives gives us the well know relationship between the SNP-heritability and observed scale[10] ($h_{o,SNP}^2$) and the liability scale ($h_{l,SNP}^2$):

$$h_{o,SNP}^2 = M\,\mathrm{Var}\left(\beta\frac{w(1-w)}{K(1-K)\varphi(t)}\right)/w(1-w) = \frac{w(1-w)}{(K(1-K)\varphi(t))^2}M\,\mathrm{Var}(\beta) = \frac{w(1-w)}{(K(1-K)\varphi(t))^2}h_{l,SNP}^2$$

Since the SNP-heritability $h_{l,SNP}^2$ is on the liability scale, we transform $\hat{\beta}$ to $\hat{\beta}_l = \hat{\beta}\frac{K(1-K)\varphi(t)}{w(1-w)} = \left(\beta_i + \sum_j^M r_{ij}\beta_j\right) + \epsilon\frac{K(1-K)\varphi(t)}{w(1-w)}$. This transformation changes the residual variance to:

$$\mathrm{Var}(\epsilon\frac{K(1-K)\varphi(t)}{w(1-w)}) \approx \frac{w(1-w)}{N}\left(\frac{K(1-K)\varphi(t)}{w(1-w)}\right)^2 = \frac{1}{N}\frac{(K(1-K)\varphi(t))^2}{w(1-w)} = \frac{1}{N_q}$$

Under this model $\hat{\beta}$ can be shrunk[11] to $\hat{\beta}_{j_{shrinkage}} = E(\beta_j|\hat{\beta}_{j,l}) = \hat{\beta}_{j,l}\bar{p}_j\left(\frac{1}{1+\frac{Mp}{h^2_{l,SNP}N}}\right)$, where:

$$\bar{p}_j = P(\beta_j \neq 0|\hat{\beta}_{j,l}, N_q, M, p, h^2_l) = \frac{\frac{p}{\sqrt{\frac{h^2_{l,SNP}}{Mp}+\frac{1}{N_q}}}exp\left\{-\frac{1}{2}\frac{\hat{\beta}^2_{j,l}}{\frac{h^2_{l,SNP}}{Mp}+\frac{1}{N_q}}\right\}}{\frac{p}{\sqrt{\frac{h^2_{l,SNP}}{Mp}+\frac{1}{N}}}exp\left\{-\frac{1}{2}\frac{\hat{\beta}^2_{j,l}}{\frac{h^2_{l,SNP}}{Mp}+\frac{1}{N_q}}\right\}+\frac{1-p}{\sqrt{\frac{1}{N_q}}}exp\left\{-\frac{1}{2}N_q\hat{\beta}^2_{j,l}\right\}}$$

When deriving the expected squared accuracy of $\hat{\beta}_{j_{shrinkage}}$ we have:

$$\mathrm{Var}(\hat{G}_{PGS-shrinkage}) = \mathrm{Cov}(\hat{G}_{PGS-shrinkage}, G)$$

and

$$\mathrm{corr}\left(\hat{G}_{PGS-shrinkage}, G\right)^2 = \frac{\left(\mathrm{Var}(\hat{G}_{PGS-shrinkage})\right)^2}{h^2_{l,SNP}\mathrm{Var}(\hat{G}_{PGS-shrinkage})} = \frac{\mathrm{Var}(\hat{G}_{PGS-shrinkage})}{h^2_{l,SNP}}$$

however, $\mathrm{Var}(\hat{G}_{PGS-shrinkage}) \leq h^2_{l,SNP} + \frac{M}{N_q}$ and can be estimated by:

$$\mathrm{Var}(\hat{G}_{PGS-shrinkage}) = \mathrm{Var}(\sum_{j=1}^{M} x_j\hat{\beta}_{j_{shrinkage}}) = \mathrm{Var}(\hat{\beta}_{shrinkage})\sum_{j=1}^{M}\mathrm{Var}(x_j) = (\hat{\beta}_{shrinkage})M$$

Thus

$$\mathrm{corr}\left(\hat{G}_{PGS-shrinkage}, G\right)^2 = \frac{M}{h^2_{l,SNP}}\int_{-\infty}^{\infty}\left(x\frac{1}{1+\frac{Mp}{h^2_{l,SNP}N_q}}P(\beta_j \neq 0|x, N_q, M, p, h^2_{l,SNP})\right)^2 P(x|N_q, M, p, h^2_{l,SNP})dx$$

$$= \frac{M}{h^2_{l,SNP}}\left(\frac{1}{1+\frac{Mp}{h^2_{l,SNP}N_q}}\right)^2\int_{-\infty}^{\infty} x^2P(\beta_j \neq 0|x, N_q, M, p, h^2_{l,SNP})^2 P(x|N_q, M, p, h^2_{l,SNP})dx$$

$$= \left(\frac{h^2_{l,SNP}}{h^2_{l,SNP} + \frac{Mp}{N_q}}\right)\left(\frac{1}{\frac{h^2_{l,SNP}}{Mp} + \frac{1}{N_q}}\right)\int_{-\infty}^{\infty} x^2 P(\beta_j \neq 0 | x, N_q, M, p, h^2_{l,SNP}) P(x | \beta_j \neq 0, N_q, M, p, h^2_{l,SNP}) dx$$

$$= \left(\frac{h^2_{l,SNP}}{h^2_{l,SNP} + \frac{Mp}{N_q}}\right)\left(\frac{1}{\frac{h^2_{l,SNP}}{Mp} + \frac{1}{N_q}}\right)\int_{-\infty}^{\infty} x^2 \frac{\frac{p}{\sqrt{\frac{h^2_{l,SNP}}{Mp} + \frac{1}{N_q}}} exp\left\{-\frac{1}{2}\frac{x^2}{\frac{h^2_{l,SNP}}{Mp} + \frac{1}{N_q}}\right\}}{\frac{p}{\sqrt{\frac{h^2_{l,SNP}}{Mp} + \frac{1}{N_q}}} exp\left\{-\frac{1}{2}\frac{x^2}{\frac{h^2_{l,SNP}}{Mp} + \frac{1}{N_q}}\right\} + \frac{1-p}{\sqrt{\frac{1}{N_q}}} exp\left\{-\frac{1}{2}N_q x^2\right\}} \frac{1}{\sqrt{\frac{h^2_{l,SNP}}{Mp} + \frac{1}{N_q}}} exp\left\{-\frac{1}{2}\frac{x^2}{\frac{h^2_{l,SNP}}{Mp} + \frac{1}{N_q}}\right\} dx.$$

(Eq.S16)

which can be evaluated with numerical integration techniques. Note that if $p = 1$, this reduces to $\mathrm{corr}\left(\hat{G}_{PGS-shrinkage}, G\right)^2 = \left(\frac{h^2_{l,SNP}}{h^2_{l,SNP} + \frac{M}{N_q}}\right)$ which is identical to Eq. S15. As in Eq. S15, we can substitute $N$ for $N_q$, and $h^2_{SNP}$ for $h^2_{l,SNP}$ if the outcome is a quantitative trait.

#### S.9 Expected correlation of PGS and PA-FGRS assuming conditional independence

Under the assumption the that $\hat{G}_{FGRS}$ and $\hat{G}_{PGS}$ are statistically independent conditional on the true genetic value ($G_i$), their correlation will be the product of their accuracies. We can show this using the law of total covariance:

$$
\begin{aligned}
\mathrm{corr}\left(\hat{G}_{PGS}, \hat{G}_{FGRS}\right) &= \frac{\mathrm{E}\left(\mathrm{Cov}\left(\hat{G}_{PGS}, \hat{G}_{FGRS}|G\right)\right) + \mathrm{Cov}\left(\mathrm{E}\left(\hat{G}_{PGS}|G\right)\mathrm{E}\left(\hat{G}_{FGRS}|G\right)\right)}{\sqrt{\mathrm{Var}(\hat{G}_{PGS})\mathrm{Var}(\hat{G}_{FGRS})}} \\
&= \frac{\mathrm{Cov}\left(\mathrm{E}\left(\hat{G}_{PGS}|G\right)\mathrm{E}\left(\hat{G}_{FGRS}|G\right)\right)}{\sqrt{\mathrm{Var}(\hat{G}_{PGS})\mathrm{Var}(\hat{G}_{FGRS})}} \\
&= \frac{\frac{\mathrm{Cov}\left(\hat{G}_{PGS}, G\right)}{\mathrm{Var}(G)} \frac{\mathrm{Cov}\left(\hat{G}_{FGRS}, G\right)}{\mathrm{Var}(G)} \mathrm{Var}(G)}{\sqrt{\mathrm{Var}(\hat{G}_{PGS})\mathrm{Var}(\hat{G}_{FGRS})}} \\
&= \frac{\mathrm{Cov}\left(\hat{G}_{PGS}, G\right)}{\sqrt{\mathrm{Var}(\hat{G}_{PGS})\mathrm{Var}(G)}} \frac{\mathrm{Cov}\left(\hat{G}_{FGRS}, G\right)}{\sqrt{\mathrm{Var}(\hat{G}_{FGRS})\mathrm{Var}(G)}} \\
&= \mathrm{corr}\left(\hat{G}_{PGS}, G\right)\mathrm{corr}\left(\hat{G}_{FGRS}, G\right)
\end{aligned}
$$

(Eq. S17)

Using the using estimates of $R_l^2$ and narrow-sense heritability, we can also calculate the expected correlation between estimated genetic values under the assumption of conditional independence as:

$$
\mathrm{corr}\left(\hat{G}_{PGS}, \hat{G}_{FGRS}\right) = \sqrt{\frac{R^2_{l,PGS} R^2_{l,FGRS}}{h^2_l h^2_l}} = \frac{\sqrt{R^2_{l,PGS} R^2_{l,FGRS}}}{h^2_l} \qquad \text{(Eq. S18)}
$$

#### S.10 On the asymptotic difference between linear predictors and PA-FGRS

If we again imagine the situation where we are studying a population in which all probands have $n$ relatives, that are all equally related to each other, with a relatedness coefficient $r_{rr}$ and equally related to the proband with relatedness coefficient $r_{pr}$.

$$\mathrm{corr}(G, \hat{G}_{linear}) = \mathrm{corr}(G, \bar{D}) = \sqrt{\frac{n_{rel} h_l^2 r_{pr}^2 \phi(t)^2}{K(1-K)+(n_{rel}-1)\left(\int_t^\infty \int_t^\infty \phi_2(x,y,r_{rr}h_l^2)\,dx\,dy - K^2\right)}}$$

As shown in **S.4** , as $n_{rel} \rightarrow \infty$, this accuracy with approach

$$\sqrt{\frac{h_l^2 r_{pr}^2 \phi(t)^2}{\int_t^\infty \int_t^\infty \phi_2(x,y,r_{rr}h^2)\,dx\,dy - K^2}} \approx \frac{r_{pr}}{\sqrt{r_{rr}}} \frac{1}{\left(1+\frac{r_{rr}h^2 t^2}{2}\right)}$$

However the PA-FGRS estimator of $G$, $\hat{G}_{PA}$ has a slightly different expected accuracy. First since $\hat{G}_{PA-FGRS}$ is an unbiased estimator of $G$, we have $E(G|\hat{G}_{PA}) = \hat{G}_{PA}$, and by the law of total covariance we can write,

$$\frac{\mathrm{Cov}(G,\hat{G}_{PA})}{\mathrm{Var}(\hat{G}_{PA})} = \frac{\mathrm{Cov}(G,\hat{G}_{PA}|\hat{G}_{PA}) + \mathrm{Cov}(E(G|\hat{G}_{PA}), E(\hat{G}_{PA}|\hat{G}_{PA}))}{\mathrm{Var}(\hat{G}_{PA})} = \frac{E(\hat{G}_{PA}^{\,2})}{\mathrm{Var}(\hat{G}_{PA})} = 1$$

we then have:

$$\mathrm{corr}(G, \hat{G}_{PA}) = \frac{\mathrm{Cov}(G,\hat{G}_{PA})}{\sqrt{\mathrm{Var}(\hat{G}_{PA})\mathrm{Var}(G)}} = \frac{\mathrm{Var}(\hat{G}_{PA})}{\sqrt{\mathrm{Var}(\hat{G}_{PA})\mathrm{Var}(G)}} = \sqrt{\frac{\mathrm{Var}(\hat{G}_{PA})}{h_l^2}} \qquad \text{(Eq. S19)}$$

and by the law of total variance we have:

$$\mathrm{Var}(\hat{G}_{PA}) = \mathrm{Var}(E(G|D_{1,\ldots,n_{rel}})) = \mathrm{Var}(G) - \mathrm{Var}(G|D_{1,\ldots,n_{rel}})$$

$\mathrm{Var}(G|D_{1,\ldots,n_{rel}})$ is estimated by the PA formula as:

$$\Omega^* = \Omega - \Omega_{,x}\left(\Omega_{x,x}^{-1} - \Omega_{x,x}^{-1}\Omega_{x,x}^*\Omega_{x,x}^{-1}\right)\Omega_{x,}$$

where Ω is the covariance matrix after conditioning on ($k$ *-1*) relatives, and $\Omega^*$ is the covariance after conditioning on $k$ relatives.

If we define: $\delta_k = 1 - \Omega_{x,x}^{-1}\Omega_{x,x}^* = 1 - \frac{\mathrm{Var}(L_k|D_{1,}\cdots D_k)}{\mathrm{Var}(L_k|D_{1,}\cdots D_{k-1})}$, we have:

$$\delta_k = \begin{cases} \frac{\lambda_k \phi(\lambda_k)}{\Phi(\lambda_k)} + \left(\frac{\phi(\lambda_k)}{\Phi(\lambda_k)}\right)^2 & \text{if } D_k = 1 \\ \left(\frac{\phi(\lambda_k)}{1-\Phi(\lambda_k)}\right)^2 - \frac{\lambda_k \phi(\lambda_k)}{1-\Phi(\lambda_k)} & \text{if } D_k = 0 \end{cases}$$

where $\lambda_k = \frac{t - \mathrm{E}(L_k | D_{1,} \cdots D_{k-1})}{\sqrt{\mathrm{Var}(L_k | D_{1,} \cdots D_{k-1})}}$.

We can write the expected conditional variance of $G_p$ as:

$$\mathrm{Var}(G_p | D_1, \cdots, D_n) = \mathrm{Var}(G_p) - \frac{\mathrm{Cov}(G_p, L_1)^2}{\mathrm{Var}(L_1)}\delta_1 - \frac{\mathrm{Cov}(G_p, L_2 | D_1)^2}{\mathrm{Var}(L_2 | D_1)}\delta_2 - \cdots - \frac{\mathrm{Cov}(G_p, L_n | D_1, \cdots, D_{n-1})^2}{\mathrm{Var}(L_n | D_1, \cdots, D_{n-1})}\delta_n$$

Similarly, we have:

$$\mathrm{Cov}(G_p, L_n | D_1, \cdots, D_{n-1}) = \mathrm{Cov}(G_p, L_n) - \frac{\mathrm{Cov}(G_p, L_1)\mathrm{Cov}(L_n, L_1)}{\mathrm{Var}(L_1)}\delta_1 - \frac{\mathrm{Cov}(G_p, L_2 | D_1)\mathrm{Cov}(L_n, L_2 | D_1)}{\mathrm{Var}(L_2 | D_1)}\delta_2 - \cdots - \frac{\mathrm{Cov}(G_p, L_{n-1} | D_1, \cdots, D_{n-2})\mathrm{Cov}(L_n, L_{n-1} | D_1, \cdots, D_{n-2})}{\mathrm{Var}(L_{n-1} | D_1, \cdots, D_{n-2})}\delta_{n-1}$$

since $0 < \delta_k < 1$, $\mathrm{Var}(G_p | D_1, \cdots, D_n)$ and $\mathrm{Cov}(G_p, L_n | D_1, \cdots, D_{n-1})$ will both be monotonically decreasing as $n \to \infty$ with $\mathrm{Cov}(G_p, L_n | D_1, \cdots, D_{n-1}) \to 0$. Meaning that in the limit conditioning on $D_{n+1}$ will not affect $\mathrm{Var}(G_i | D_{1,\ldots,n})$.

Since all $\mathrm{Cov}(G_p, L_k) = h_l^2 r_{pr}$ , we can factor out $-h_l^2 r_{pr}$ from $\mathrm{Var}(G_p | D_1, \cdots, D_n)$ which becomes:

$$\mathrm{Var}(G_p | D_1, D_2, \cdots, D_n) = \mathrm{Var}(G_p) - \frac{\mathrm{Cov}(G_p, L_1)^2}{\mathrm{Var}(L_1)}\delta_1 - \frac{\left(\mathrm{Cov}(L_2, G_p) - \frac{\mathrm{Cov}(L_2, L_1)\mathrm{Cov}(G_p, L_1)}{\mathrm{Var}(L_1)}\delta_1\right)\mathrm{Cov}(G_p, L_2 | D_1)}{\mathrm{Var}(L_2 | D_1)}\delta_2 - (\cdots)$$

$$= \mathrm{Var}(G_p) - h_l^2 r_{pr} \left(\frac{\mathrm{Cov}(G_p, L_1)}{\mathrm{Var}(L_1)}\delta_1 + \frac{\left(1 - \frac{\mathrm{Cov}(L_2, L_1)}{\mathrm{Var}(L_1)}\delta_1\right)\mathrm{Cov}(G_p, L_2 | D_1)}{\mathrm{Var}(L_2 | D_1)}\delta_2 + (\cdots)\right)$$

(Eq. S20)

And similarly, since all $\mathrm{Cov}(L_j, L_k) = h_l^2 r_{rr}$ ; $j \neq k$ we can factor out $- h_l^2 r_{rr}$ from $\mathrm{Cov}(G_p, L_n | D_1, D_2, \cdots, D_n)$ which becomes:

$$\mathrm{Cov}(G_p, L_n | D_1, D_2, \cdots, D_n) =$$

$$\mathrm{Cov}(G_p, L_n) - \frac{\mathrm{Cov}(G_p, L_1)\mathrm{Cov}(L_n, L_1)}{\mathrm{Var}(L_1)}\delta_1 - \frac{\left(\mathrm{Cov}(L_2, L_n) - \frac{\mathrm{Cov}(L_2, L_1)\mathrm{Cov}(L_n, L_1)}{\mathrm{Var}(L_1)}\delta_1\right)\mathrm{Cov}(G_p, L_2 | D_1)}{\mathrm{Var}(L_2 | D_1)}\delta_2 - (\cdots) =$$

$$\mathrm{Cov}(G_p, L_n) - h_l^2 r_{rr} \left( \frac{\mathrm{Cov}(G_p, L_1)}{\mathrm{Var}(L_1)}\delta_1 + \frac{\left(1 - \frac{\mathrm{Cov}(L_2, L_1)}{\mathrm{Var}(L_1)}\delta_1\right)\mathrm{Cov}(G_p, L_2 | D_1)}{\mathrm{Var}(L_2 | D_1)}\delta_2 + (\cdots) \right)$$

As $n_{rel} \rightarrow \infty$, the $\mathrm{Cov}(G_p, L_n | D_1, D_2, \cdots, D_n)$ will approach 0:

$$\lim_{n \to \infty} \mathrm{Cov}(G_p, L_n | D_1, D_2, \cdots, D_n) = 0 =$$

$$\lim_{n \to \infty} \mathrm{Cov}(G_p, L_n) - \mathrm{Cov}(L_j, L_k) \left( \frac{\mathrm{Cov}(G_p, L_1)}{\mathrm{Var}(L_1)}\delta_1 + \frac{\left(1 - \frac{\mathrm{Cov}(L_2, L_1)}{\mathrm{Var}(L_1)}\delta_1\right)\mathrm{Cov}(G_p, L_2 | D_1)}{\mathrm{Var}(L_2 | D_1)}\delta_2 + (\cdots) \right)$$

which we can rearrange to get:

$$\lim_{n \to \infty} \left( \frac{\mathrm{Cov}(G_p, L_1)}{\mathrm{Var}(L_1)}\delta_1 + \frac{\left(1 - \frac{\mathrm{Cov}(L_2, L_1)}{\mathrm{Var}(L_1)}\delta_1\right)\mathrm{Cov}(G_p, L_2 | D_1)}{\mathrm{Var}(L_2 | D_1)}\delta_2 + (\cdots) \right) = \frac{\mathrm{Cov}(G_p, L_n)}{\mathrm{Cov}(L_j, L_k)} \quad \text{(Eq.21)}$$

inserting this into (Eq. S20), gives us:

$$\lim_{n \to \infty} \mathrm{Var}(G_p | D_1, \cdots, D_n) = \mathrm{Var}(G_p) - \mathrm{Cov}(G_p, L_k)\frac{\mathrm{Cov}(G_p, L_k)}{\mathrm{Cov}(L_j, L_k)}$$

and thus

$$\lim_{n \to \infty} \mathrm{Var}(E(G_p | D_1, \cdots, D_n)) = \frac{(h_l^2 r_{pr})^2}{h_l^2 r_{rr}} = \frac{h_l^2 r_{pr}^2}{r_{rr}}$$

giving the asymptotic accuracy of:

$$\lim_{n\to\infty} \mathrm{corr}(G_p, \hat{G}_{PA}) = \sqrt{\frac{h_l^2 r_{pr}^2}{r_{rr} h^2}} = \frac{r_{pr}}{\sqrt{r_{rr}}} .$$

#### S.11 Exact estimator for PA-FGRS accuracy

While this gives the asymptote, we can also derive the expected accuracy for of the PA-FGRS, starting with Eq.S6, we have:

$$\mathrm{corr}(G, \hat{G}_{PA}) = \sqrt{\frac{\mathrm{Var}(\hat{G}_{PA})}{h_l^2}}$$

We can obtain the the expected variance of $\hat{G}_{PA}$ given number of relatives $n_{rel}$, prevalence $K$ and the variance-covariance matrix, $\Sigma$, for the random vector of lilities $\left[G_p, L_1, \cdots, L_{n_{rel}}\right]$, If $d$ denotes the set of $m = 2^{n_{rel}}$ possible configurations of the vector $D$, $\hat{G}_{PA}$will be discrete mixture distribution:

$$\hat{G}_{PA} \sim \sum_{i=1}^{m} P\left(D = d_i, \Sigma, K\right) \mathrm{E}\left(G | D = d_i,, \Sigma, K\right)$$

and

$$\mathrm{Var}(\hat{G}_{PA}) = \sum_{i=1}^{m}\left(P\left(D = d_i, \Sigma, K\right) \mathrm{E}\left(G | D = d_i, \Sigma, K\right)^2\right)$$

However, if $n_{rel}$ is large, numerical integration of the $m = 2^{n_{rel}}$ , $n_{rel}$-variate normal distributions will quickly become infeasible. E.g. with $n_{rel} = 20$ relatives $m = 2^{20} = 1\,048\,576$. However, if all $\mathrm{Cov}(G, L_k) = h^2 r_{pr}$ and all $\mathrm{Cov}(L_j, L_k) = h^2 r_{rr}$ ; $j \neq k$. The probability mass function can be written as a simpler mixture distribution:

$$\hat{G}_{PA} \sim \sum_{n_{aff}=0}^{n_{rel}} \frac{n_{rel}}{n_{aff}} P\left(\sum_{j=1}^{n_{rel}} D_j = n_{aff}, n_{rel}, \Sigma, K\right) \mathrm{E}\left(G | \sum_{j=1}^{n_{rel}} D_j = n_{aff}, n_{rel}, \Sigma, K\right)$$

which has a variance of:

$$\mathrm{Var}(\hat{G}_{PA}) = \sum_{n_{aff}=0}^{n_{rel}} \left( \frac{n_{rel}}{n_{aff}} P\left(\sum_{j=1}^{n_{rel}} D_j = n_{aff},\ n_{rel}, \Sigma, K\right) \mathrm{E}\left(G | \sum_{j=1}^{n_{rel}} D_j = n_{aff},\ n_{rel}, \Sigma, K\right)^2 \right)$$

where

$$P\left(\sum_{j=1}^{n_{rel}} D_j = n_{aff},\ n_{rel}, \Sigma, K\right) = \int_{l_1}^{u_1}\int_{l_2}^{u_2}\cdots\int_{l_{n_{rel}}}^{u_{n_{rel}}} \phi_{n_{rel}}([0, \cdots, 0],\ \Sigma')$$

in which Σ' is the variance-covariance matrix for the random vector of liabilities in the relatives $\left[L_1, \cdots, L_{n_{rel}}\right]$, $l_i$ and $u_i$ are $-\ \infty$ and $t$ for the number of unaffected and $t$ and ∞ for the number of affected relatives. Inserting this into equation Eq.S19 we get:

$$\mathrm{corr}(G, \hat{G}_{PA}) = \sqrt{\frac{\sum_{n_{aff}=0}^{n_{rel}} \left( \frac{n_{rel}}{n_{aff}} P\left(\sum_{j=1}^{n_{rel}} D_j = n_{aff}, n_{rel}, \Sigma, K\right) \mathrm{E}\left(G | \sum_{j=1}^{n_{rel}} D_j = n_{aff}, n_{rel}, \Sigma, K\right)^2 \right)}{h^2}} \qquad \text{(Eq. S22)}$$

Using numerical estimates of $\int_{l_1}^{u_1}\int_{l_2}^{u_2}\cdots\int_{l_{n_{rel}}}^{u_{n_{rel}}} \phi_{n_{rel}}([0, \cdots, 0],\ \Sigma')$ and estimating $\mathrm{E}\left(G | \sum_{j=1}^{n_{rel}} D_j = n_{aff},\ n_{rel}, \Sigma, K\right)$ by the PA-FGRS estimator, we can compute the expected accuracy.

Supplementary Figure S15 shows the empirical accuracies of a linear estimator $\hat{G}_{linear}$, the $\hat{G}_{PA}$ and the expected accuracies as estimated by (Eq. S18).

### Supplementary References:


1. Lee, S. H., Goddard, M. E., Wray, N. R. & Visscher, P. M. A better coefficient of determination for genetic profile analysis. *Genet. Epidemiol.* **36**, 214–224 (2012).
2. Hujoel, M. L. A., Loh, P.-R., Neale, B. M. & Price, A. L. Incorporating family history of disease improves polygenic risk scores in diverse populations. *Cell Genom* **2**, (2022).
3. Golan, D., Lander, E. S. & Rosset, S. Measuring missing heritability: inferring the contribution of common variants. *Proc. Natl. Acad. Sci. U. S. A.* **111**, E5272–81 (2014).
4. Dempster, E. R. & Lerner, I. M. Heritability of Threshold Characters. *Genetics* **35**, 212–236 (1950).
5. Wray, N. R., Yang, J., Goddard, M. E. & Visscher, P. M. The genetic interpretation of area under the ROC curve in genomic profiling. *PLoS Genet.* **6**, e1000864 (2010).
6. Hazel, L. Principles of a selection index which involves several characteristics and utilizes information concerning relatives. (1941).
7. Daetwyler, H. D., Villanueva, B. & Woolliams, J. A. Accuracy of predicting the genetic risk of disease using a genome-wide approach. *PLoS One* **3**, e3395 (2008).
8. Dudbridge, F. Power and predictive accuracy of polygenic risk scores. *PLoS Genet.* **9**, e1003348 (2013).
9. Wu, T., Liu, Z., Mak, T. S. H. & Sham, P. C. Polygenic power calculator: Statistical power and polygenic prediction accuracy of genome-wide association studies of complex traits. *Front. Genet.* **13**, 989639 (2022).
10. Lee, S. H., Wray, N. R., Goddard, M. E. & Visscher, P. M. Estimating missing heritability for disease from genome-wide association studies. *Am. J. Hum. Genet.* **88**, 294–305 (2011).
11. Vilhjálmsson, B. J. *et al.* Modeling Linkage Disequilibrium Increases Accuracy of Polygenic Risk Scores. *Am. J. Hum. Genet.* **97**, 576–592 (2015).
